# “Medical specialists in LMICs; a systematic review and best-fit framework synthesis of the evidence on their roles and contribution to health systems”

**DOI:** 10.1101/2025.01.06.25320057

**Authors:** Giuliano Russo, Veena Sriram, Tamara Mulenga Willows, Renata Alonso Miotto, Ana Olga Mocumbi, Mário Scheffer

## Abstract

**Background:** Medical specialists are integral to the medical workforce and play a pivotal role in referral systems. However, in low- and middle-income countries (LMICs), there is perception specialists often fail to align with local health needs, system capacities, and Universal Health Coverage (UHC) objectives.

**Methods:** A systematic review was conducted in 2024 using a best-fit framework to assess the contributions of specialists to health systems and population health in LMICs. Searches covered eight databases and specialist journals, guided by an expert-validated *a priori* framework for data extraction and analysis. We used the Johanna Briggs Institute critical appraisal tools to assess the quality of the papers, and the PRISMA guidelines to report the findings. The study protocol was registered in the PROSPERO database (CRD42024572877).

**Findings:** We found and reviewed 89 studies, highlighting a critical shortage of specialists, particularly surgeons, anaesthesiologists, and psychiatrists. Evidence linked specialists’ availability to improved health outcomes such as lives saved through expanded surgical capacity, though broader health system contributions were less clear. Specialists were reported to play key roles in referrals, hospital management, mentoring, and research. Governance of their professions was found to be variable across LMICs, with wide differences in specialty types, training curricula, accreditation systems, and regulation of private-sector involvement. Reports frequently documented specialists’ engagement with private health markets, revealing blurred boundaries between public and private care. A dynamic market for specialists was also observed, driven by a sustained global demand for their services. However, few policies were found addressing shortages and improving governance, with existing strategies focusing on task-shifting, clinical training, and sharing responsibilities.

**Conclusions:** This review offers an evidence-based framework for understanding specialists’ roles and health system engagement in LMICs. We highlight the need to reconsider specialists’ deployment, prioritising alignment with UHC goals and enhancing governance to optimize their contributions to health systems.

## 1 BACKGROUND

Medical specialists – doctors with a recognised advanced qualification and training in a narrower field of medicine – are considered the pinnacle of health system’s referral systems, essential to providing the highest quality of services, medical training, and professional leadership [1]. Nowadays, they also already represent the majority of the medical workforce in high-income countries (HICs) [2].

In low- and middle-income countries (LMICs), where opportunities for training and resources are scarcer, the number of specialists are steadily increasing, driven by growing demand for specialised services as well as doctors’ own interest to increase their career options [3]. However, types of medical specialties and ratios of specialists to doctors without a further specialization vary considerably across such countries [4].

From a theoretical standpoint, specialist healthcare services seem ostensibly at odds with LMICs’ specific health needs and health systems; here, primary care services and cost-effective health staff are key for working closer to communities, often in rural areas, and realising Universal Health Coverage (UHC) goals [5]. Most specialists are in fact known to work predominantly in urban settings, tertiary care level hospitals, and engage often with the less affordable private sector market [6]. Consistently, evidence from the United States suggests that health systems relying excessively on specialists would neither be cost efficient or improve population health outcomes [7].

Although primary care is firmly at the centre of UHC and strengthening health systems in resource-scarce settings [8], attention has also been drawn on the equity implications of basic care-only-focussed health systems, and the perils of creating lower-quality services for the poor [9]. A growing consensus exists that PHC also comprises essential specialist services, such as life-saving, cost-effective surgical procedures [10] and emergency care [11]. Scholars from LMICs have also contested the goal of securing universal coverage of basic services as opposed to access to universal systems [12], and recognised the importance of specialised services to realise Sustainable Development Goals and health rights [13], and to consolidate the professions through adequate planning [14].

From a sociology of professions perspective, specialization is often seen as a trajectory that the medical profession is increasingly built around [2]; rather than leaving it entirely to market forces to shape contents and contours of specialties, some interventions from decision makers (government, regulatory agencies or colleges) might be needed to ensure that this growth takes place alongside health goals for the country.

## 2 METHODS

The aim of this literature review is to take stock of the arguments, theoretical aspects, and evidence on the role of medical specialists in health systems in LMICs. To lend substance to such arguments, we explore the specific evidence on specialists’ contribution to health systems strengthening across its pillars[15], and to provision of services in resource-scarce settings.

The research questions to be answered by this review are as follows:

1. What is the contribution of medical specialists to population health and health outcomes in LMICs?
2. Within national healthcare systems, what is their contribution to provision of services and system strengthening in such contexts?
3. What are the factors and drivers of specialists’ contributions in resource-scarce settings?

### 2.1 Approach and design of the review

We used a ‘best-fit framework’ synthesis approach [16] to organise the initial searches, iteratively refined to accommodate the empirical evidence uncovered, and develop a theoretical interpretation of our findings on medical specialists’ contribution to health system strengthening and population health in LMICs.

Our a priori framework drew from previous conceptualizations of specialists contribution to health systems strengthening [1,2,13], and was validated by a panel of experts from high- and low-income countries (see the list of panel members in Appendix 1). Our framework identifies specific functions that specialists typically perform that contribute to health system strengthening and population health – from provision of specialistic services, to referral for complicated cases, or contribution to medical research, teaching, and policy elaboration. We understand such functions are shaped by the current governance of the medical area in the country, and by the organisation/regulation of individual specialties. Underlying health labour market conditions and specialists’ motivation to choose their specific area of practice are likely to influence both the governance of the professions and the functions carried out by specialists in different countries (Figure 1 below).

**Figure 1:**
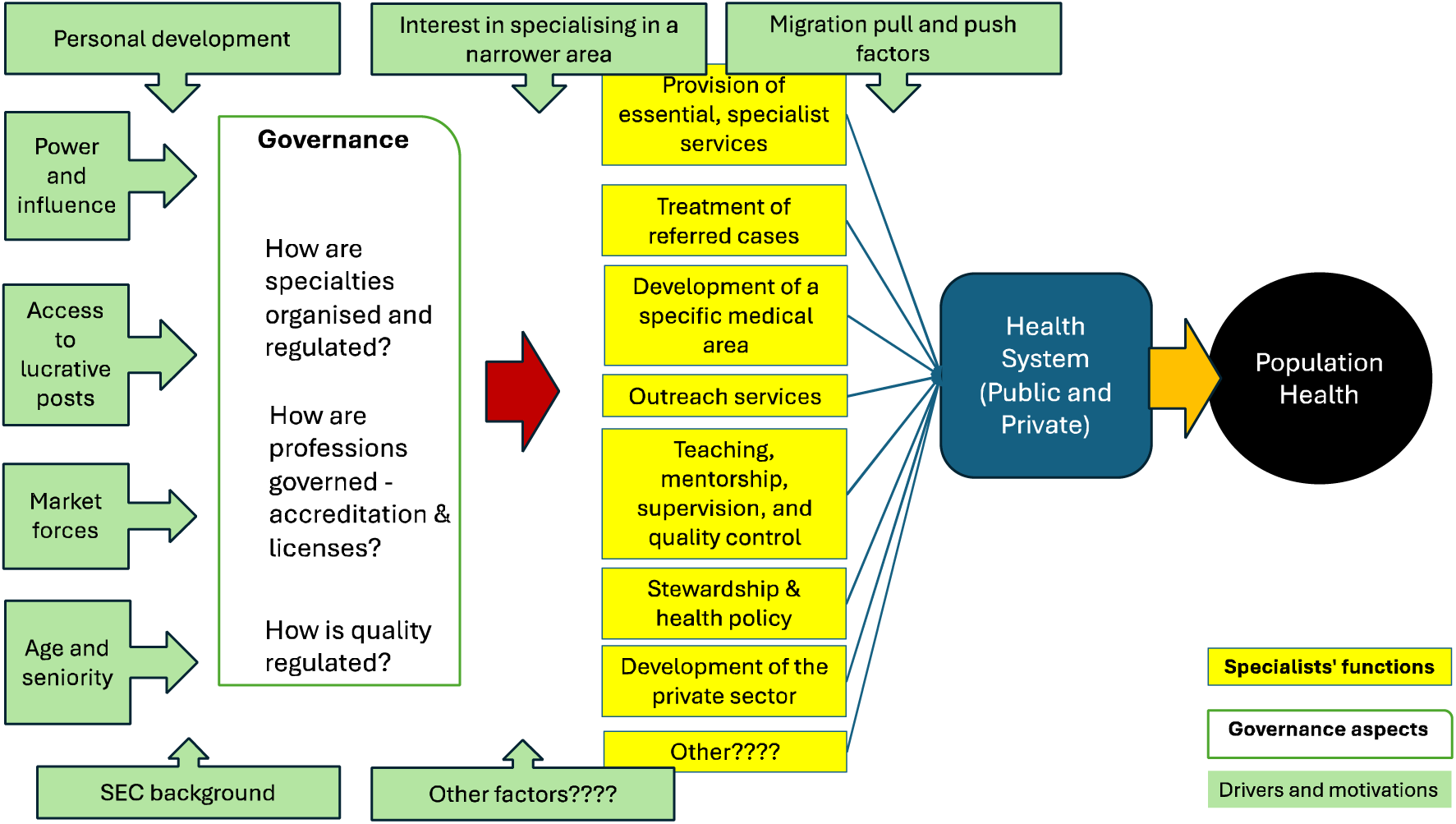
A priori framework on contribution and governance of specialist doctors for health system strengthening in LMICs.

The study protocol was registered in the PROSPERO database after the elaboration of the a priori framework, and before the formal screening of the identified studies [17] – registration number CRD42024572877.

Our outcomes of interests referred to the relevant parts of the conceptual framework (see Fig.1 above), in relation to specialists’ impact on population health and health systems, their functions as described in the literature – referral of services, teaching, research etc – drivers of the professions, and governance of specialties (see Appendix 2 for the full list of outcomes of interests and search terms).

### 2.2 Search strategy

We searched eight relevant databases across the health, health systems and economic literature (PubMed; ISI Web of Science—core collection; Scopus; Cochrane Library; PDQ-Evidence; Health Evidence.org; Scielo; and Econ Lit). We separately searched the websites of specialist journals publishing on our topic, such as The Lancet Global Health, BMJ Global Health, Social Science and Medicine, Health Policy and Planning, Human Resources for Health, and The International Journal of Health Policy and Management.

Searches for specialist doctors included general terms such as: “specialist*”, “medical special*” as well as terms of specific specialties highlighted as ‘essential’ for providing universal health coverage in resource-scarce settings by the Lancet Non-Communicable Disease (NCDI) Poverty Commission [18] (such as Gynaecolog* OR Cardiothoracic surg* OR Surg* OR “Internal medicine” OR Critical care OR Palliative care OR Oncolog* OR Cardiolog* OR Ophthalmolog* OR Patholog* OR Radiolog* OR Rehabilitation OR “Orthopaedic surg*” OR Otorhinolaryngolog* OR Neurolog* OR Psychiatr* OR Paediatric* OR Obstetric*) – see Appendix 2 for detailed search strategy and terms.

We used the OECD Development Assistance Committee’s list of Official Development Assistance recipients to identify low- and middle-income countries, which as of 2024 includes 47 least-developed and low-income countries, 35 lower-middle income, and 59 upper-middle income countries and territories [19].

### 2.3 Eligibility criteria

We included academic literature published in English, Portuguese, French, Spanish or Italian, based on primary empirical evidence from quantitative, qualitative or mixed-methods studies, or original analysis of secondary data (including systematic reviews). We excluded: unpublished study reports, papers published in non-peer-reviewed journals, commentaries and opinion pieces, non-systematic reviews, and papers with no title or abstract in any of the five languages above.

For the purposes of this review, drawing from the medical science literature [7,20,21], we adopted the following definition of specialist physicians: “Medical doctors with a recognised advanced qualification in a <u>narrower field</u> of the medical science, who operate primarily from a hospital setting.” Conversely, generalist doctors were defined as those <u>without</u> a further qualification beyond the statutory medical degree, with undifferentiated experience across multiple clinical specialities, and those recognised as primary care doctors working with both adults and children, practicing closer to communities rather than in hospital settings.

We considered publications focusing on: (1) organisation, regulation and policies on medical specialties and specialty services in LMICs; (2) contribution of specialty services to health outcomes and population health; (3) population access to specialty services; (4) medical specialists’ functions and performance in LMICs; (5) contribution of medical specialists to health system strengthening (see the inclusion and exclusion criteria section in Appendix 3).

### 2.4 Data management, selection process and quality assessment

Bibliographic references were managed in the open-source Zotero software; characteristics of each study and respective key findings were organised in an Excel database and analysed through pivot dynamic tables.

After being shared through Zotero software and de-duplicated, all identified references went through titles-and-abstracts screening (Level 1 screening), performed on a random sample of 5% of the entries by one researcher with systematic review experience. Search terms were refined accordingly. Level 2 screening (full-text assessment) was divided among three authors.

The database searches were conducted by TMW and RAM. The initial Level 1 screening was conducted by GR to identify sources that may be of relevance to our study objectives. Level 2 screening was conducted by TMW, RAM and GR, with each reviewer independently looking at the abstract and full text to assess whether the source met the inclusion criteria. The Level 2 reviewers returned three possible decisions: ‘included,’ ‘excluded’ or ‘Uncertain: seek further information.’ For each source that was excluded, the reviewers recorded a reason from related to the inclusion and exclusion criteria. For each source that was judged as ‘seek further information’, a third reviewer (GR) made an independent assessment regarding whether the sources should be included.

Where studies met the eligibility criteria, their methods’ quality was also appraised using the Joanne Briggs Institute’s critical appraisal tools covering 12 different types of study designs [22]. The reviewers who performed the Level 2 screenings also performed this assessment, using one checklist per paper, appropriate for the study design.

### 2.5 Data collection and data items

A data extraction form was populated based on the a priori conceptual framework and covering key methodological and contextual features (such as type of study, type of evidence, type of specialist, impact on health system) for the included studies. The rubrics of this form guided the extraction of relevant empirical evidence and the refinement of the a priori framework. The contents of papers were extracted independently by three reviewers, without the use of a specific coding structure.

The quantitative and qualitative findings from the included papers were coded and synthesized against the a priori framework, which was refined throughout the process to produce the ‘best-fit’ framework [16]. Whenever the empirical evidence did not match the categories of the a priori framework, we revised the framework to accommodate emerging themes and diverging evidence, particularly for different contexts [23].

## 3 FINDINGS

Our initial searches generated 2,722 records from eight databases. After removing duplicates and level 1 screening, we assessed 174 records for eligibility. 55 records were excluded because not relevant to our inclusion criteria, and 30 for low quality, following the JBI’s assessment criteria. 89 records from 80 studies were eventually included in this systematic review (Figure 2 below). See a list of the papers excluded, with reason in Appendix 4.

**Figure 2:**
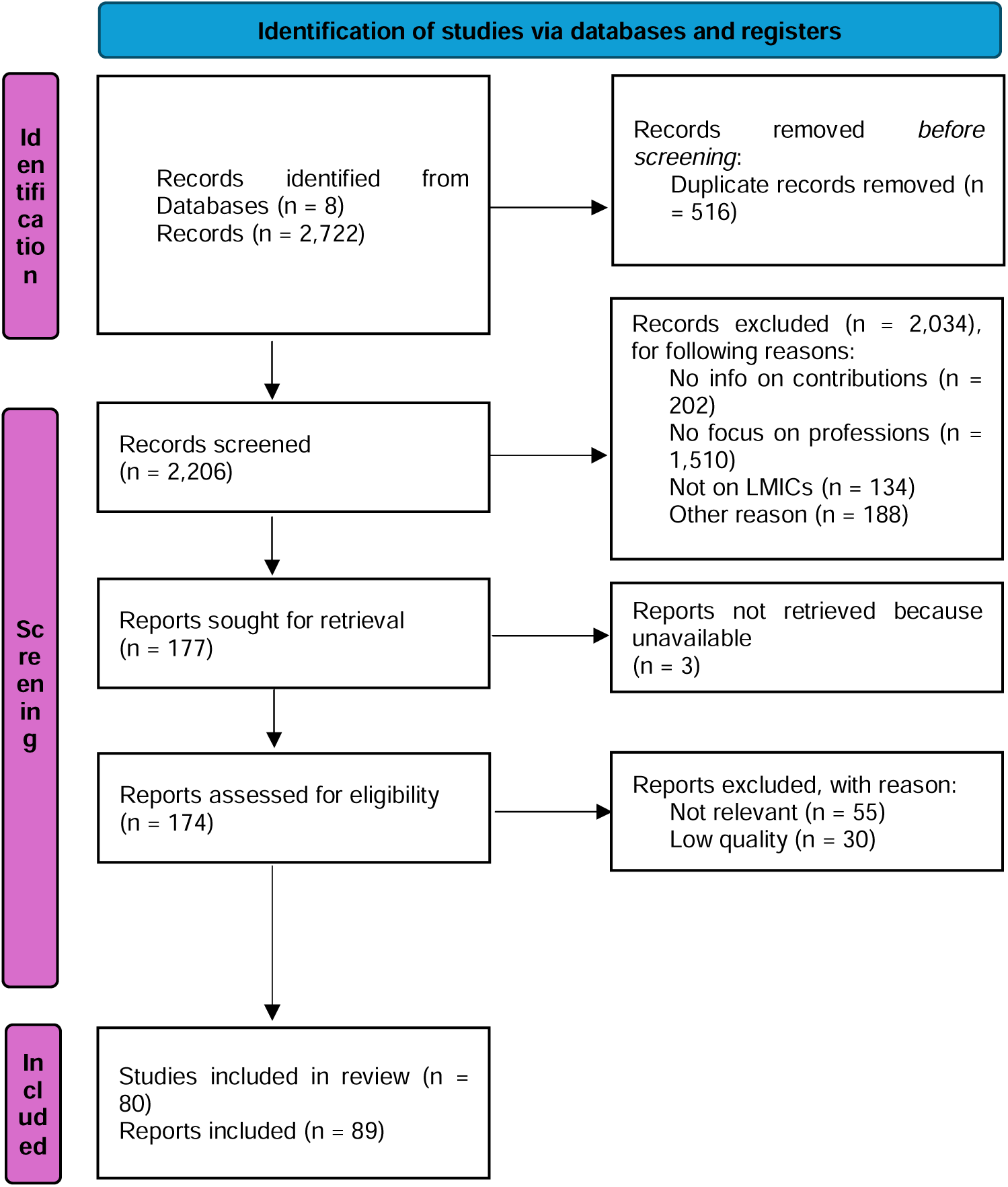
PRISMA flow diagram for searches, assessment and selection of the literature.

The papers retained focussed either on low-income countries in general, or on specific countries from Sub-Saharan Africa (SSA). Although many papers (28) discussed medical specialists in general, 27 focussed specifically on surgeons and anaesthetists jointly as parts of surgical teams, and 6 on anaesthetists only. Medical specialty students were the subject of 6 papers, psychiatrists of 5, followed by other specialties like obstetrics/gynaecology and paediatrics (4 papers each) – see Figure 3 below.

**Figure 3:**
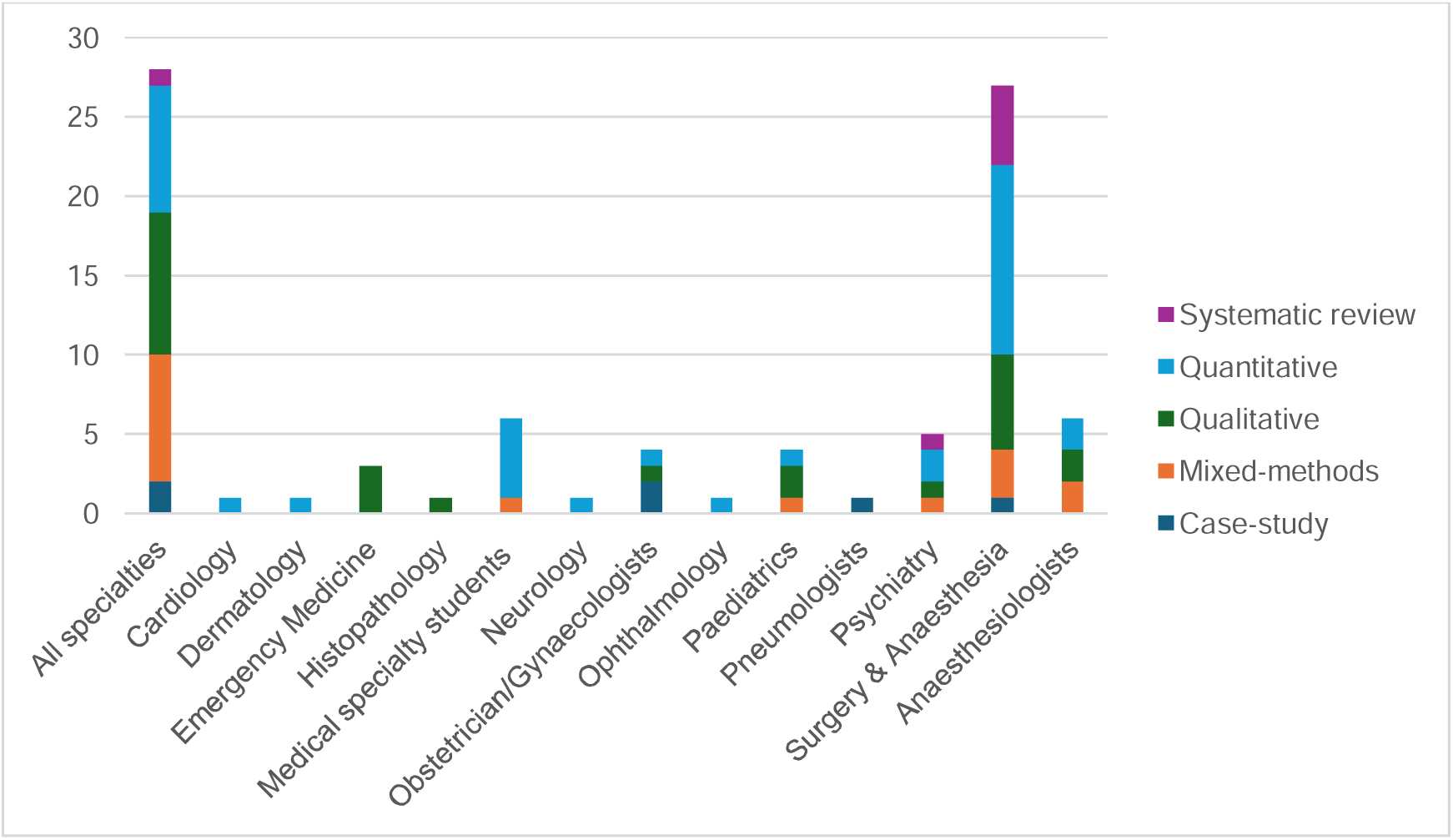
Papers included in the review, by medical specialty and methods.

Of the papers eventually included, 35 were based on quantitative analysis of census data, surveys, or discrete choice experiments, and modelling exercises. There were 25 qualitative papers, 16 mixed-methods, and 6 case-studies reporting on specific medical specialty development experiences. Four main bodies of evidence emerged, covering scarcity of specialists in low-income settings, their contribution to health systems and population health, their governance, and their interaction with health markets (see Table 1 below).

**Table 1:**
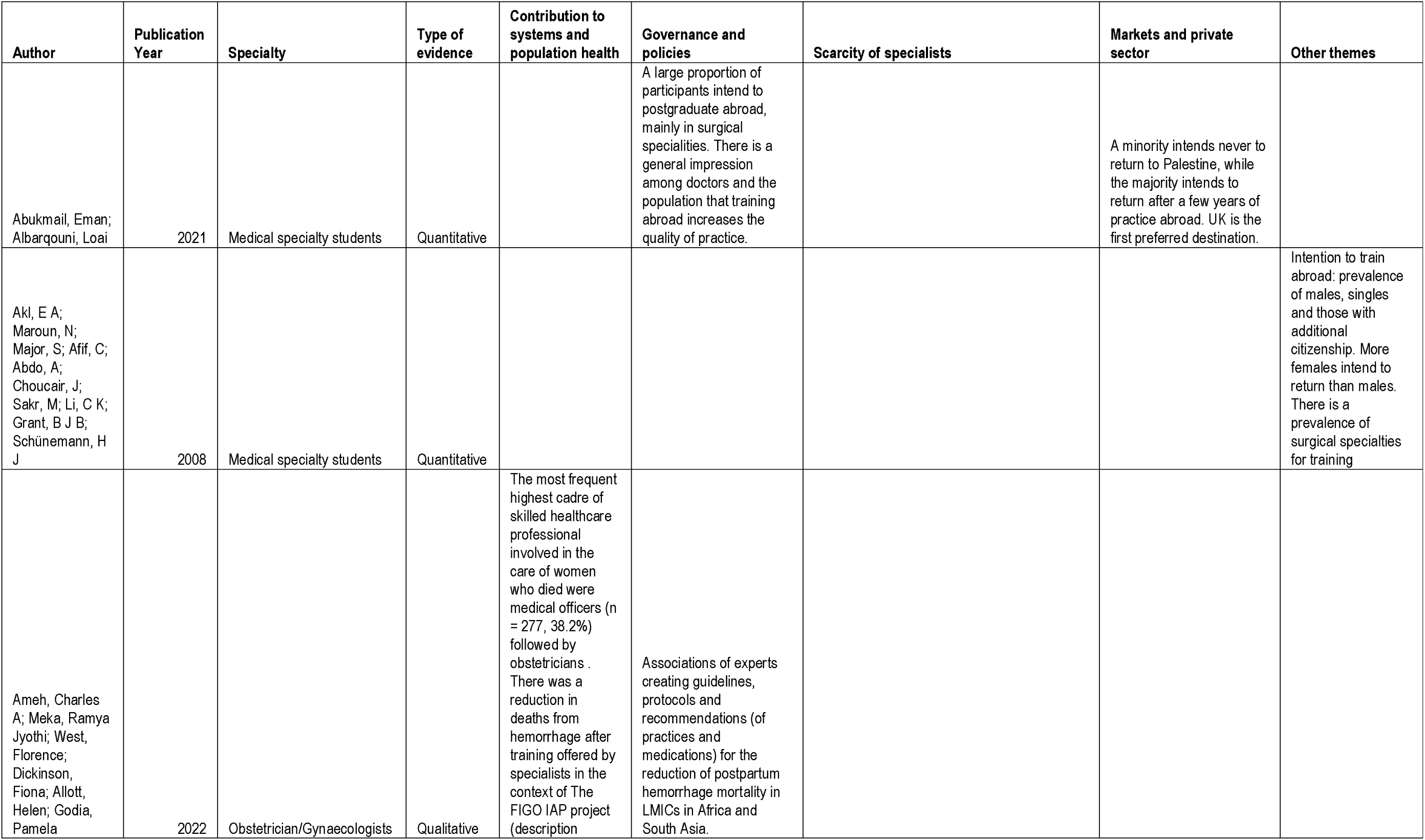
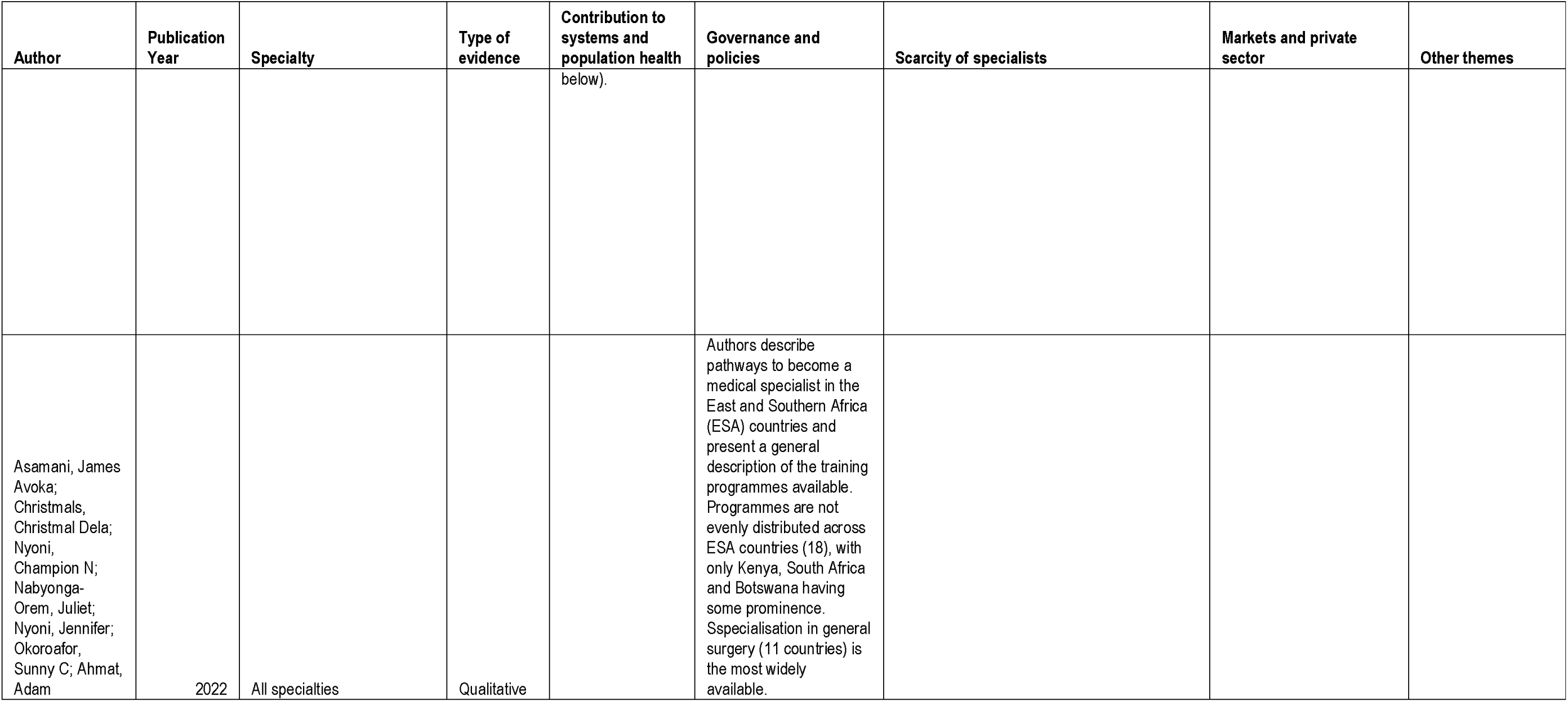
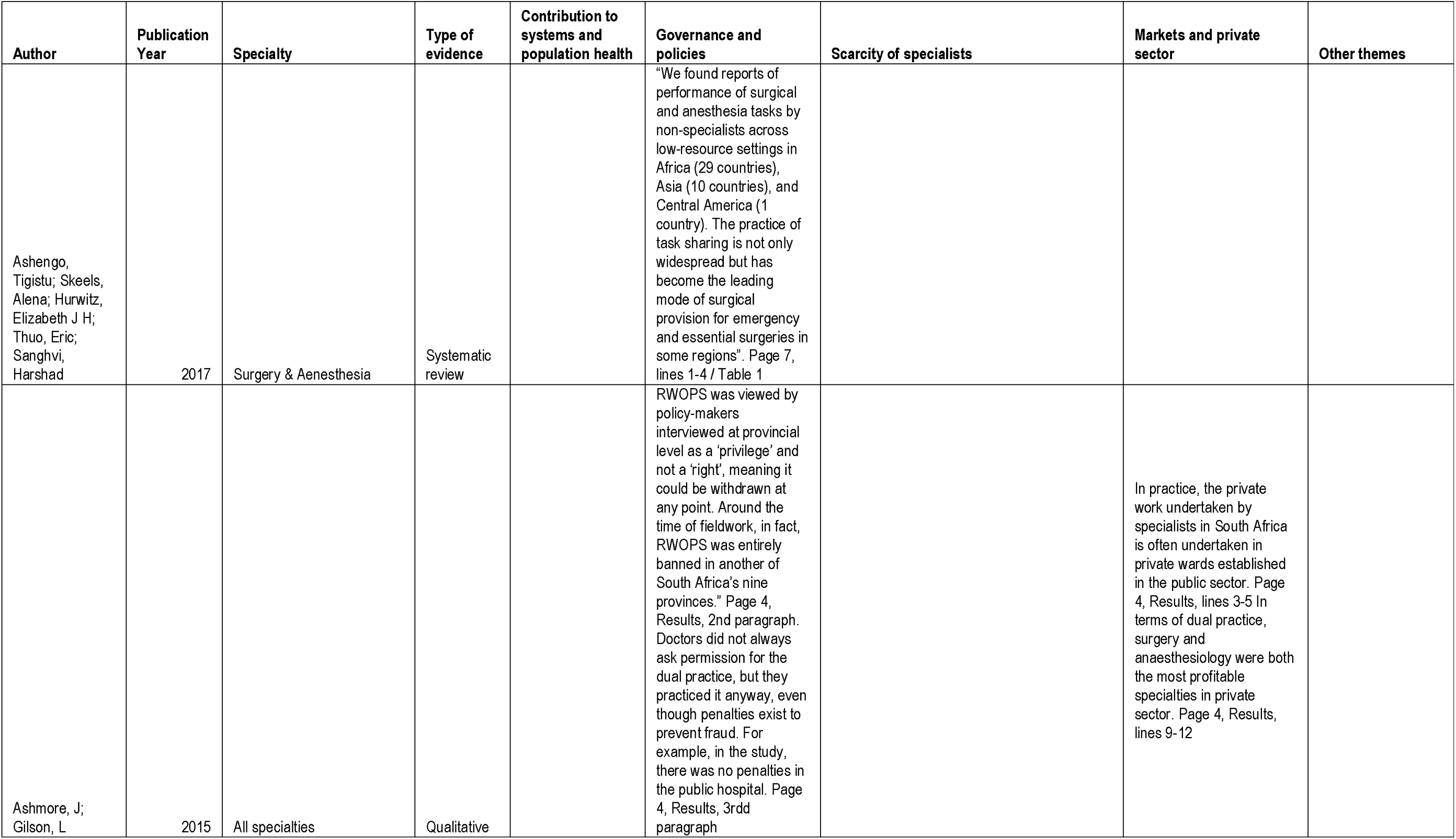
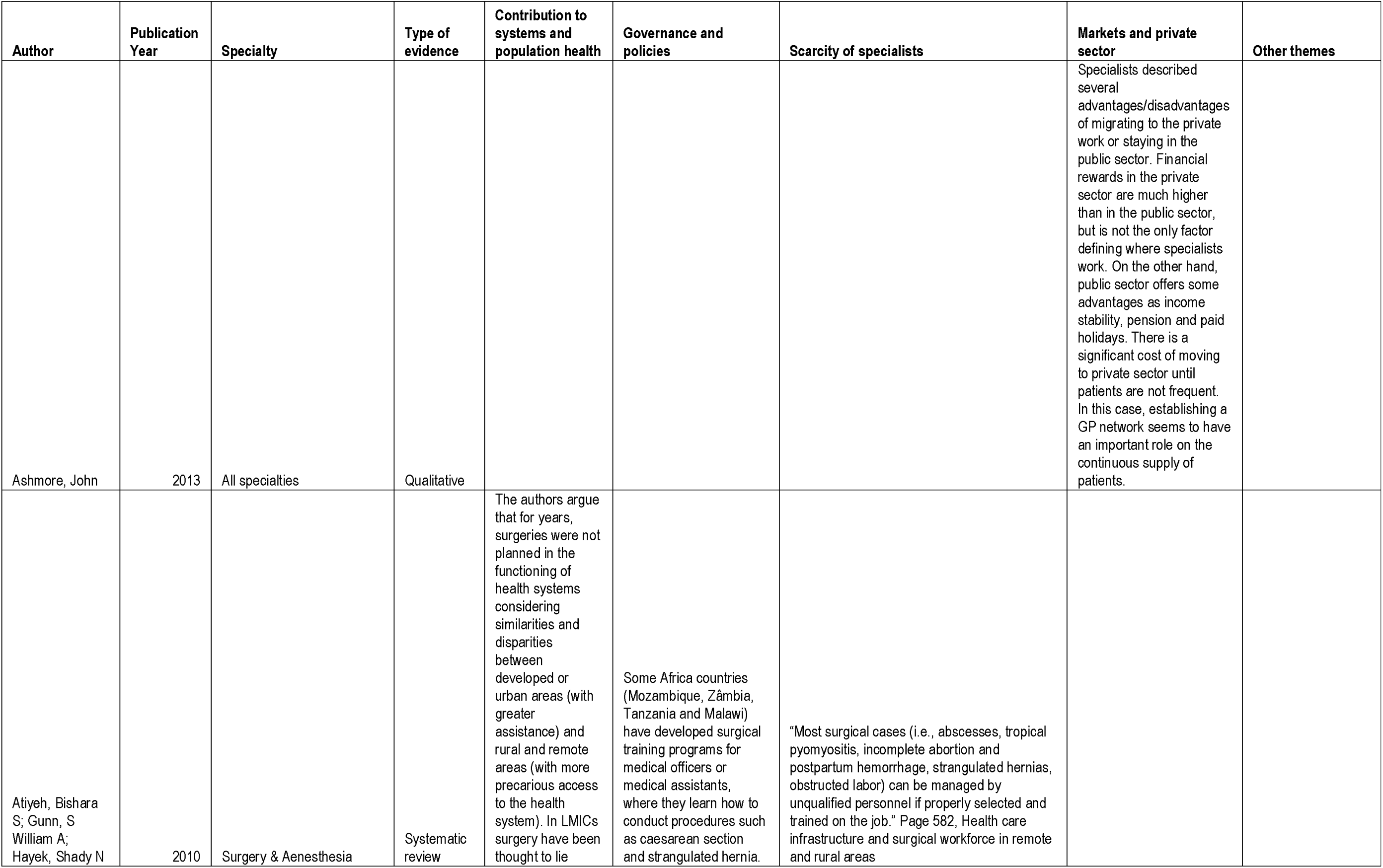
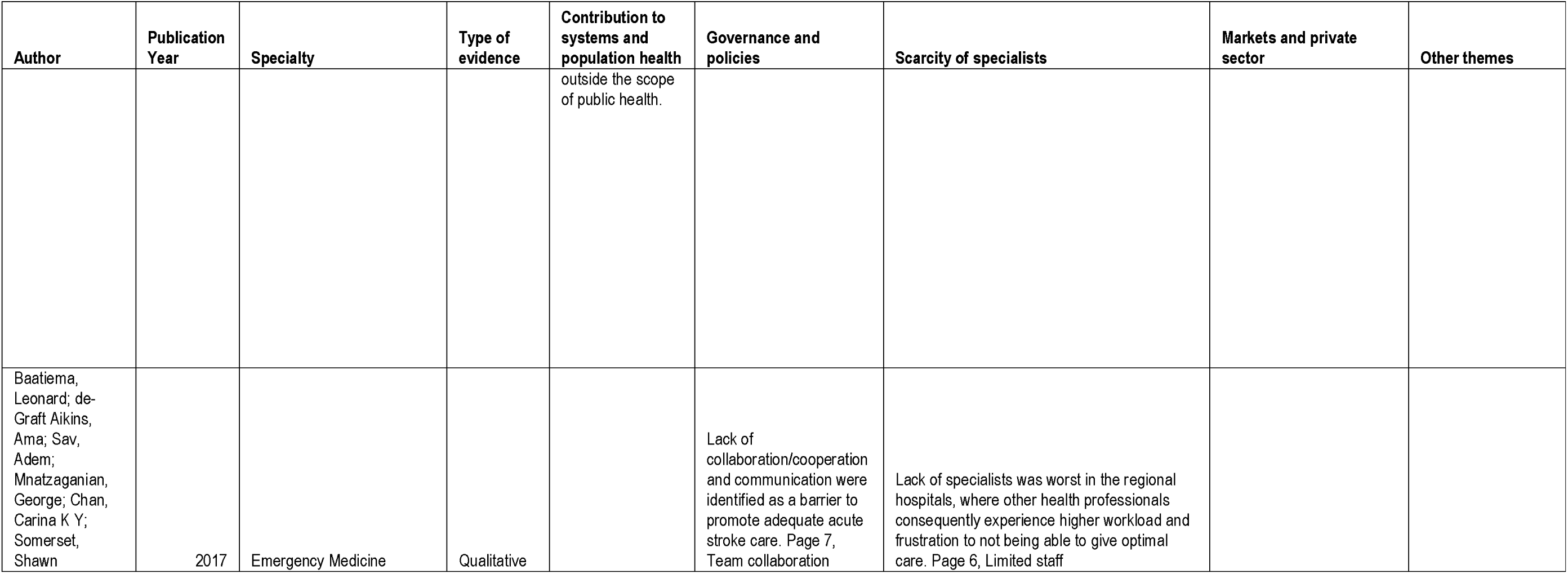
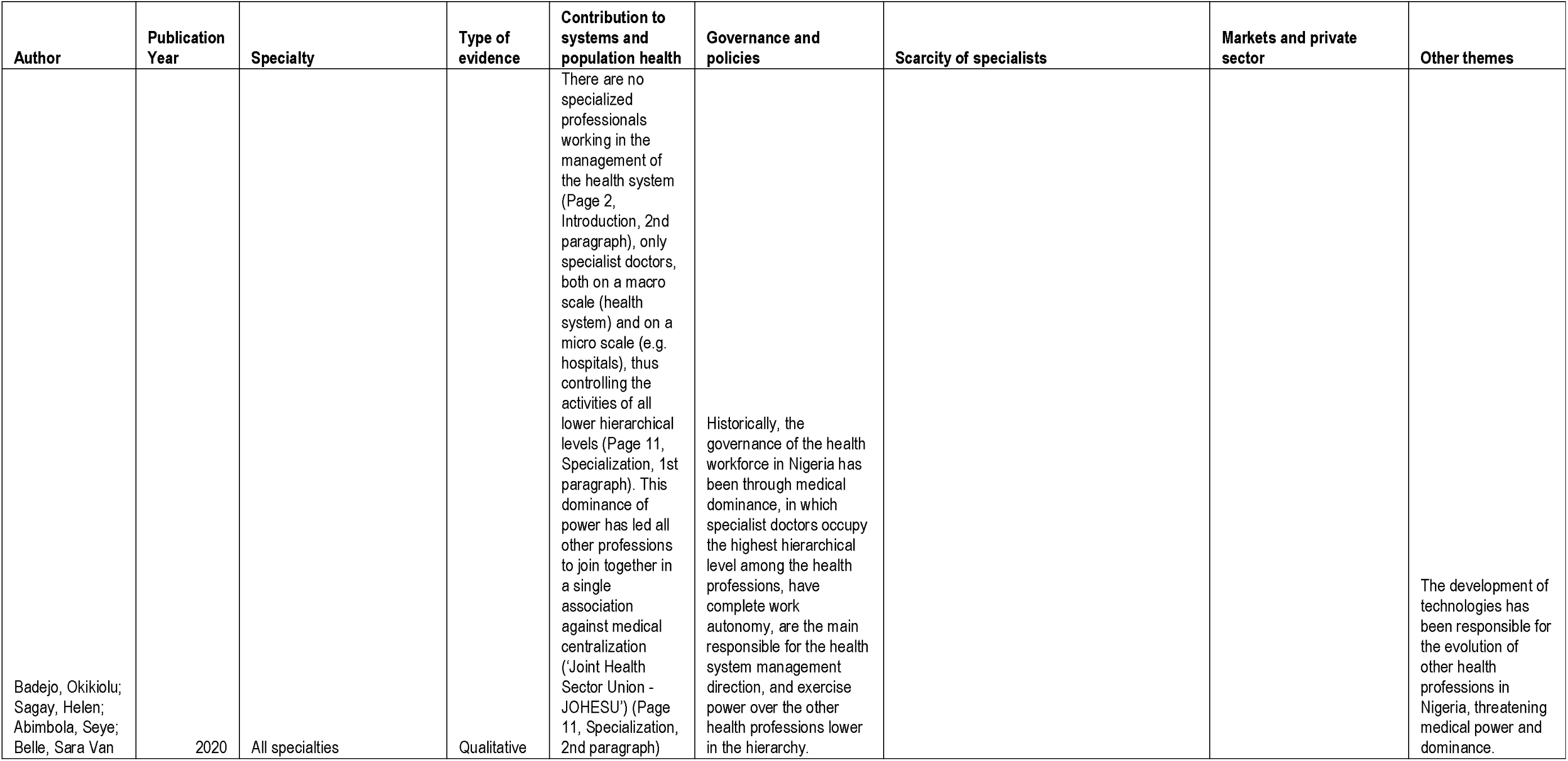
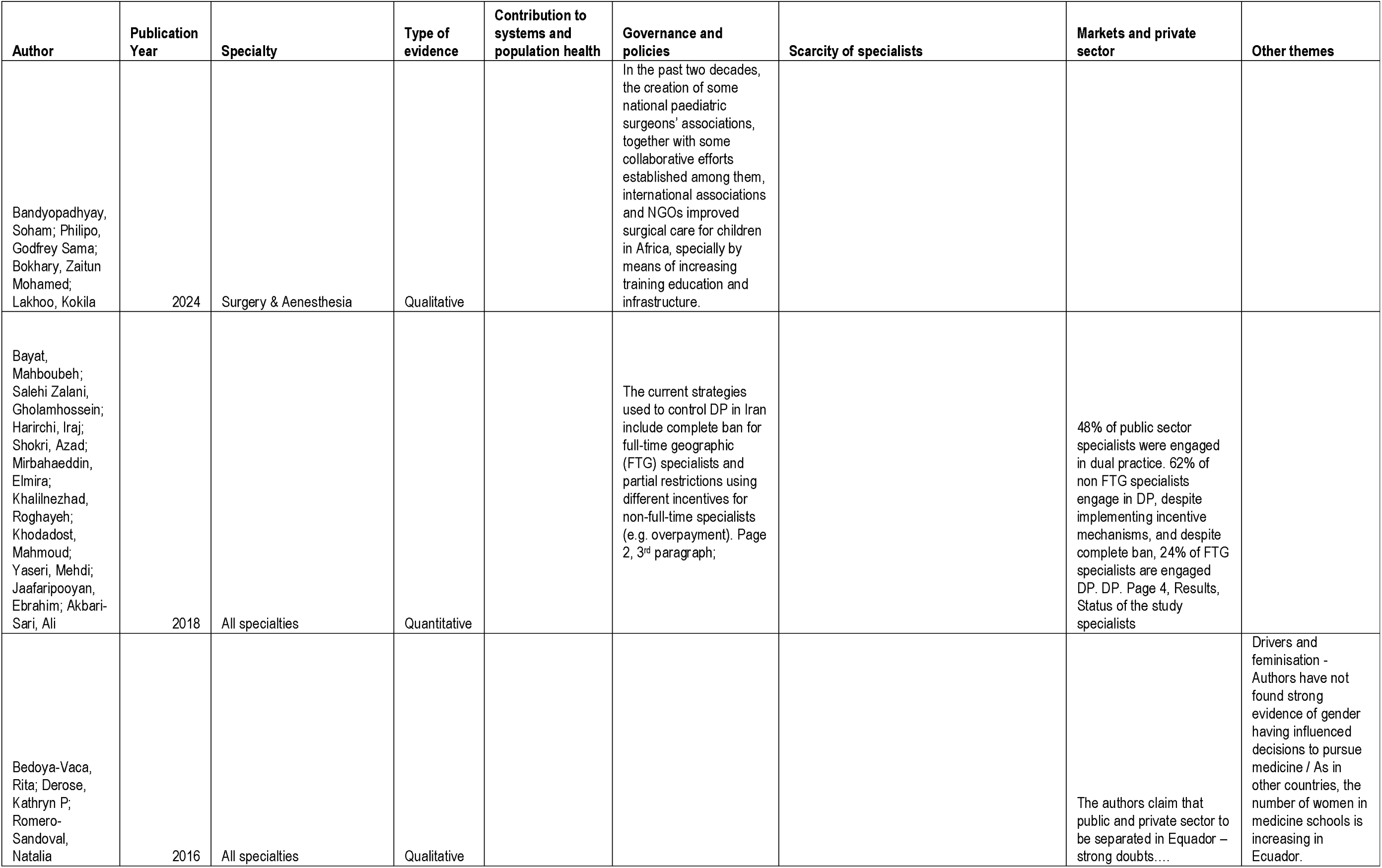
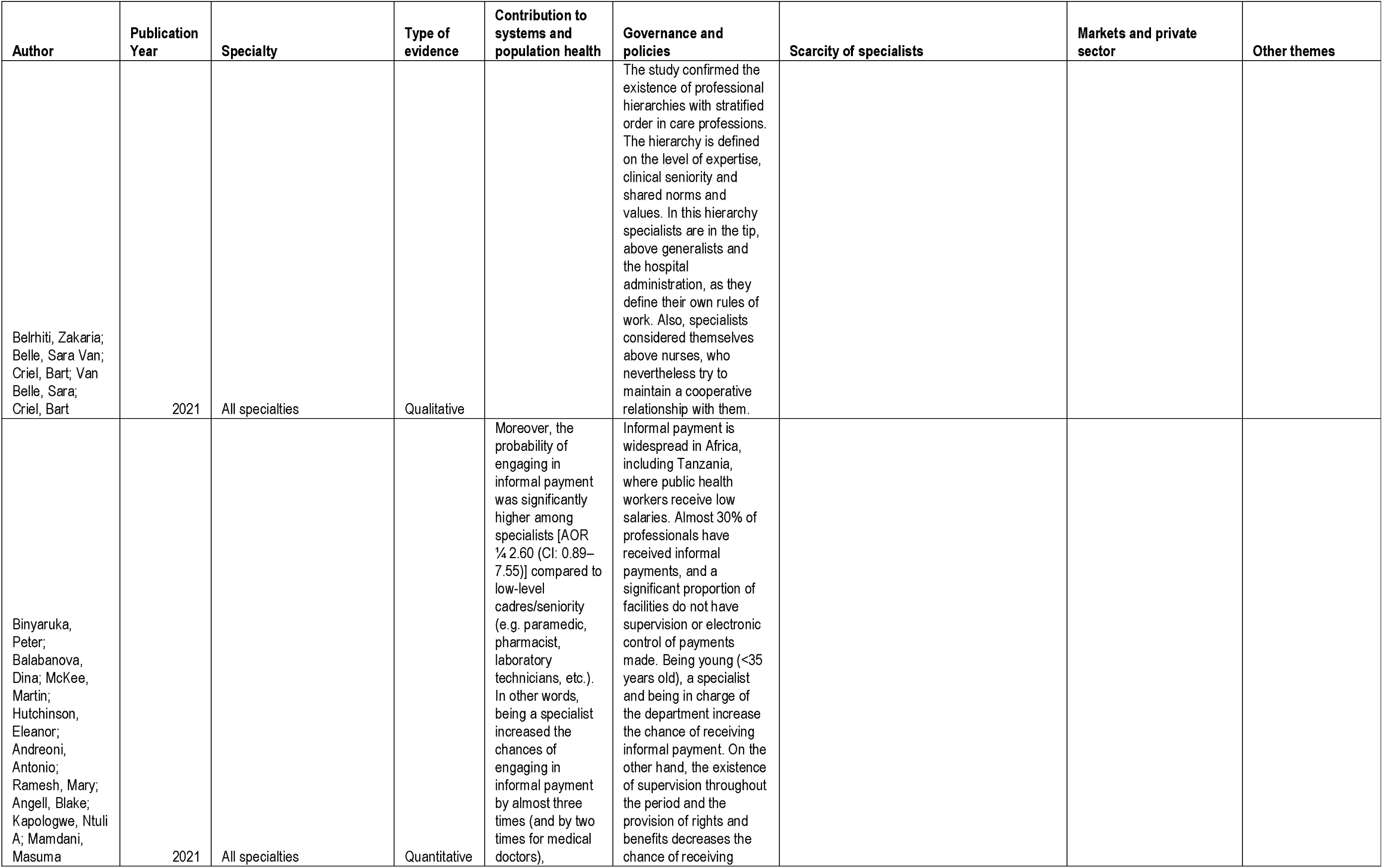
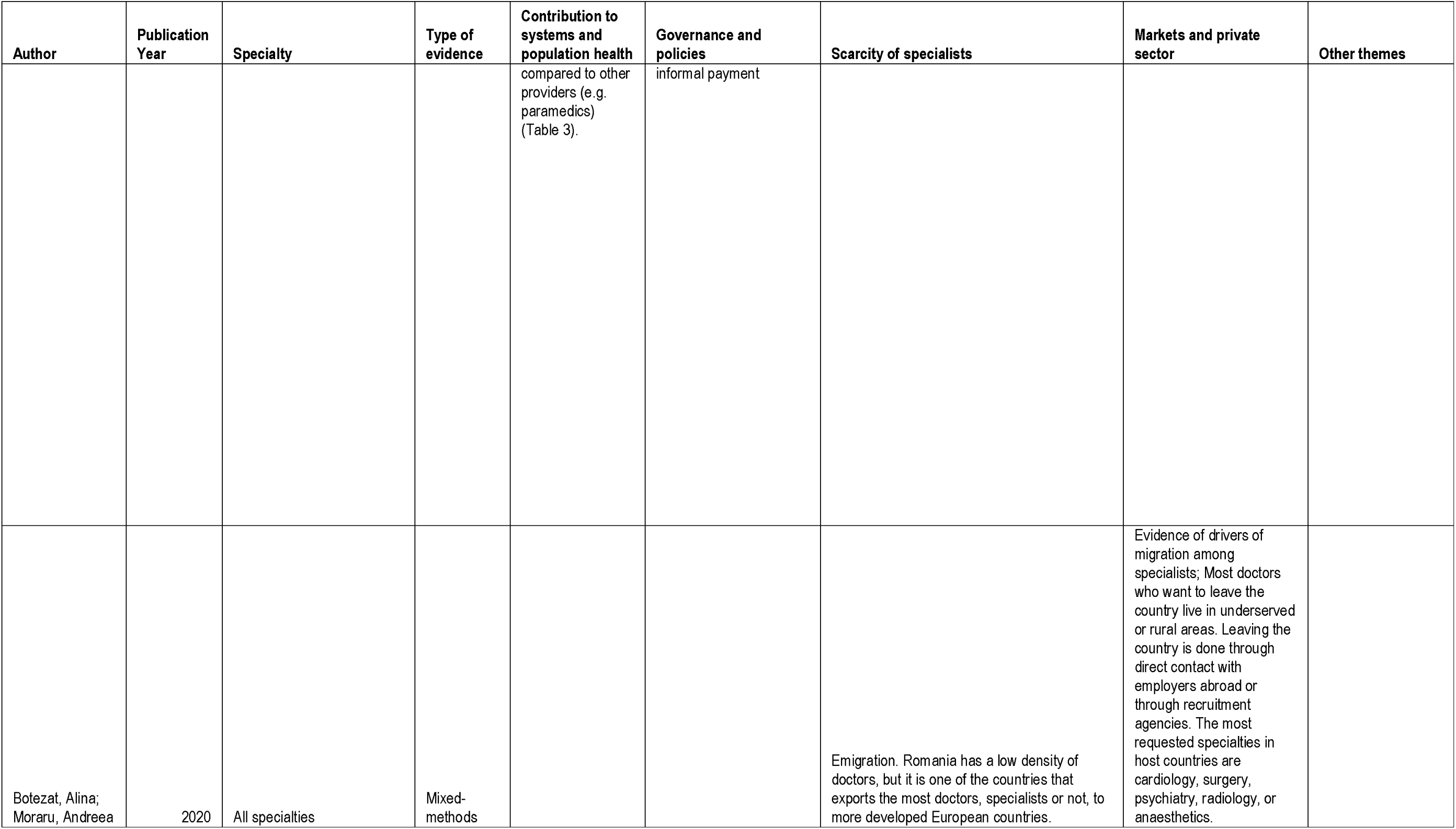
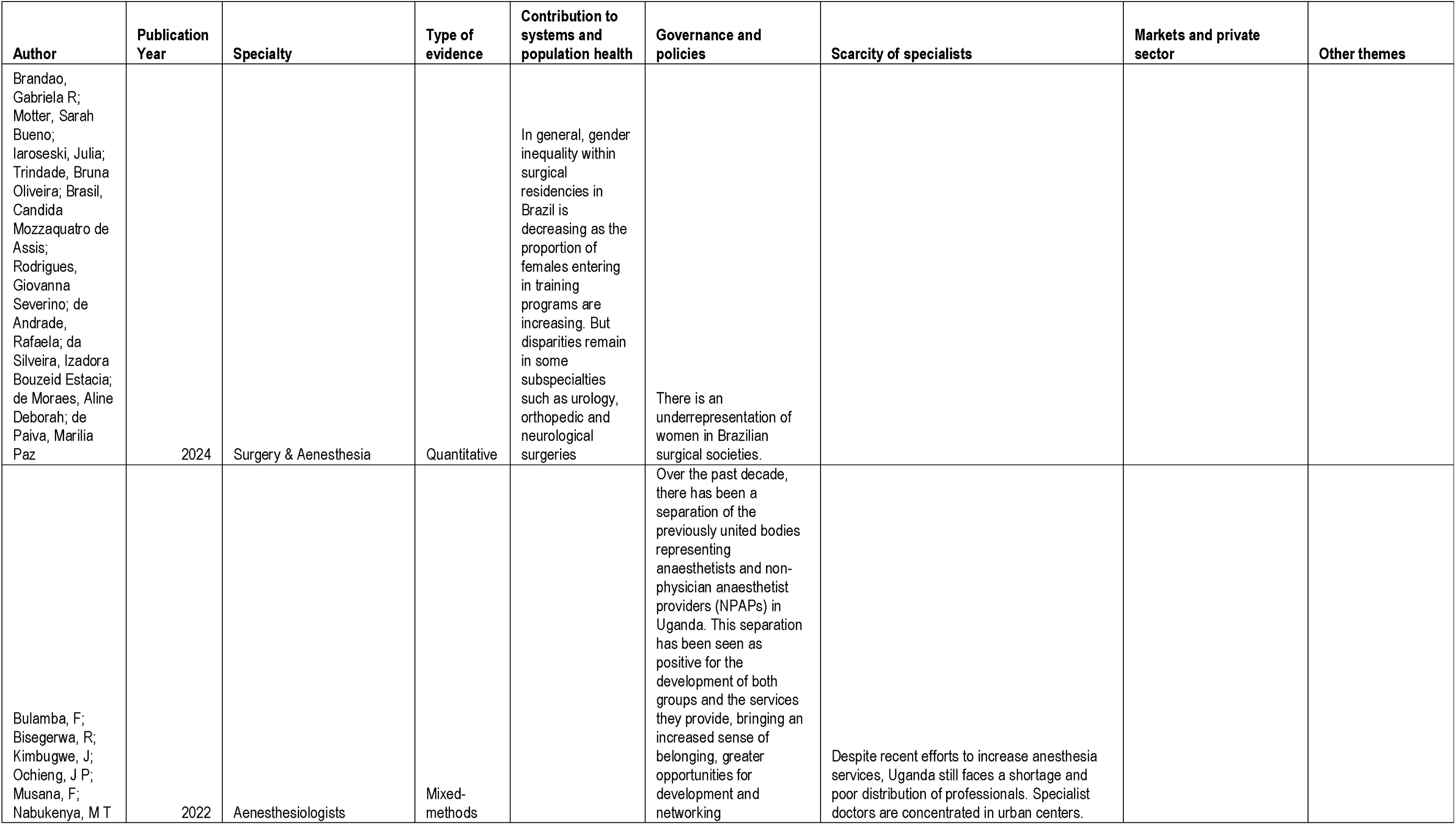
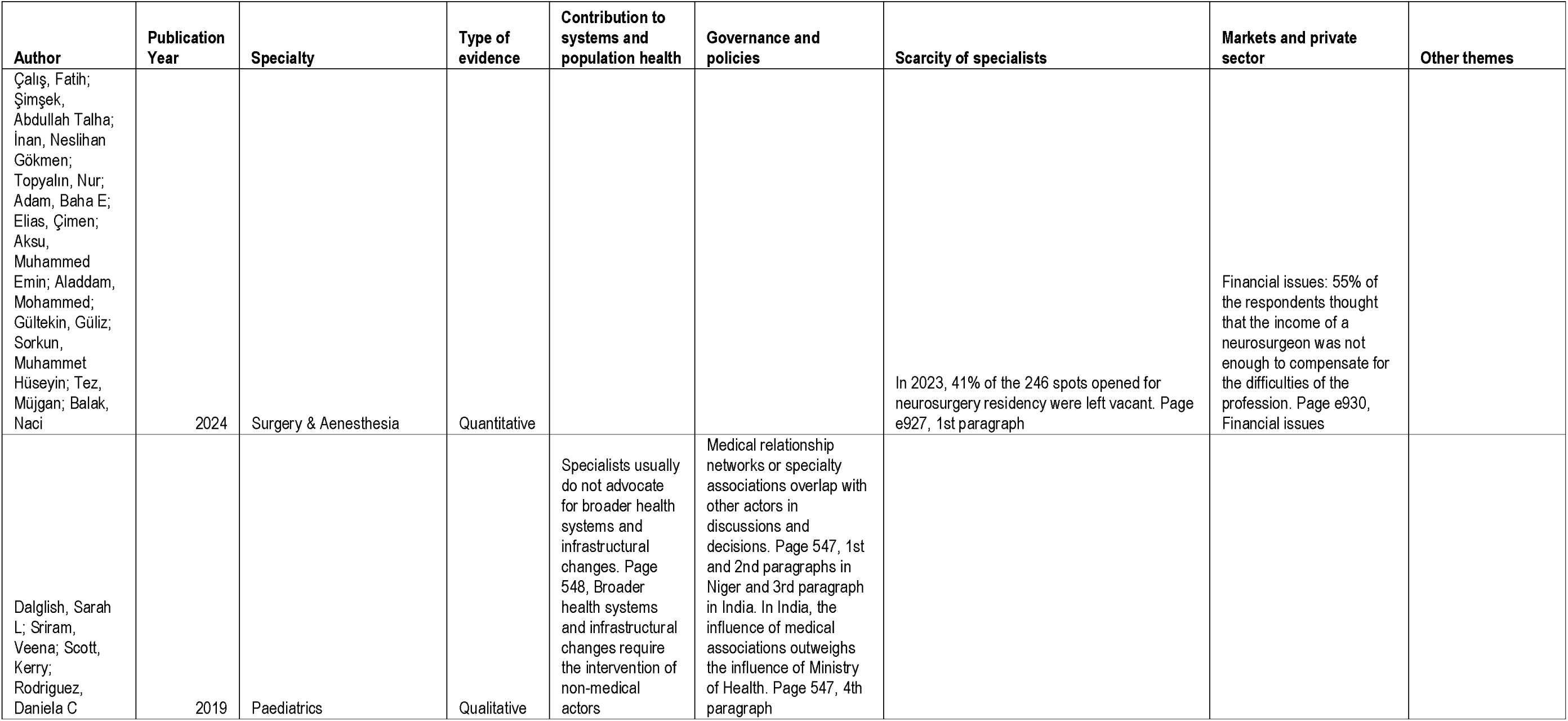
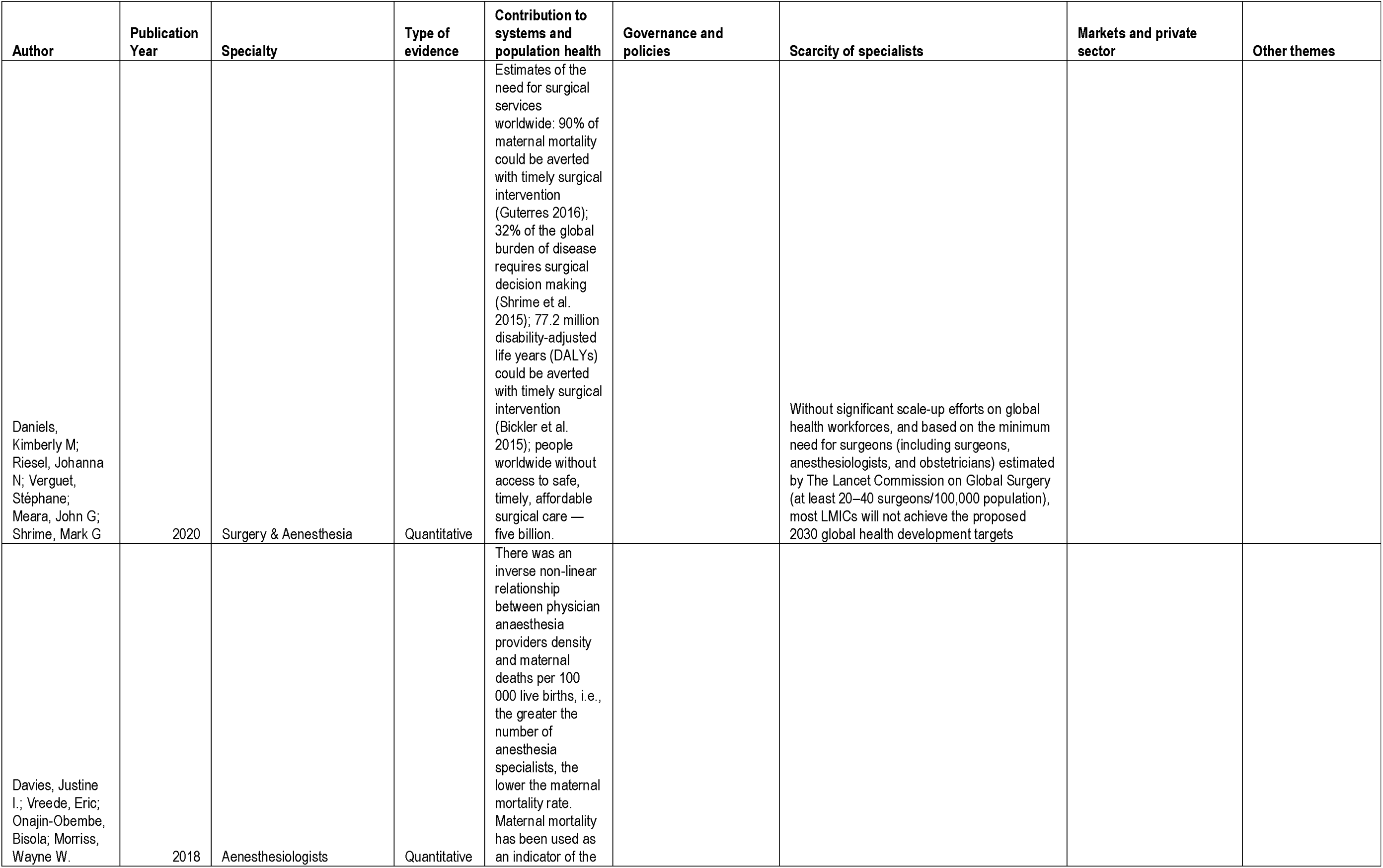
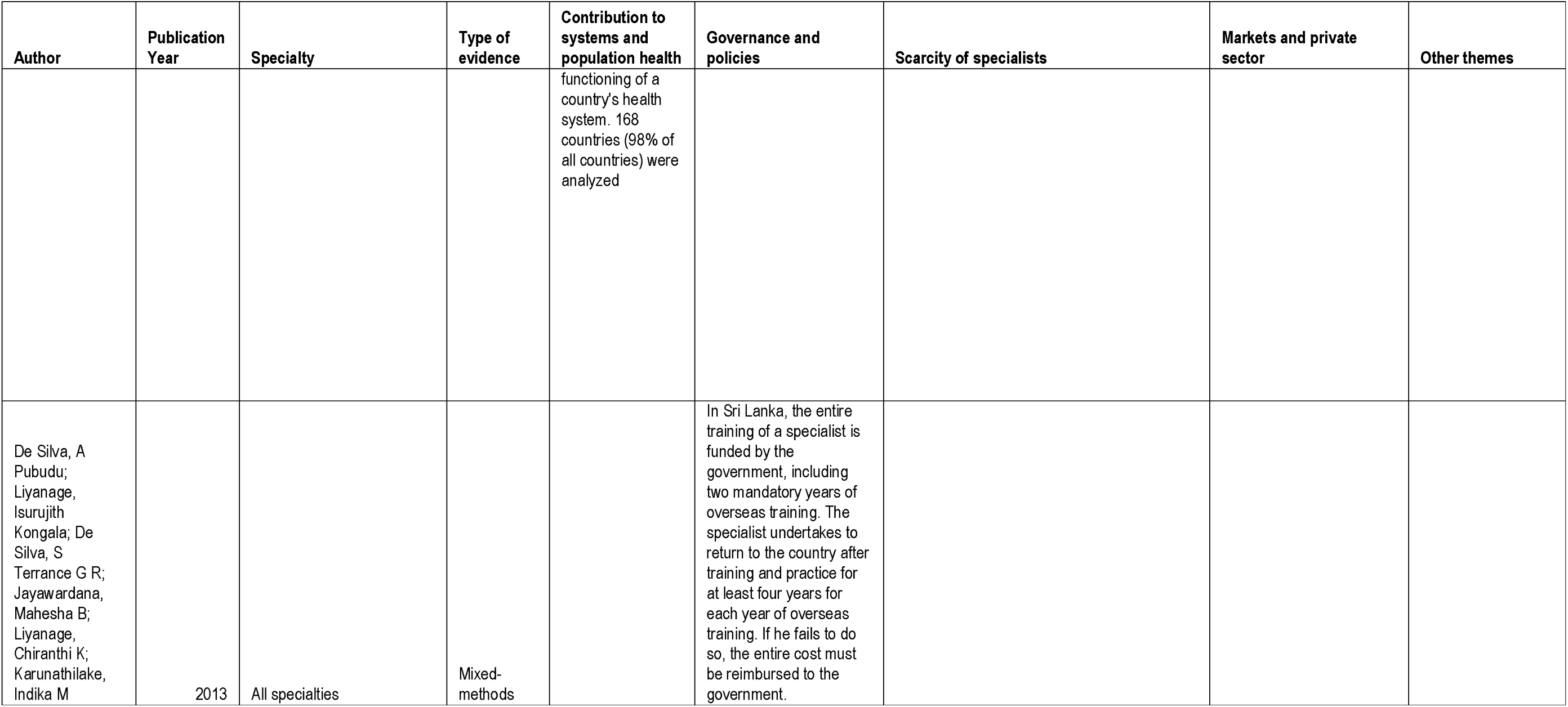
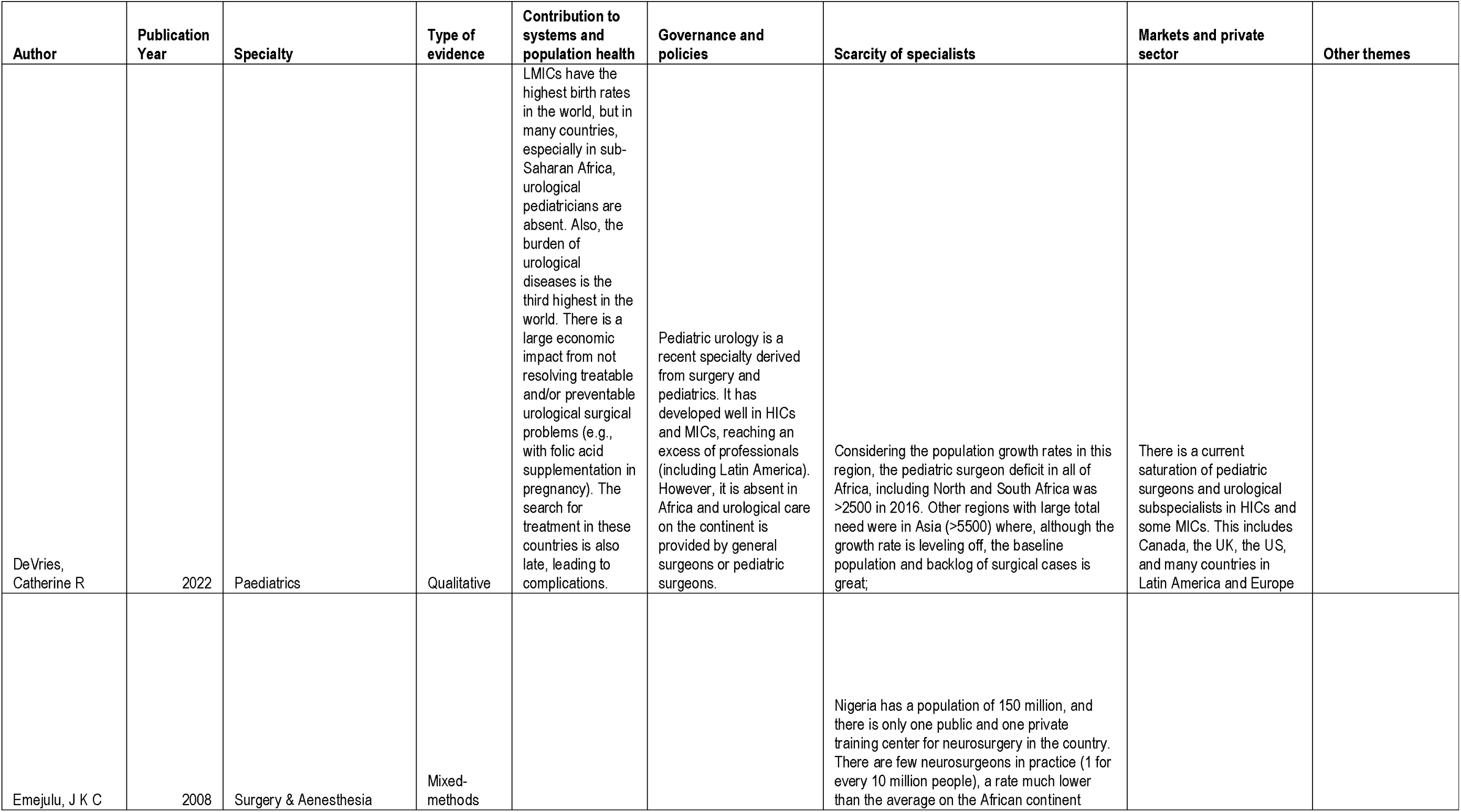
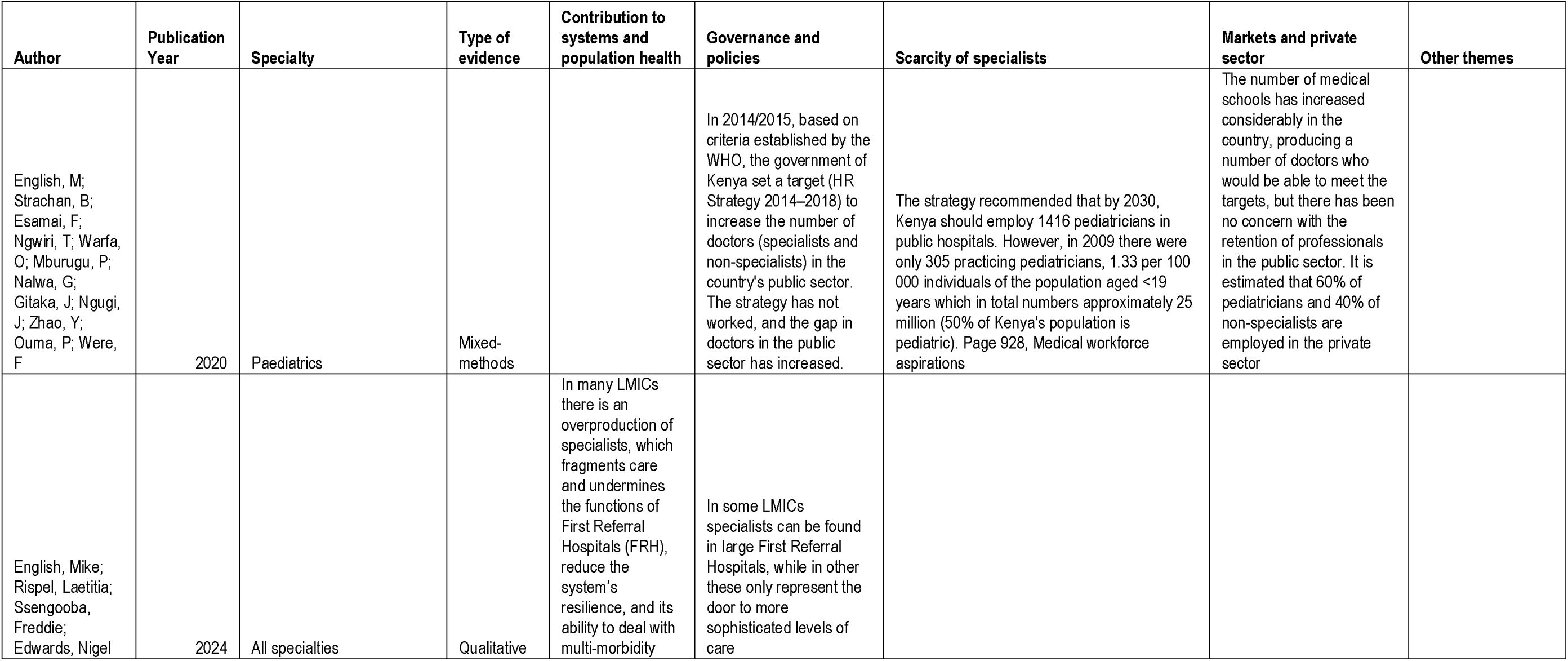
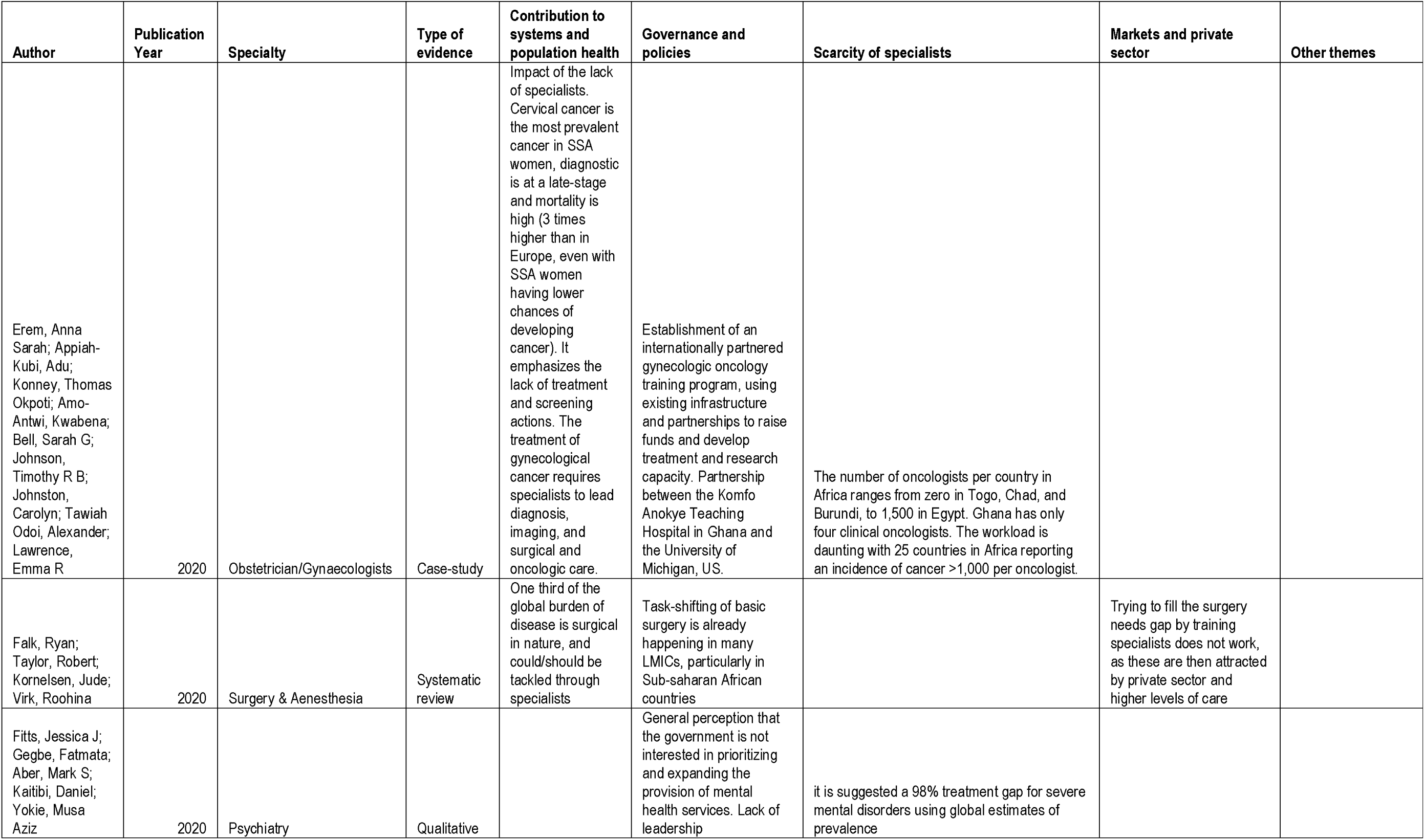
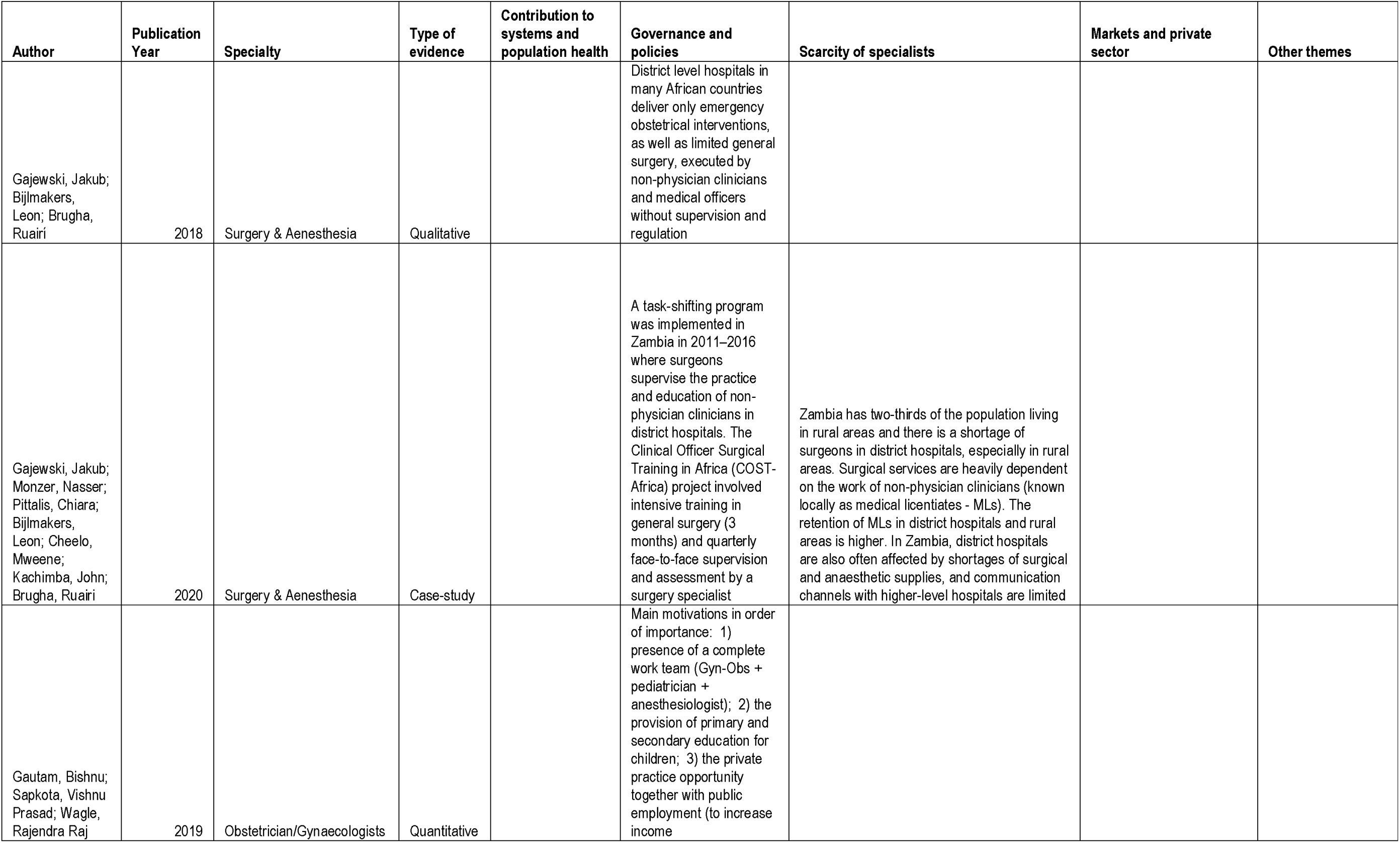
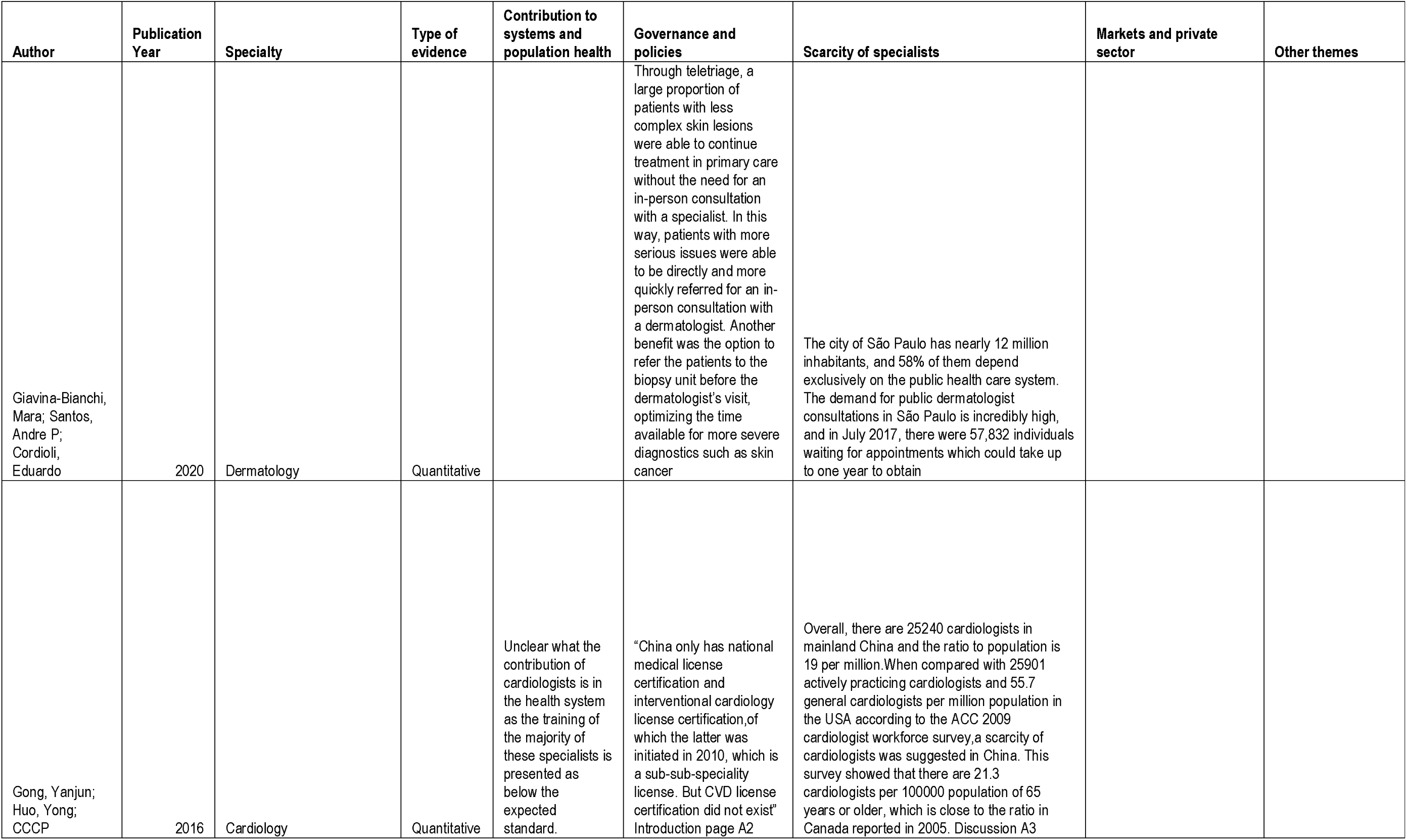
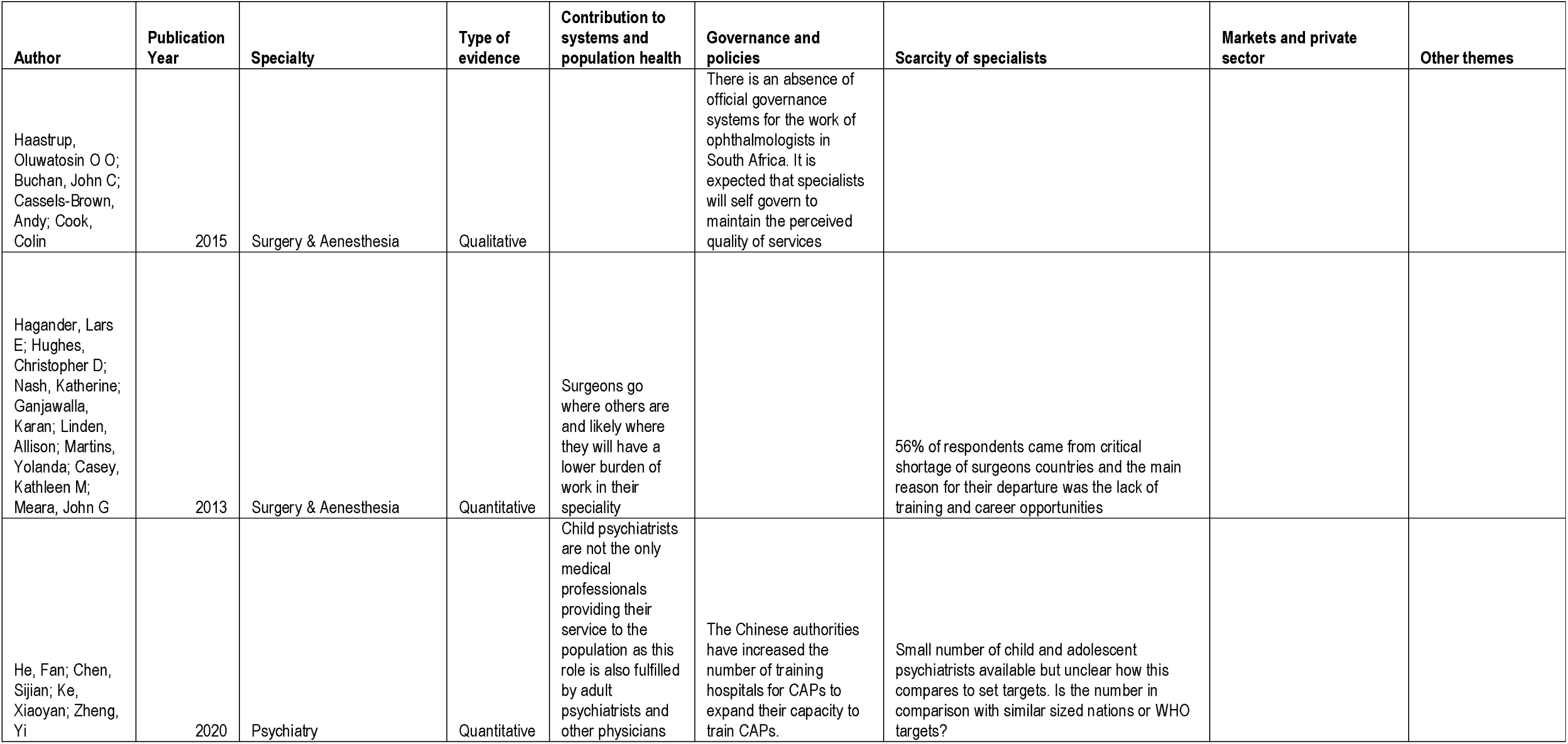
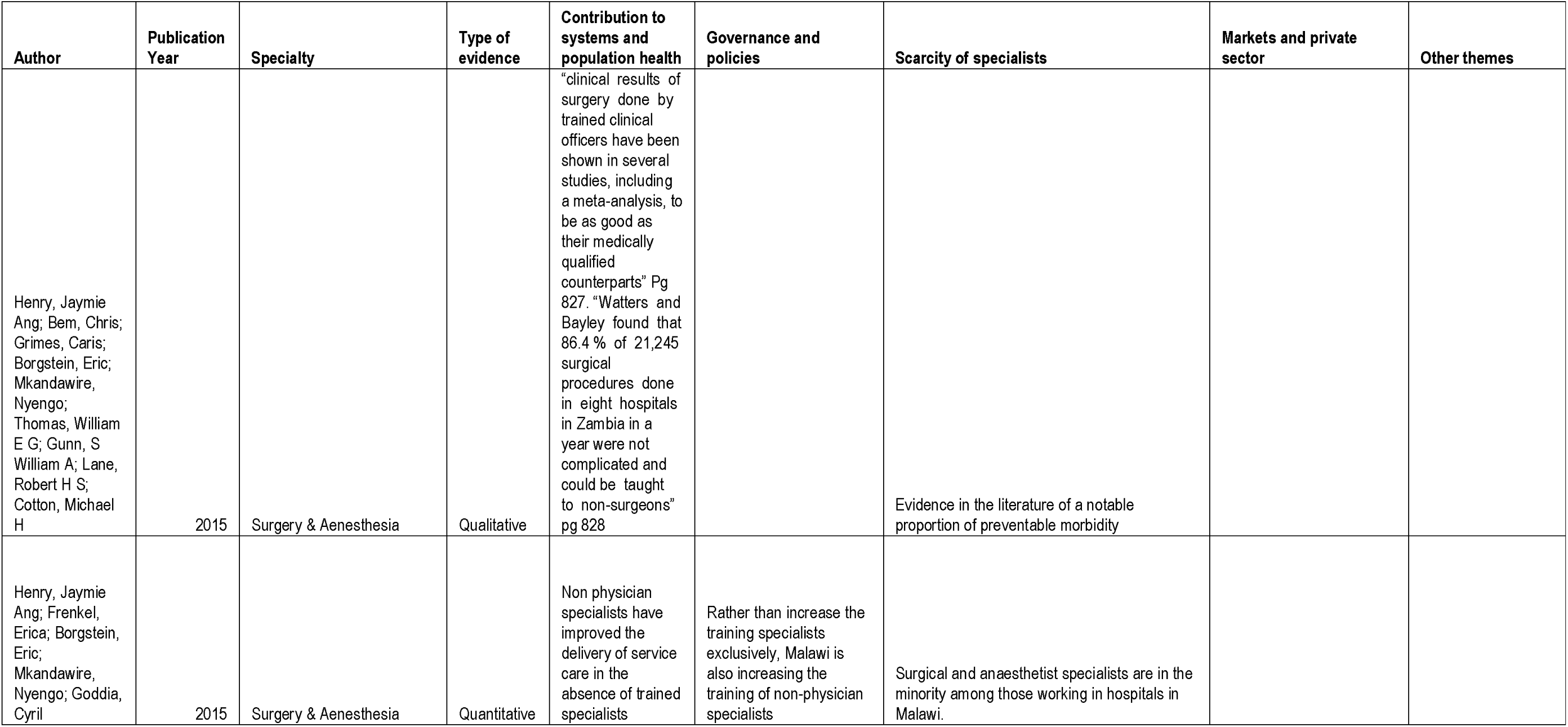
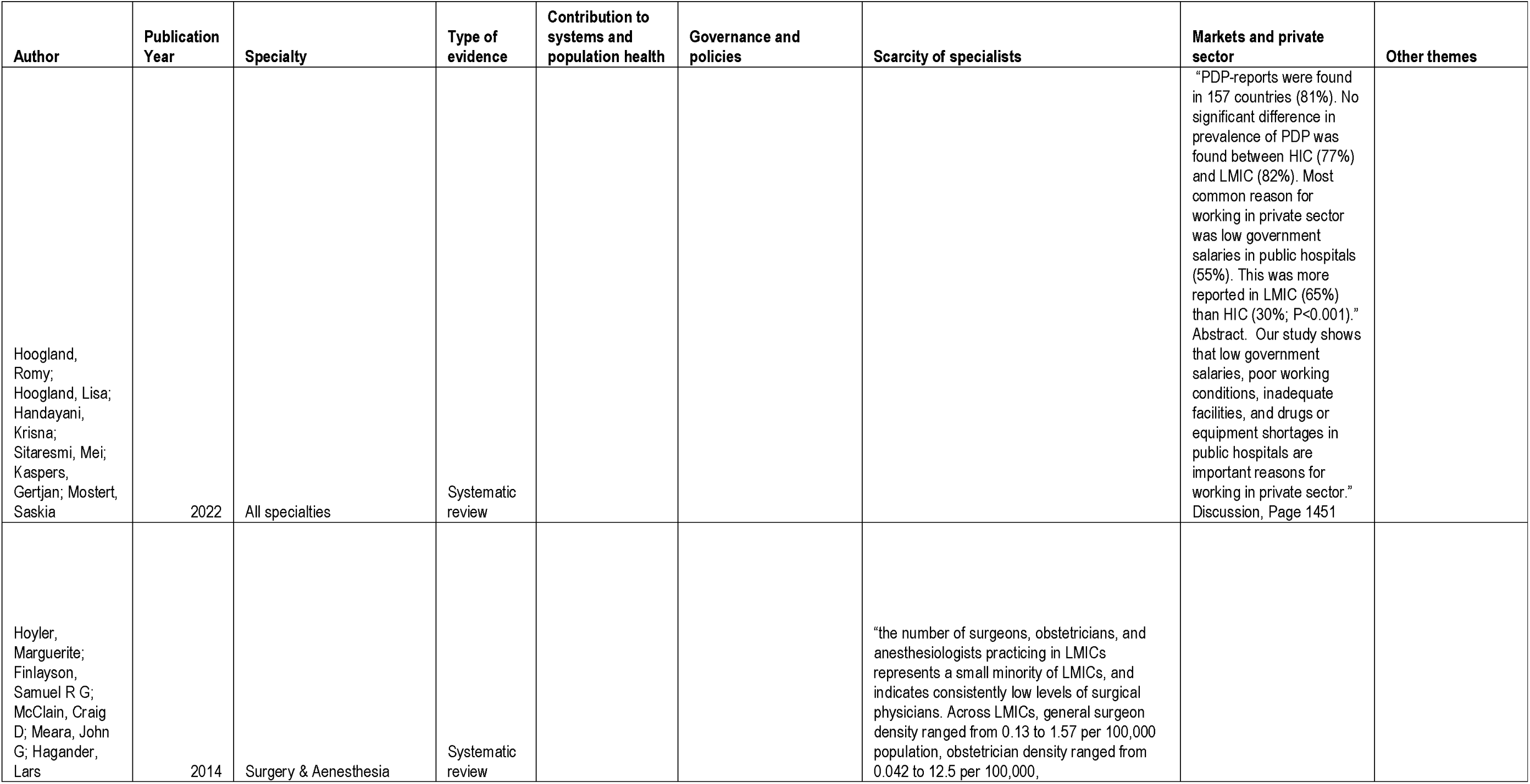
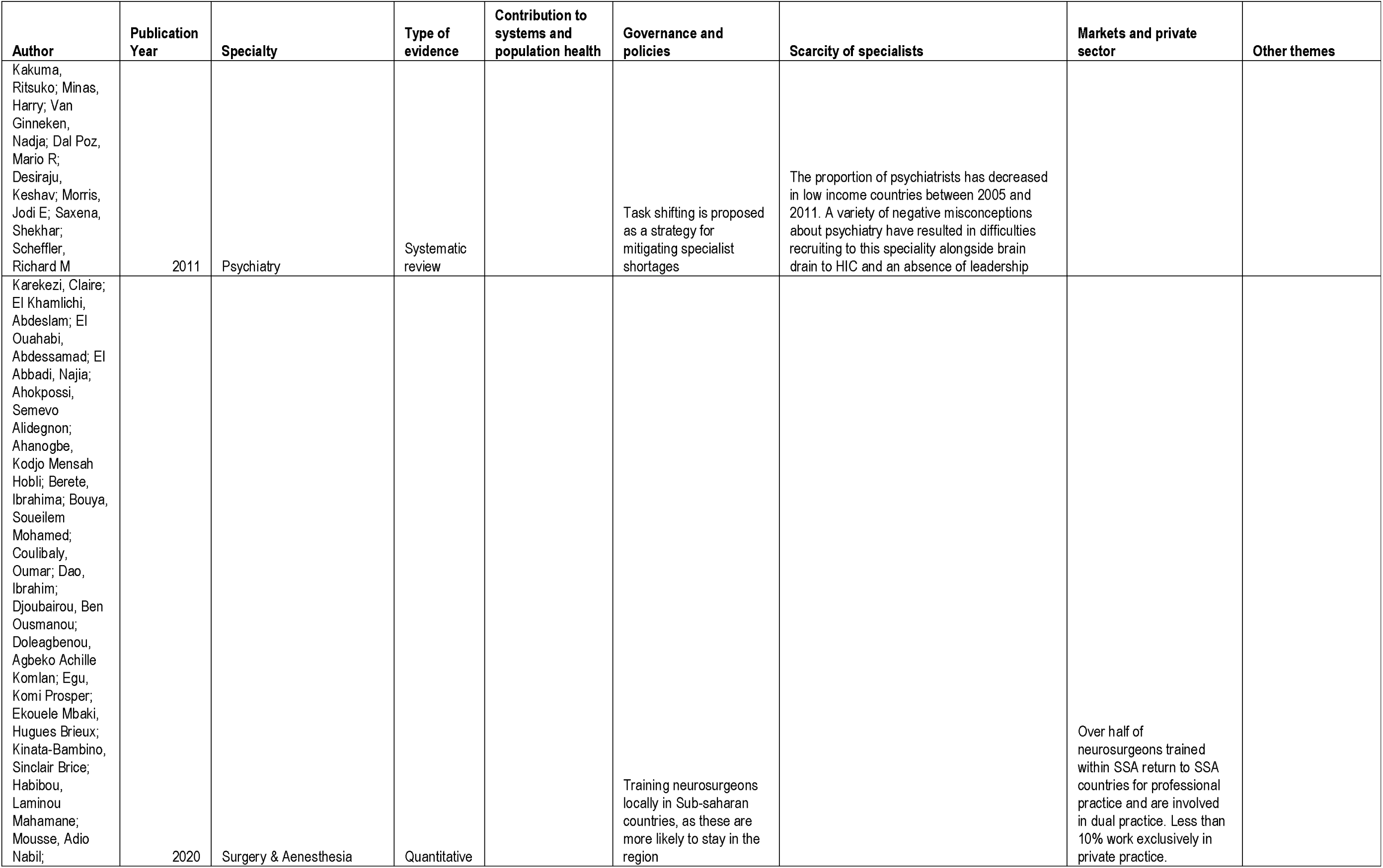
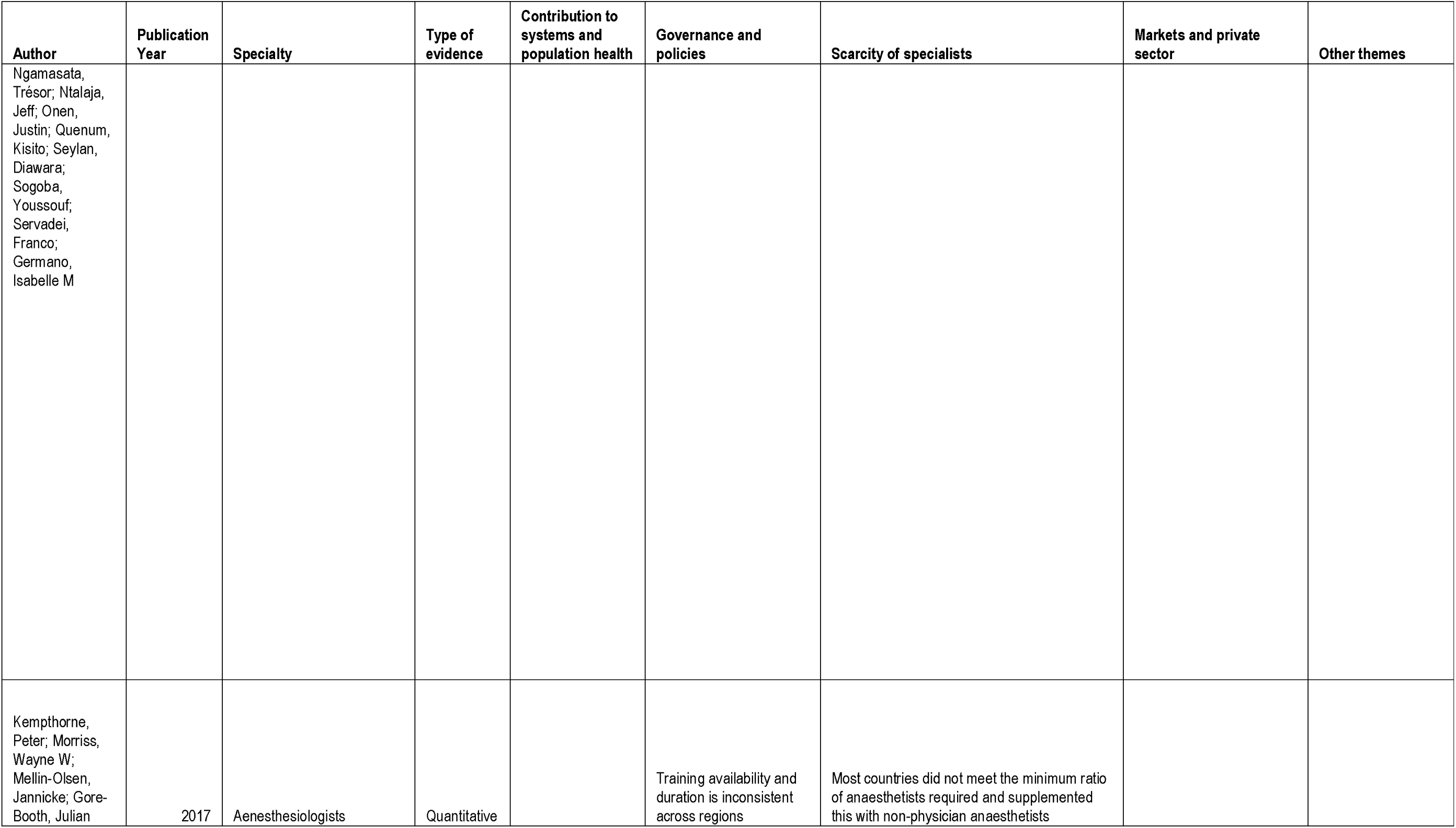
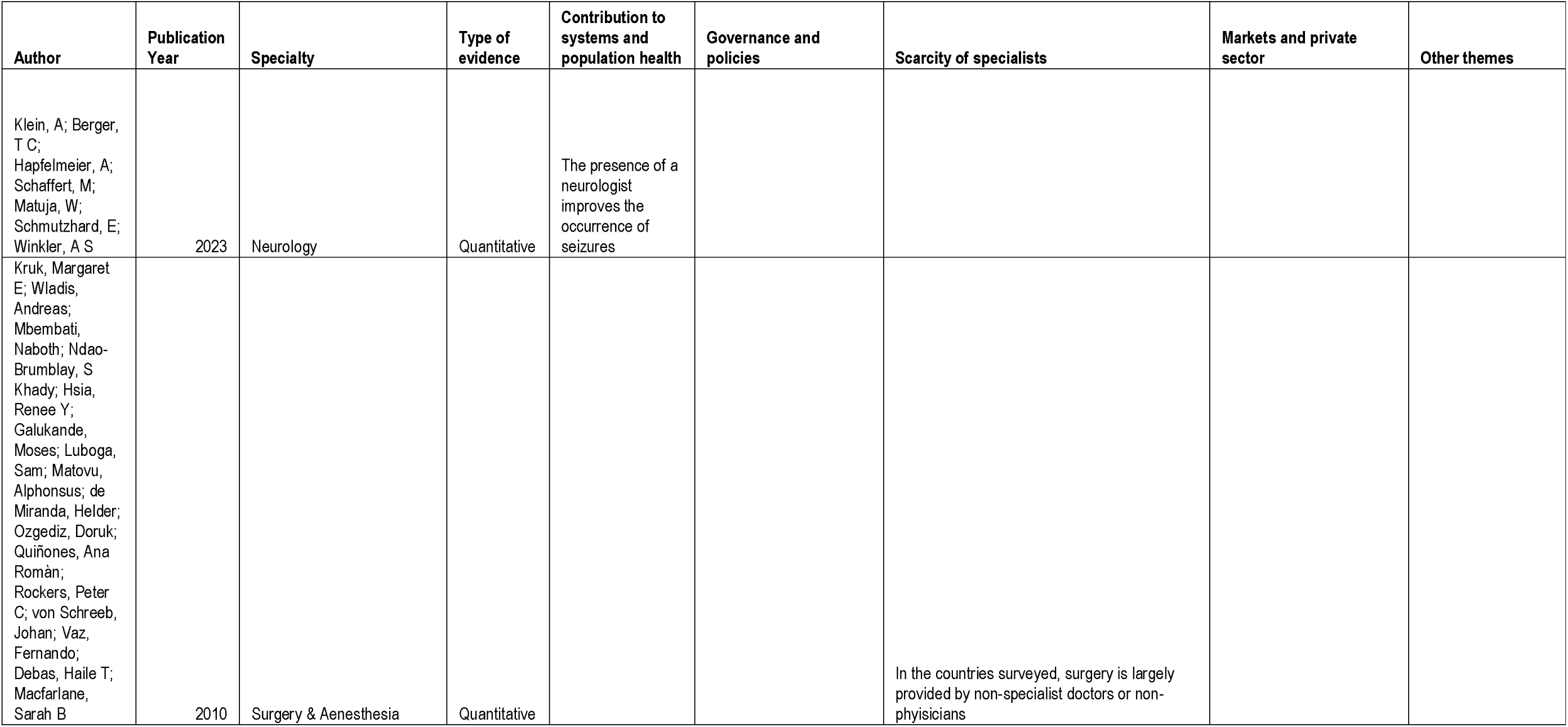
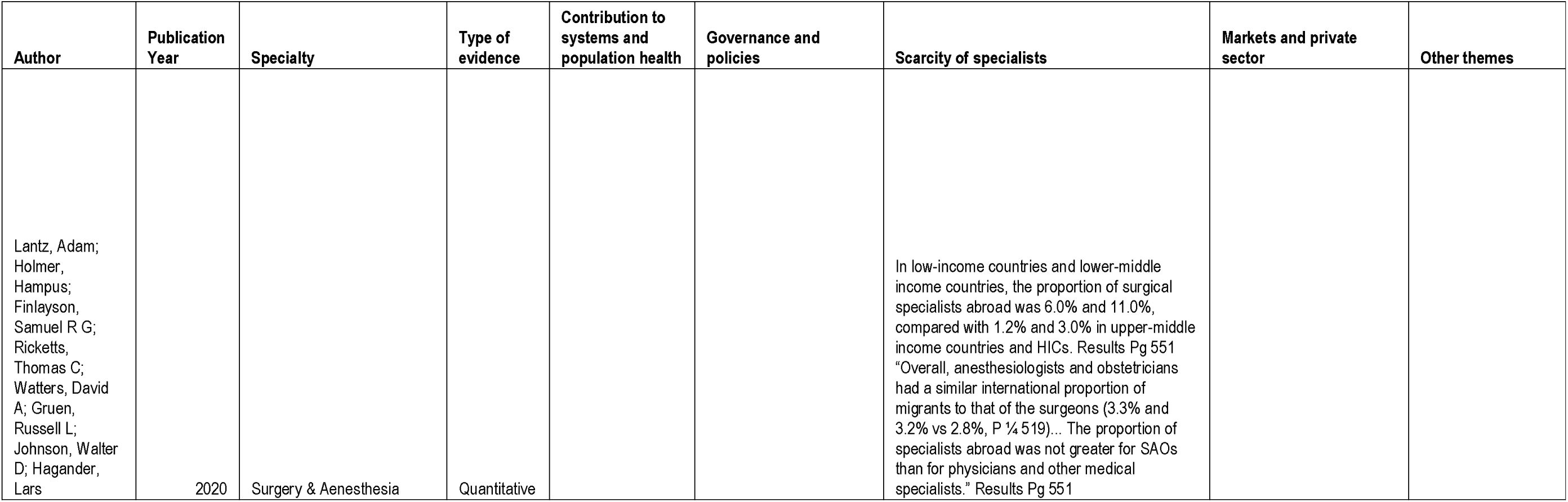
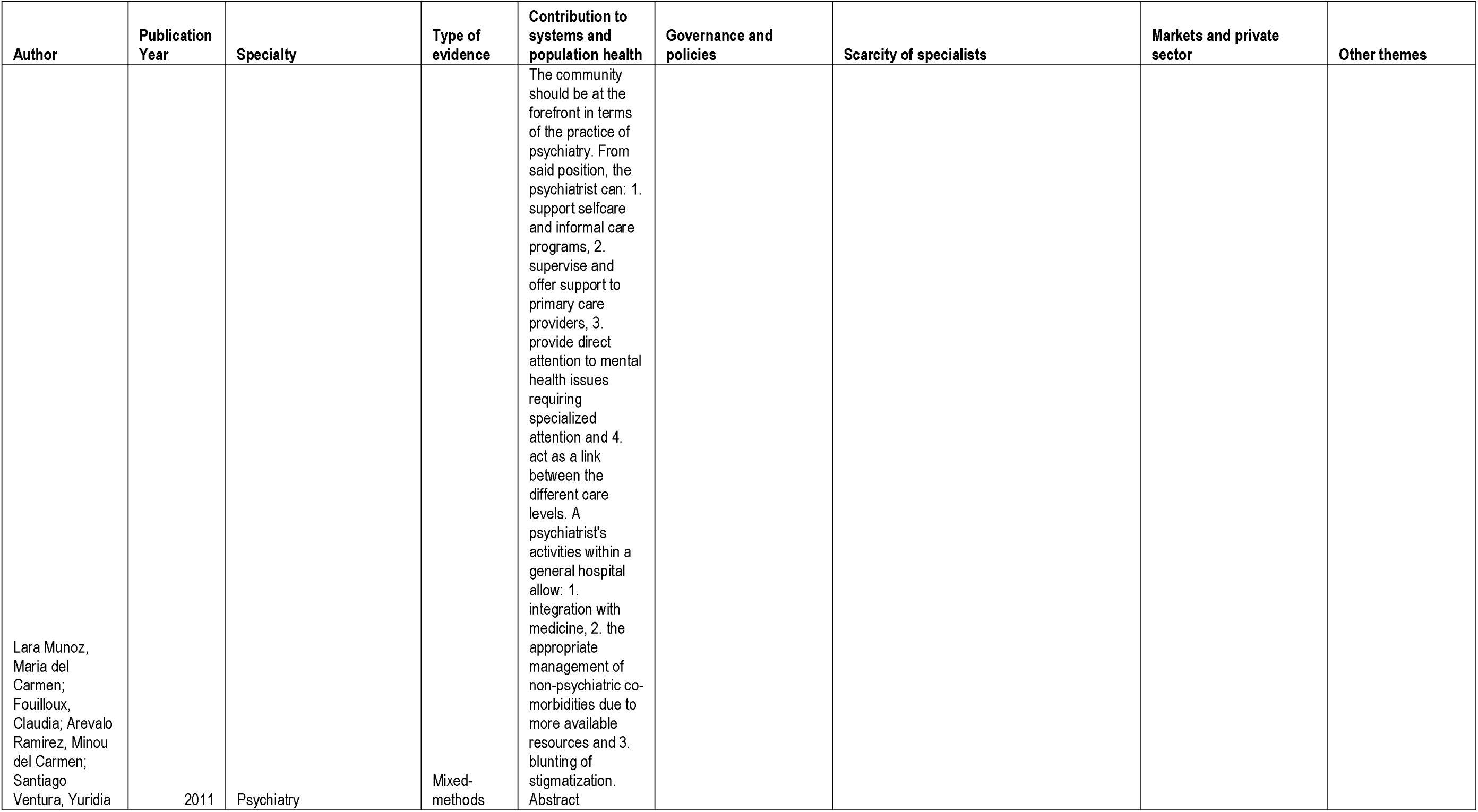
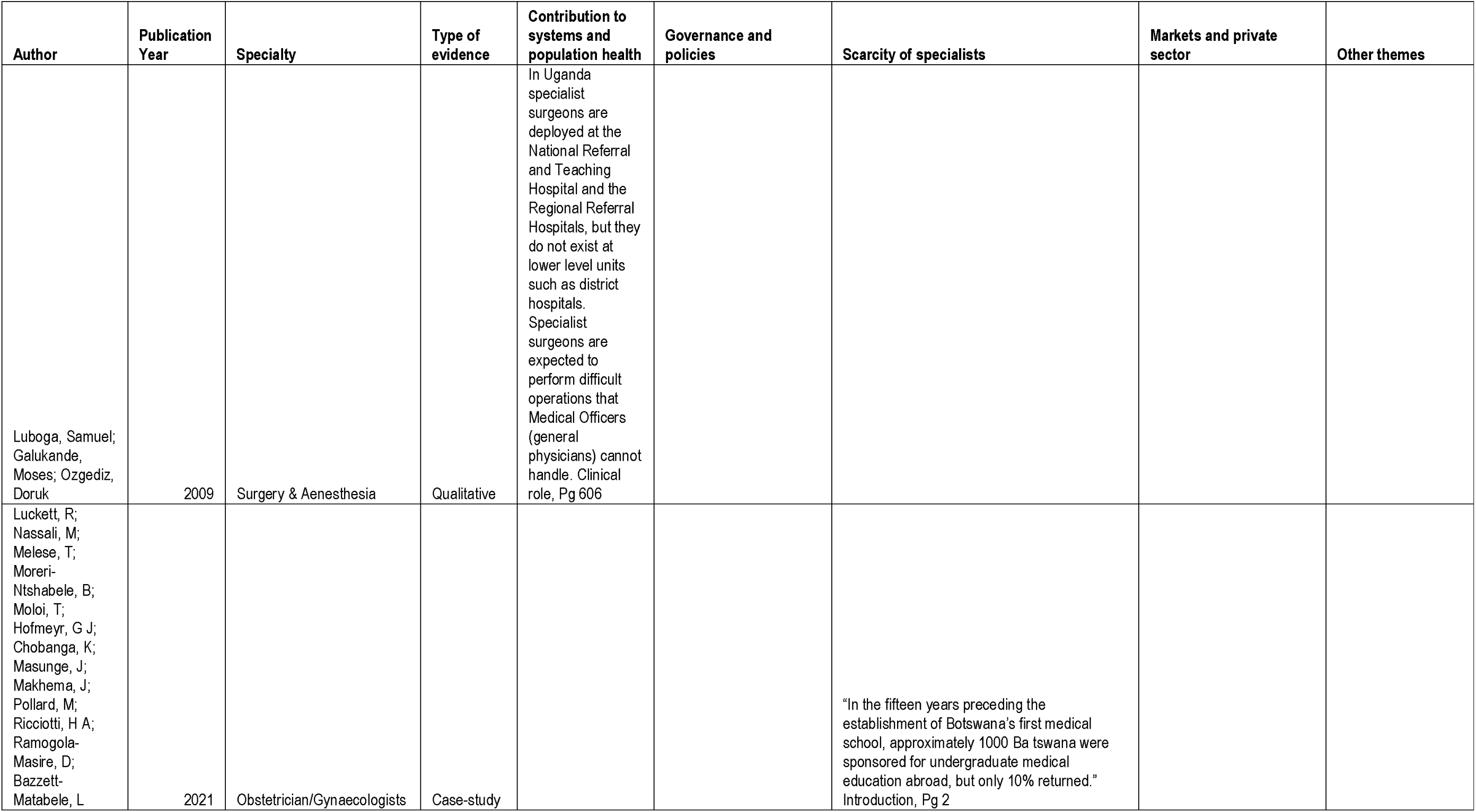
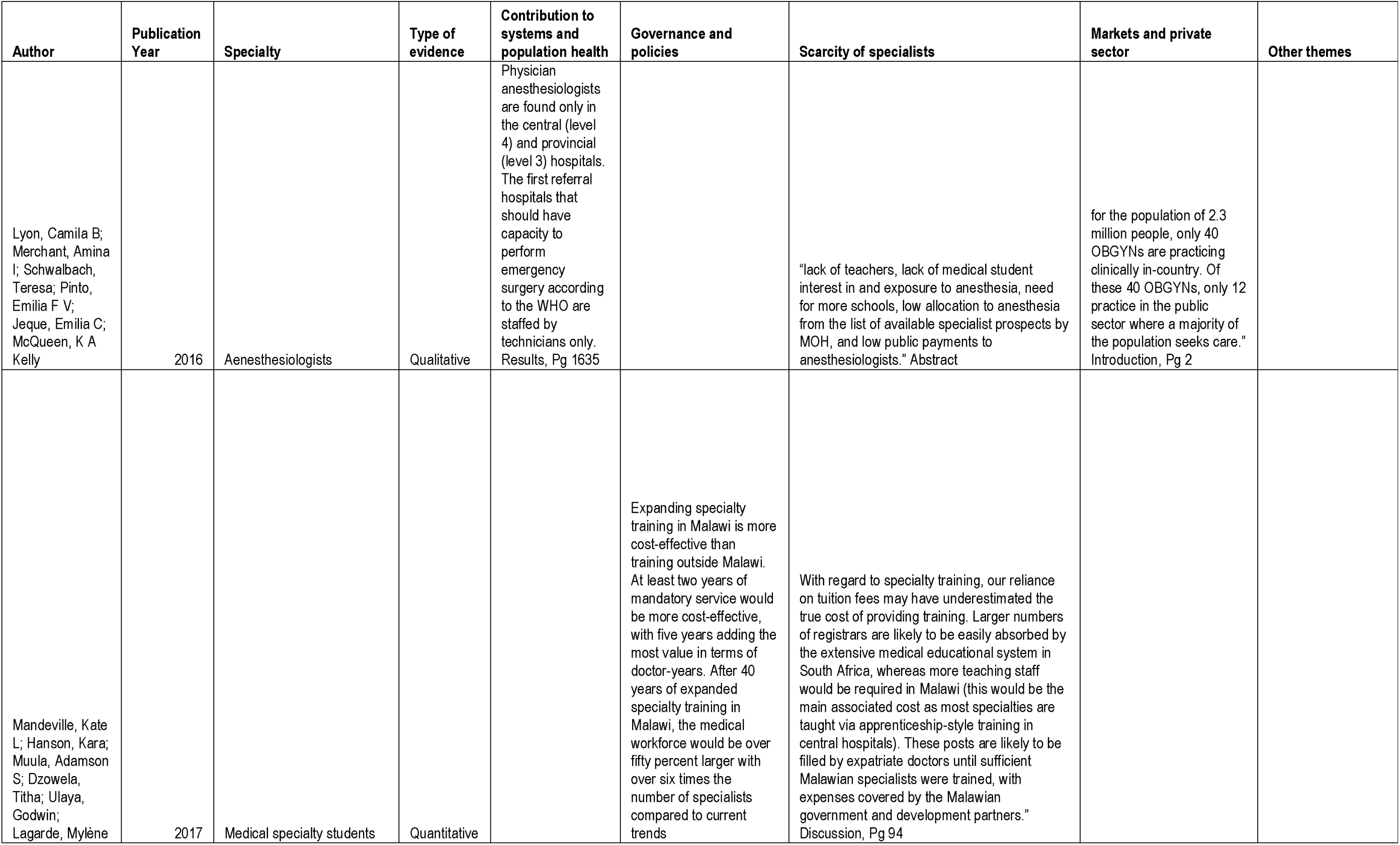
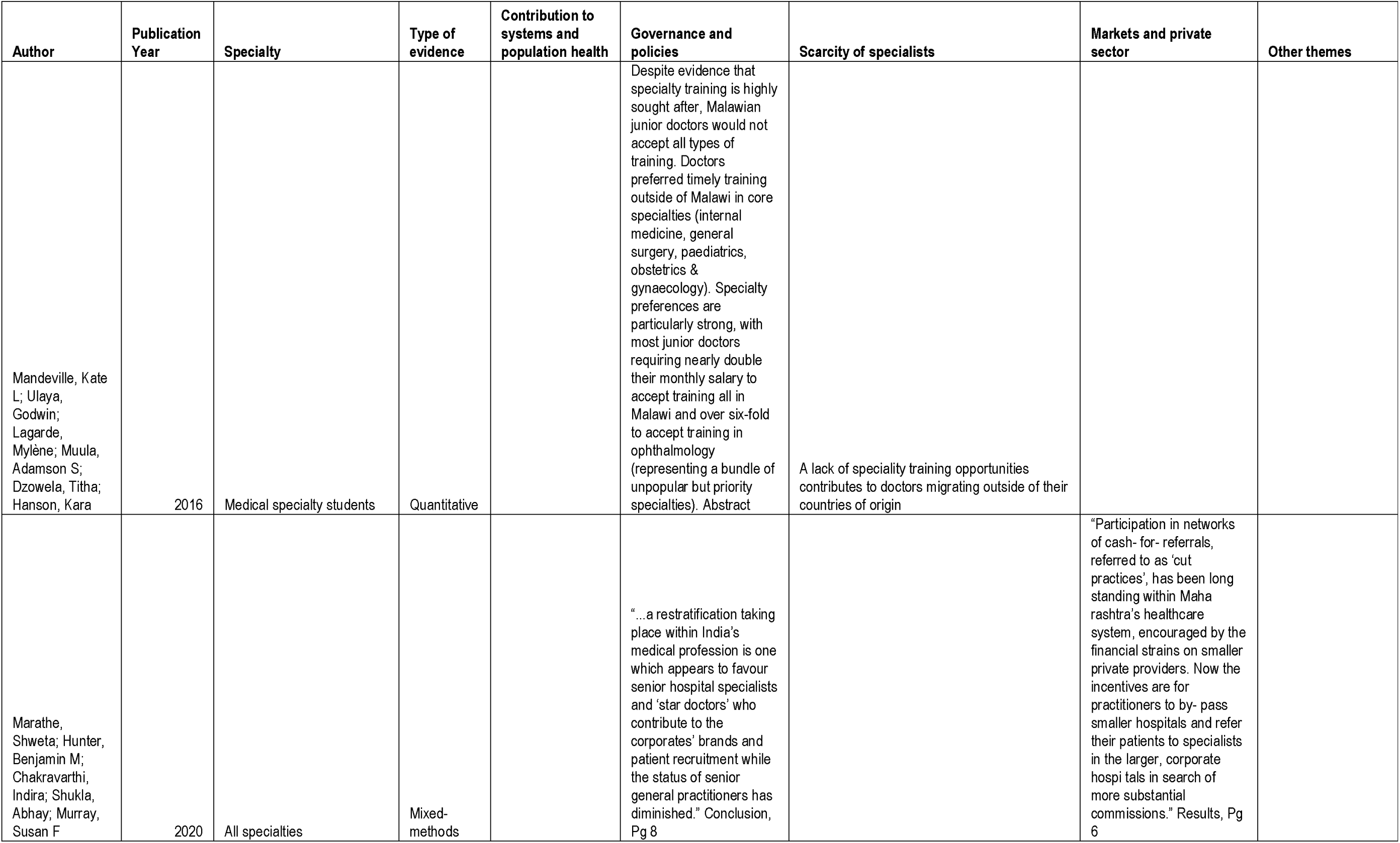
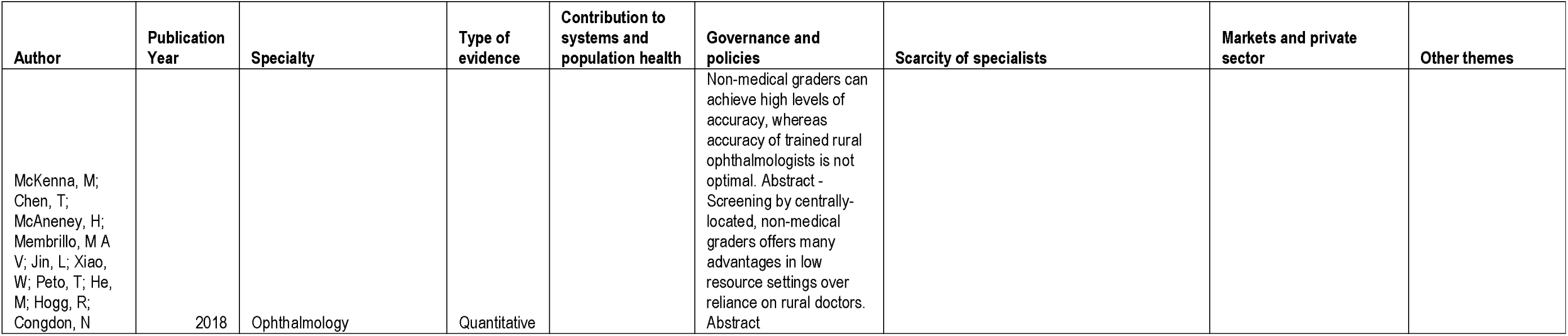
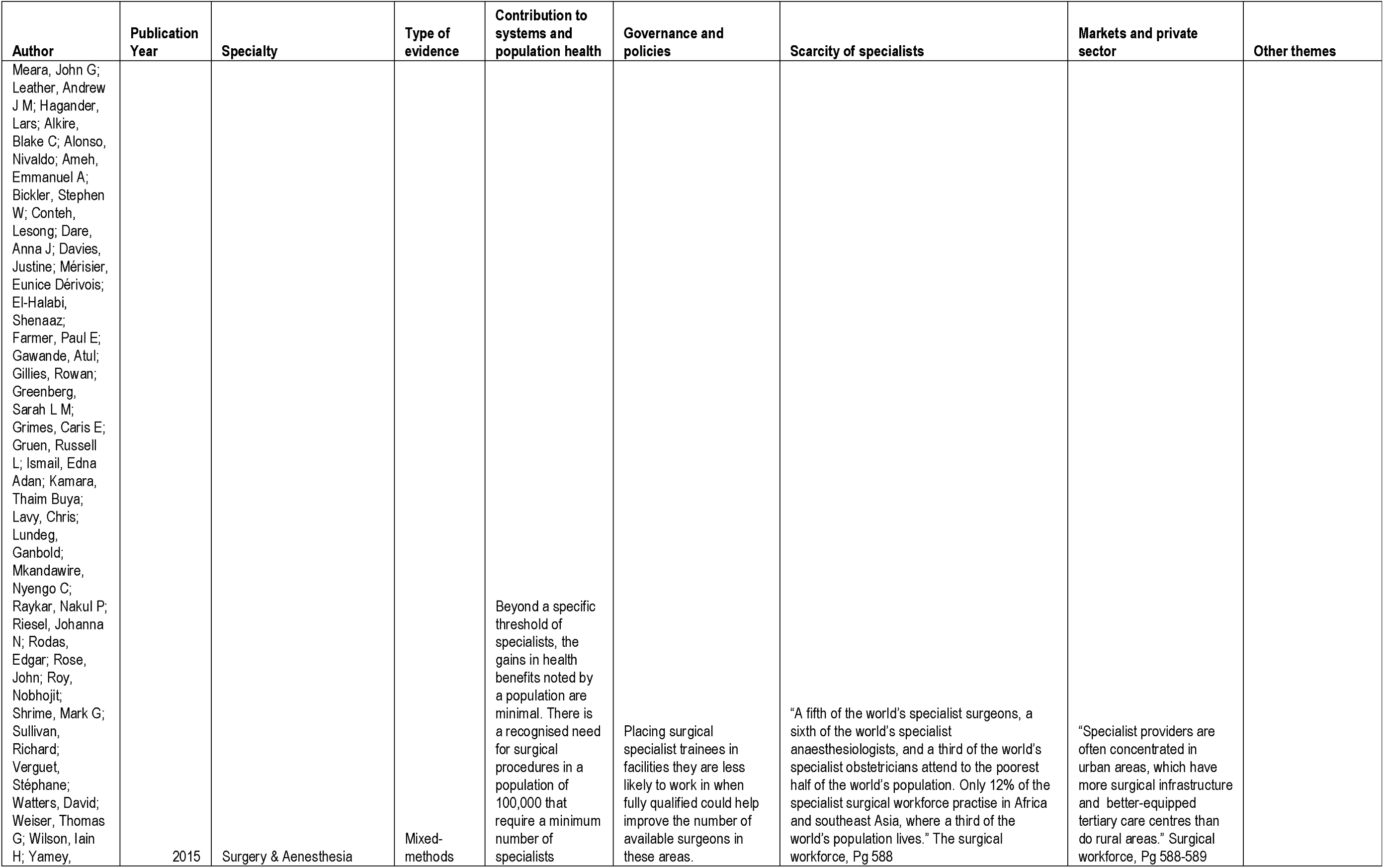
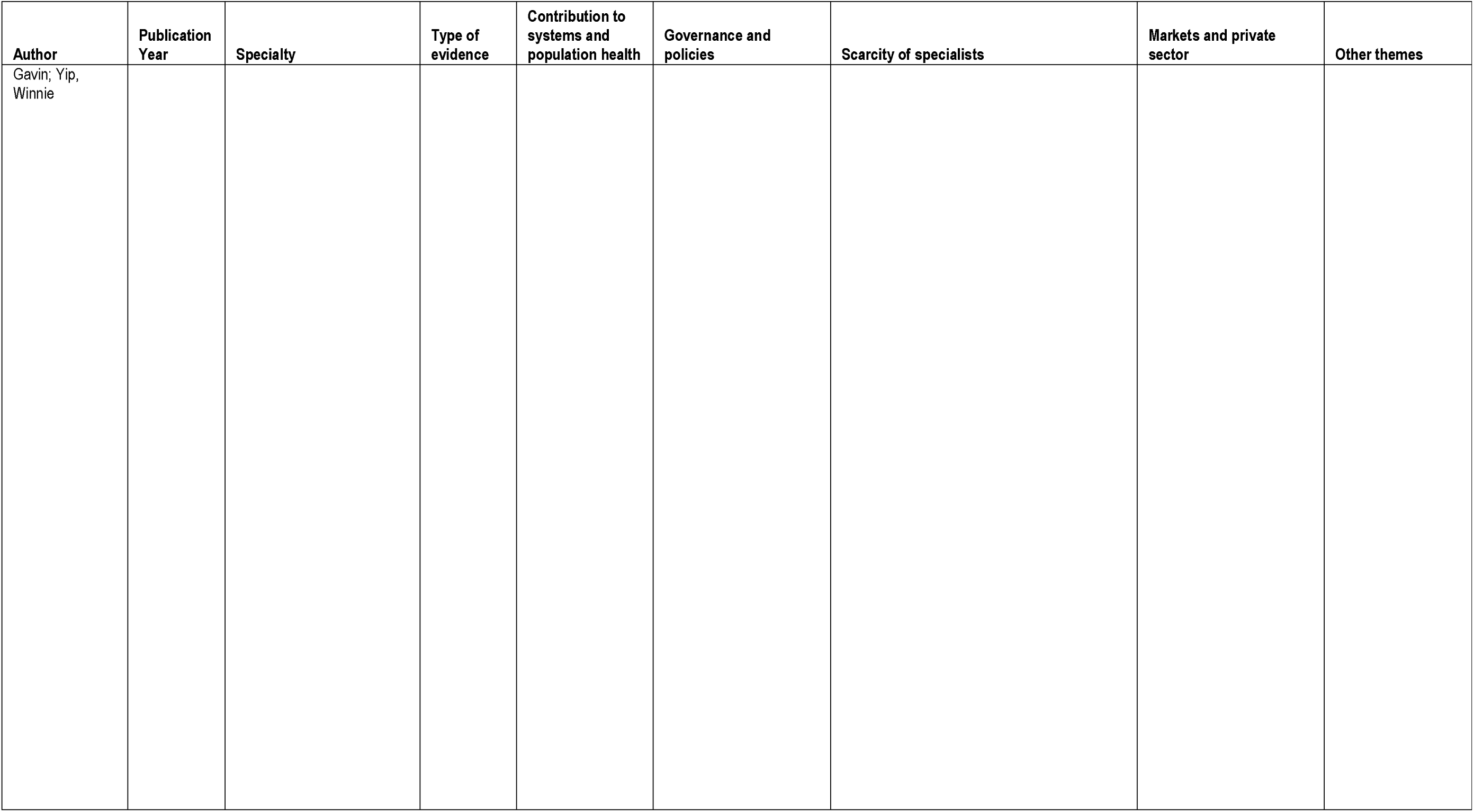
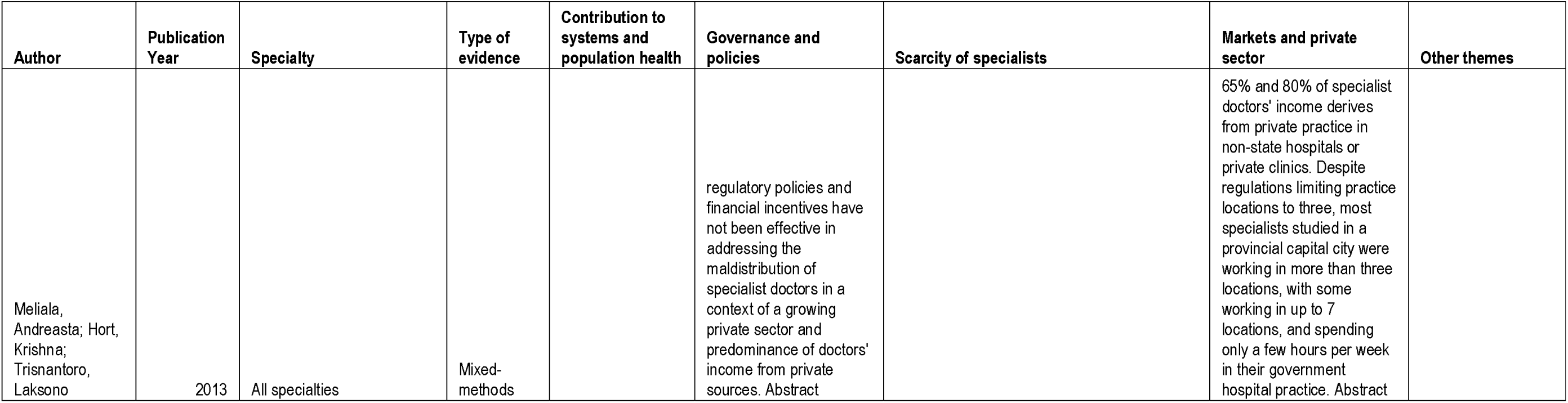
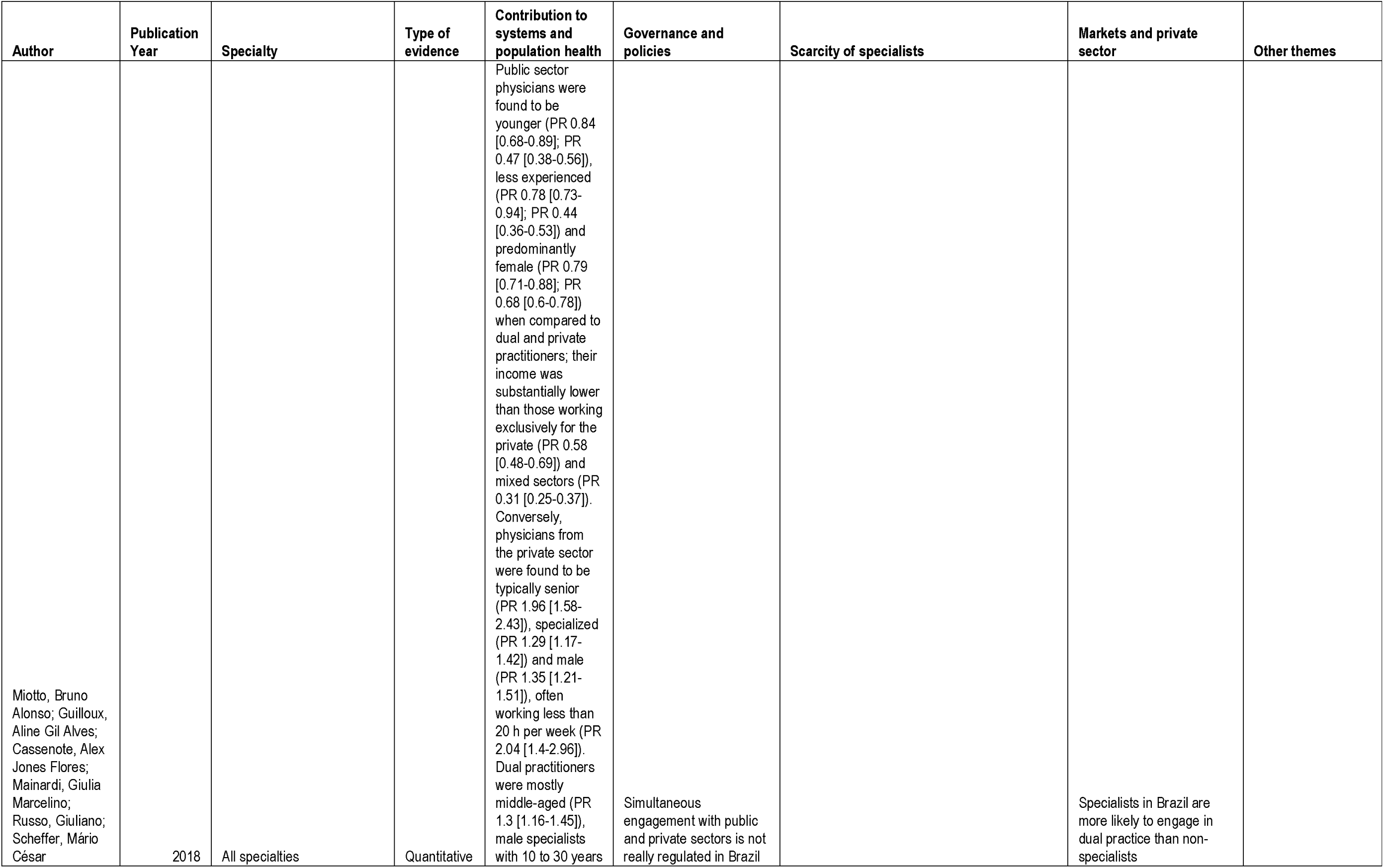
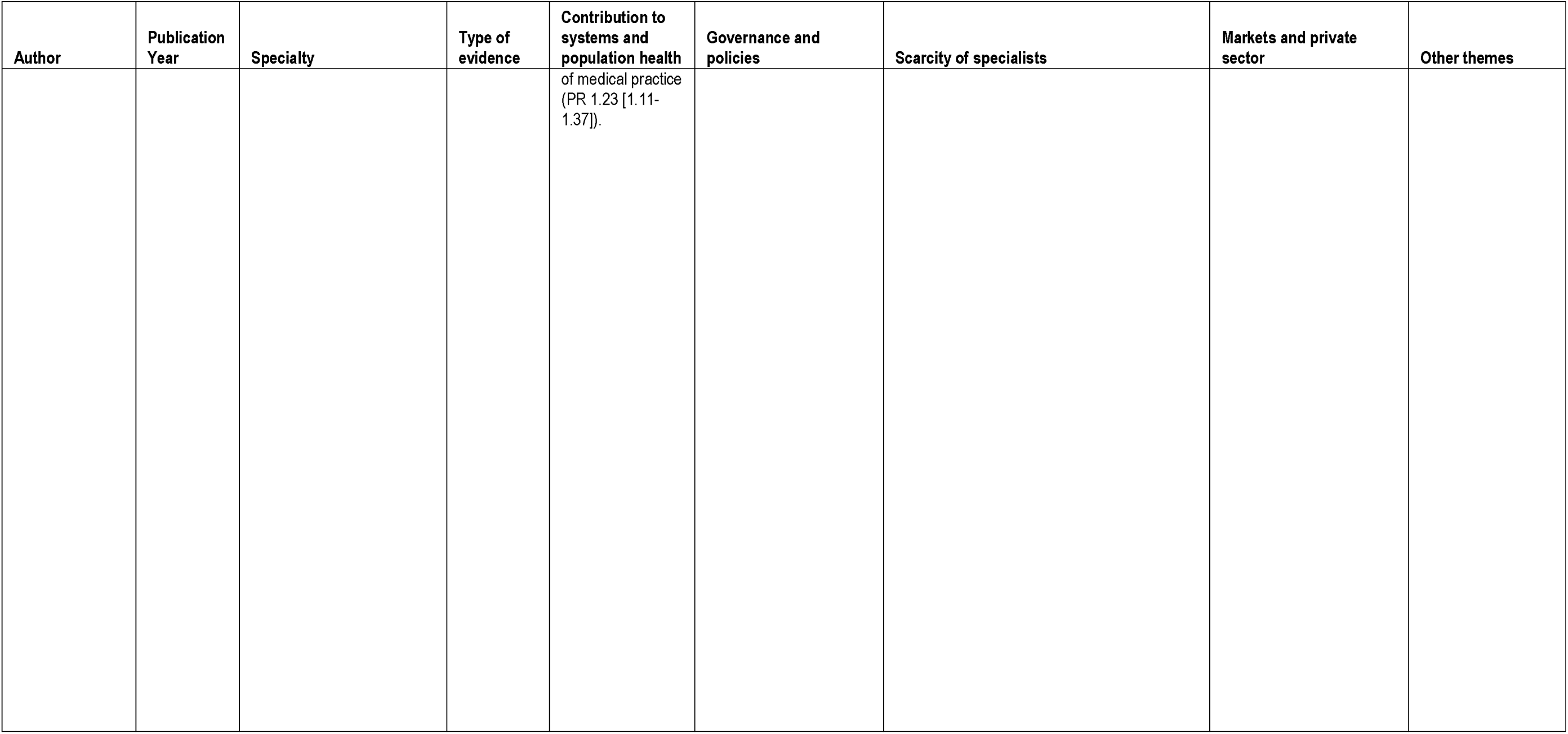
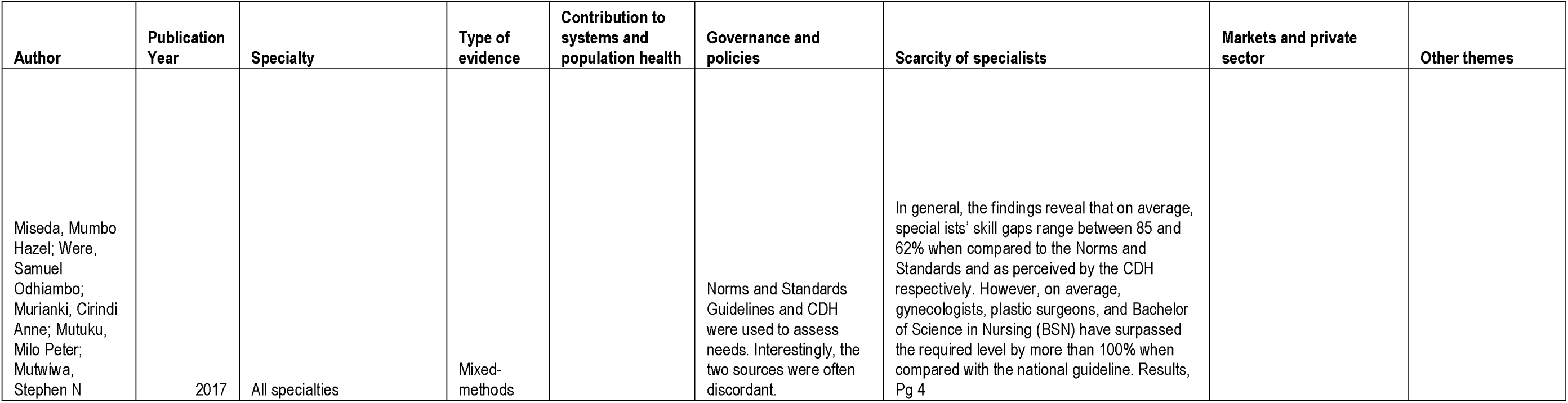
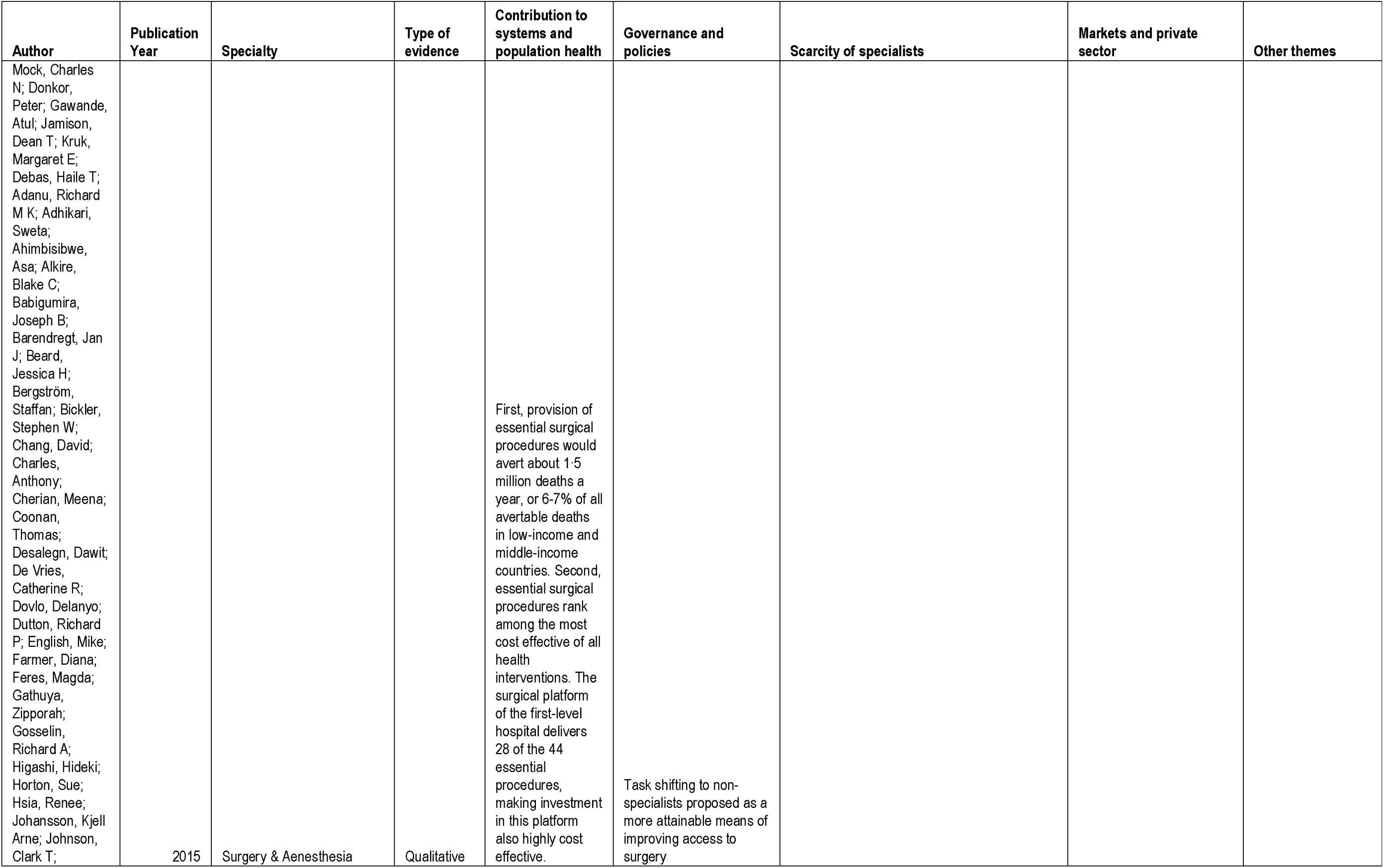
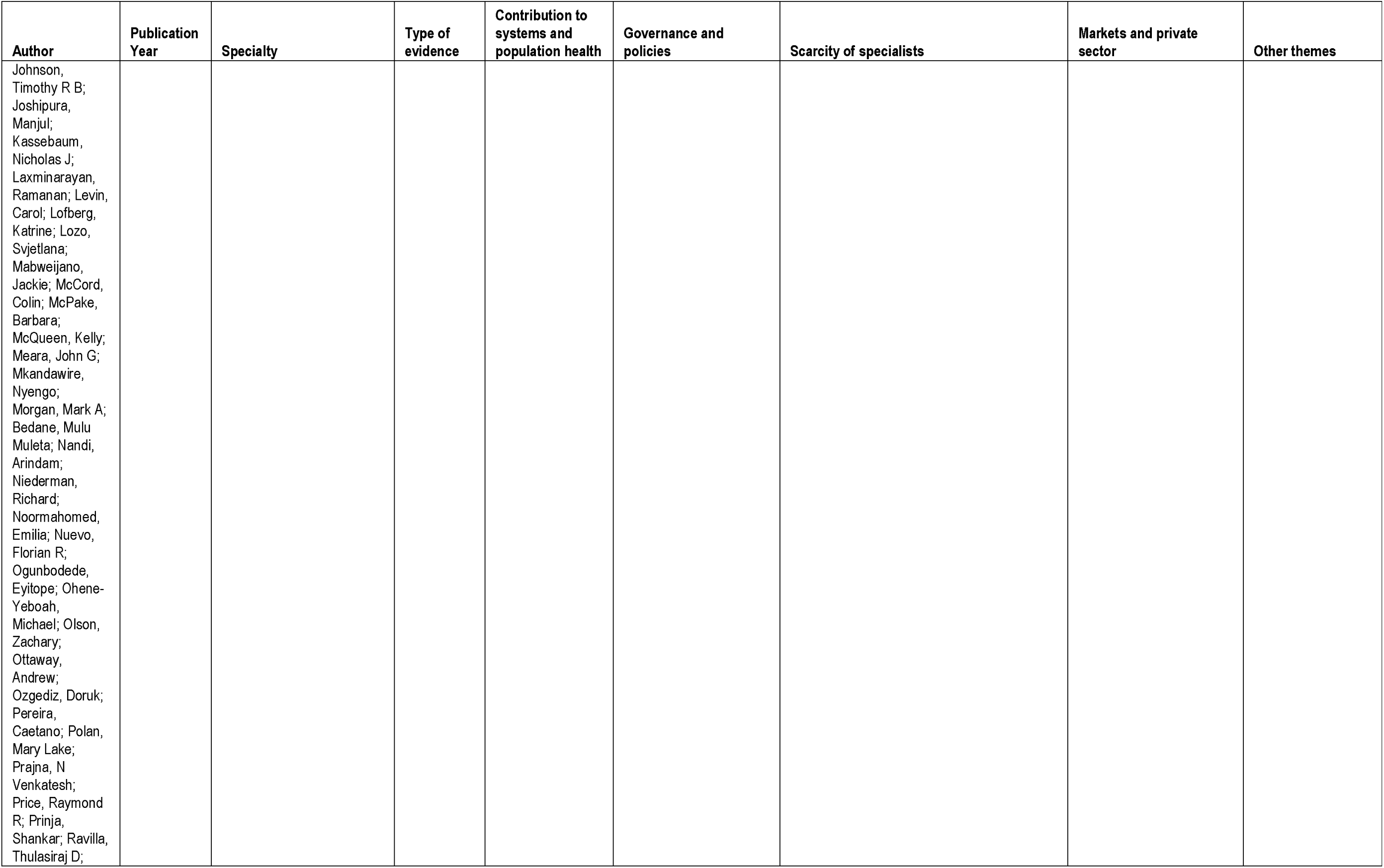
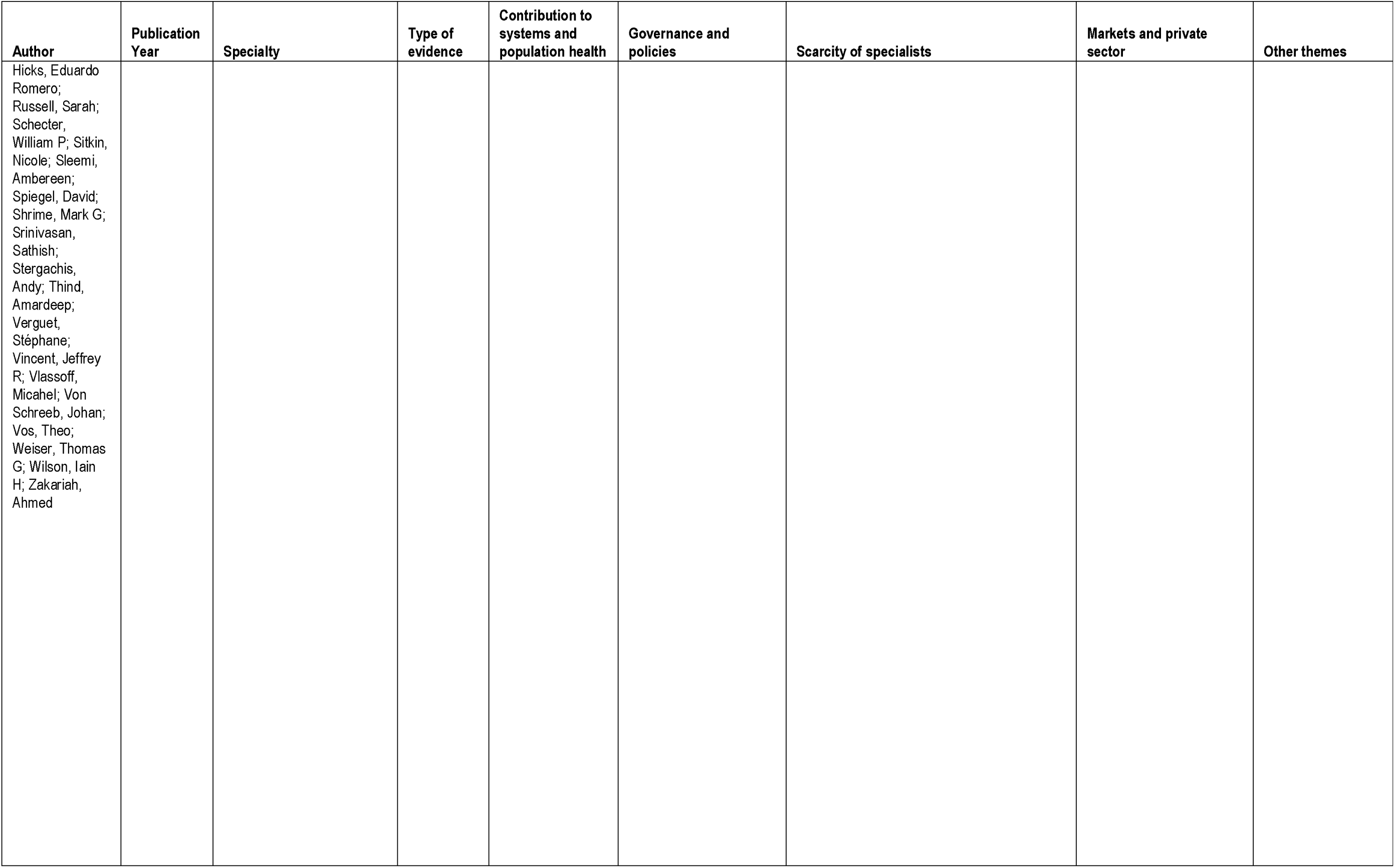
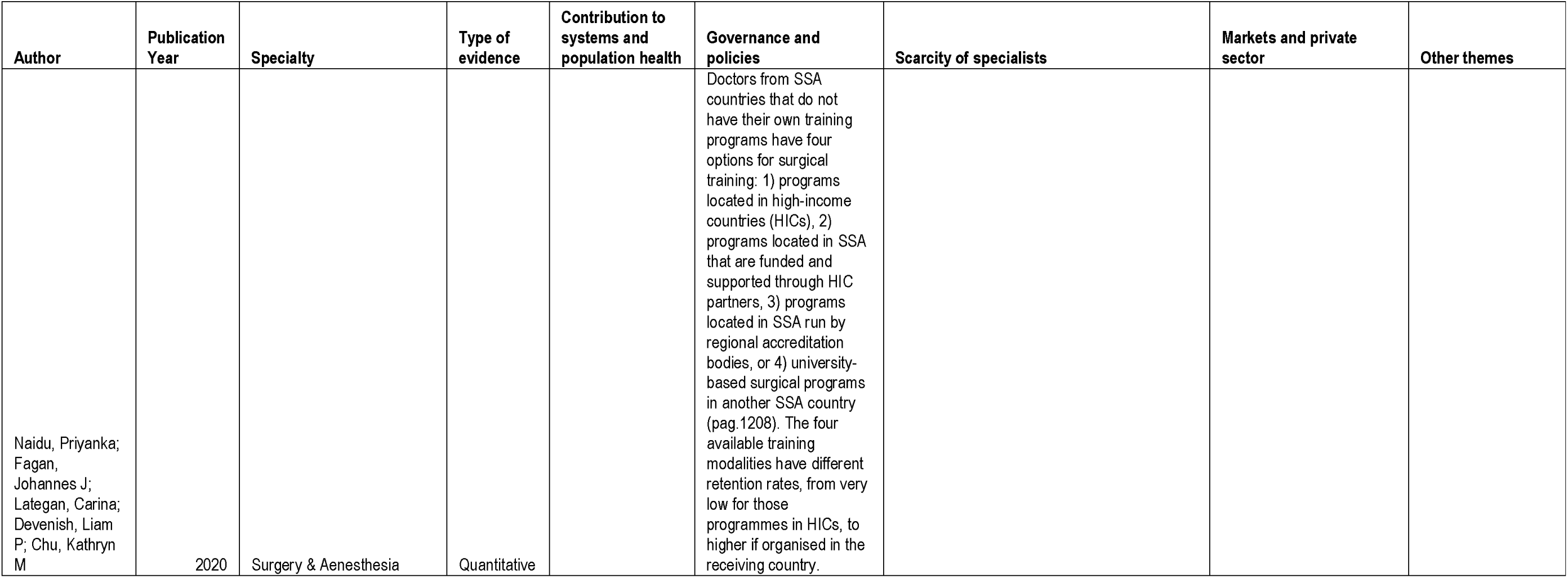
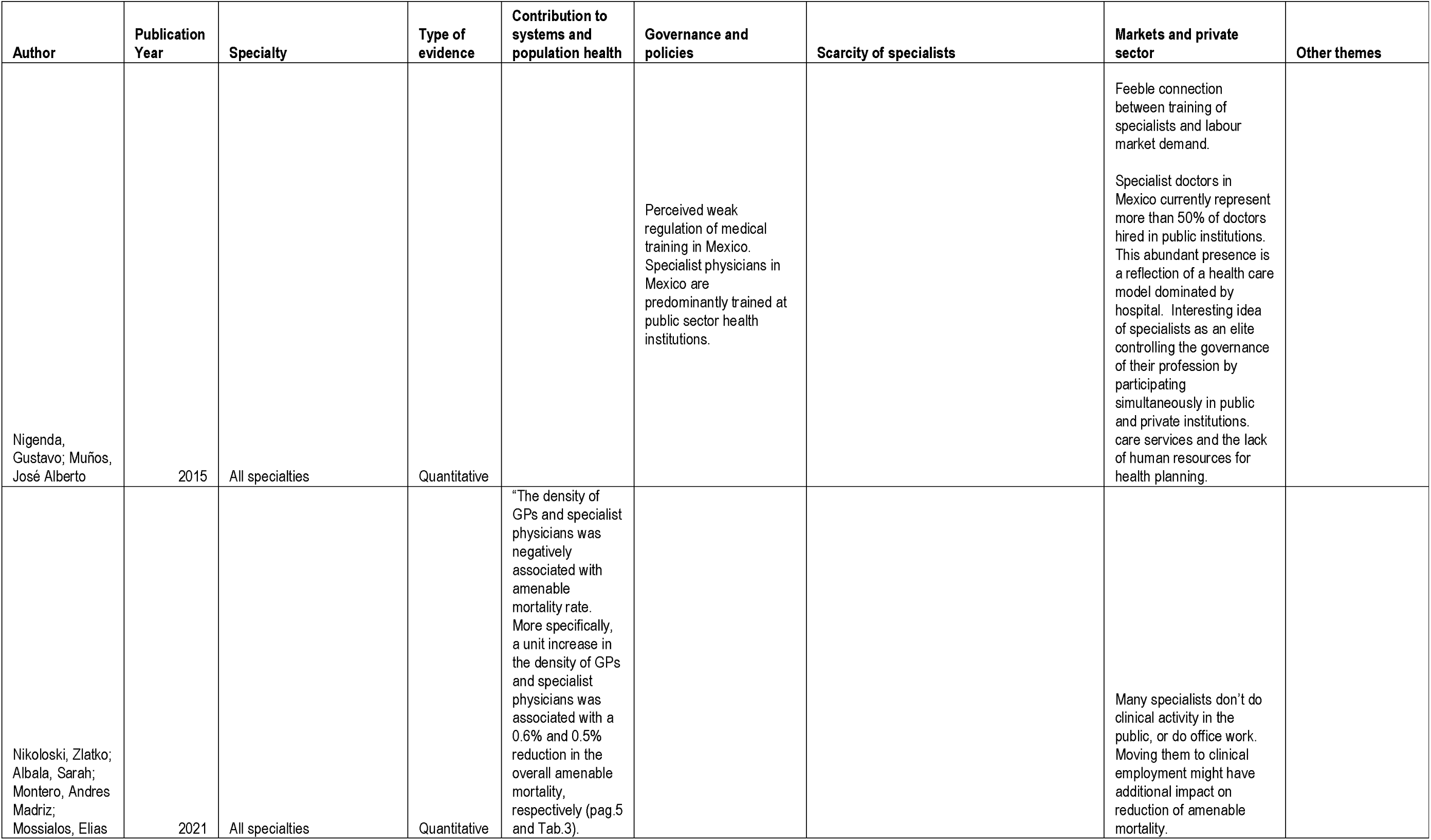
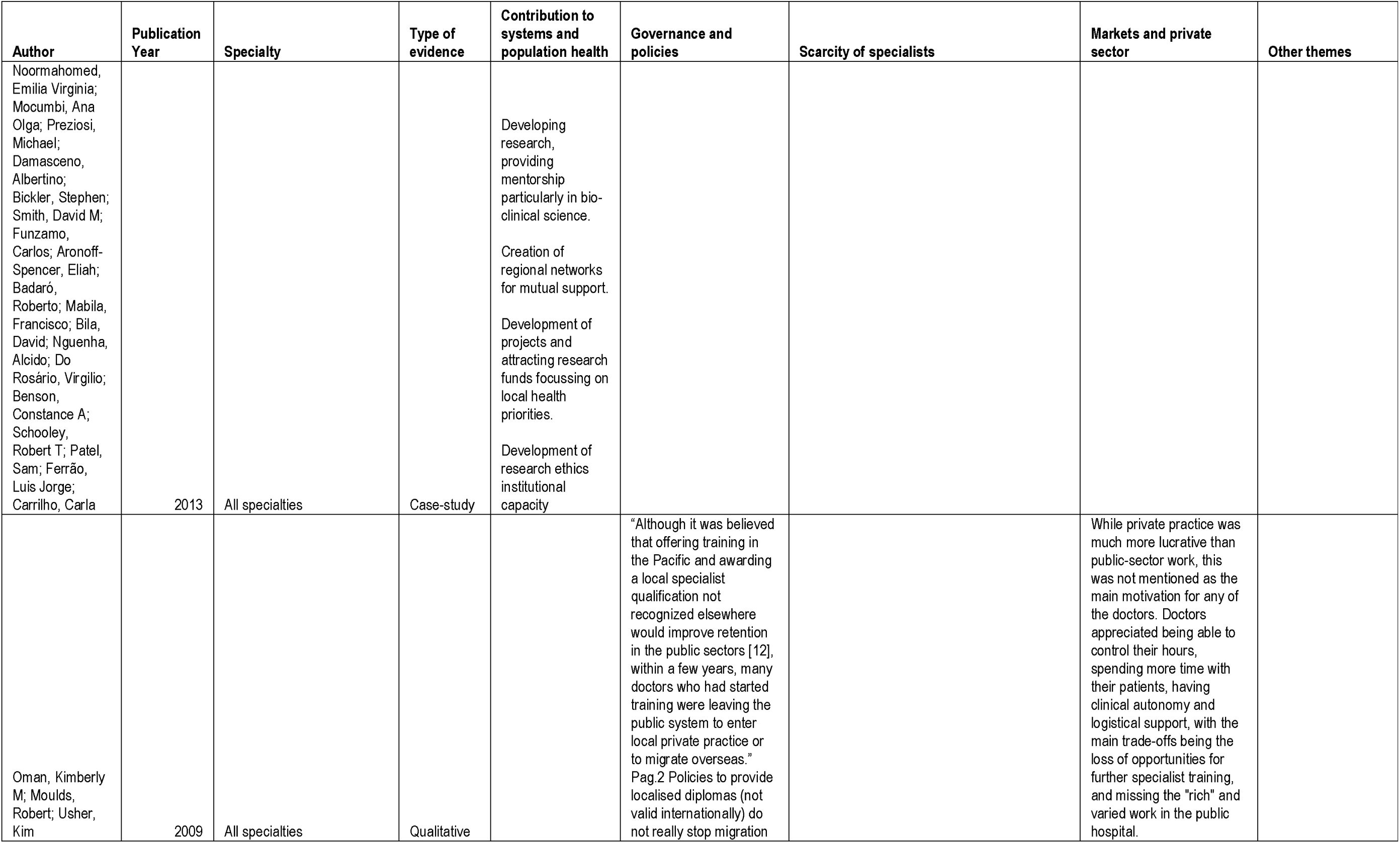
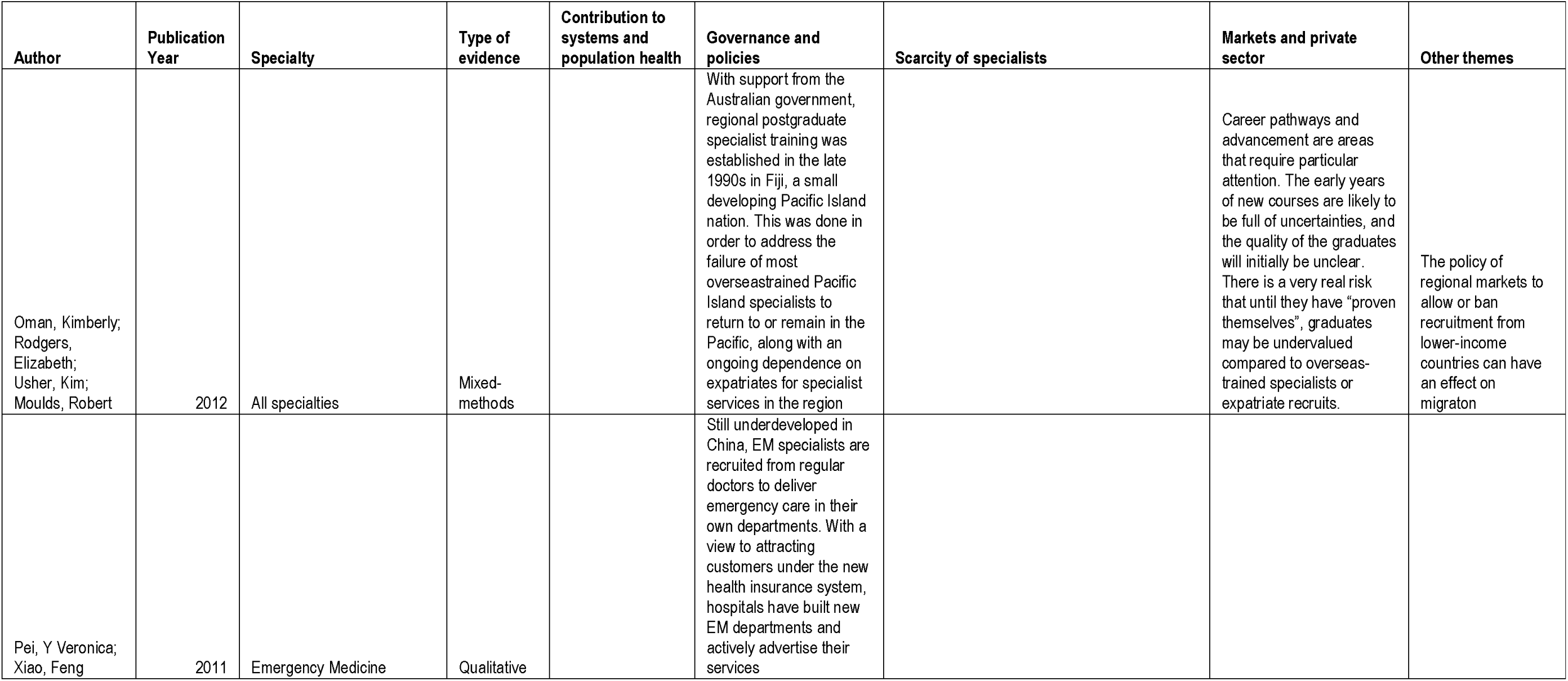
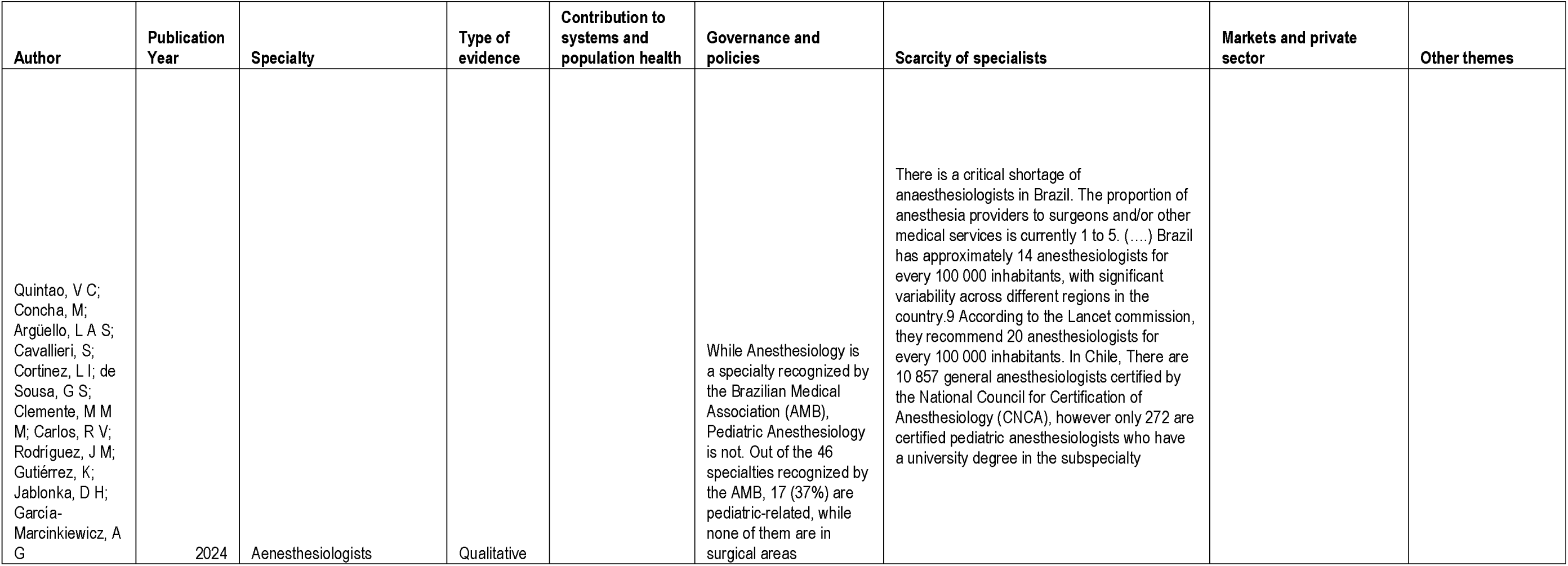
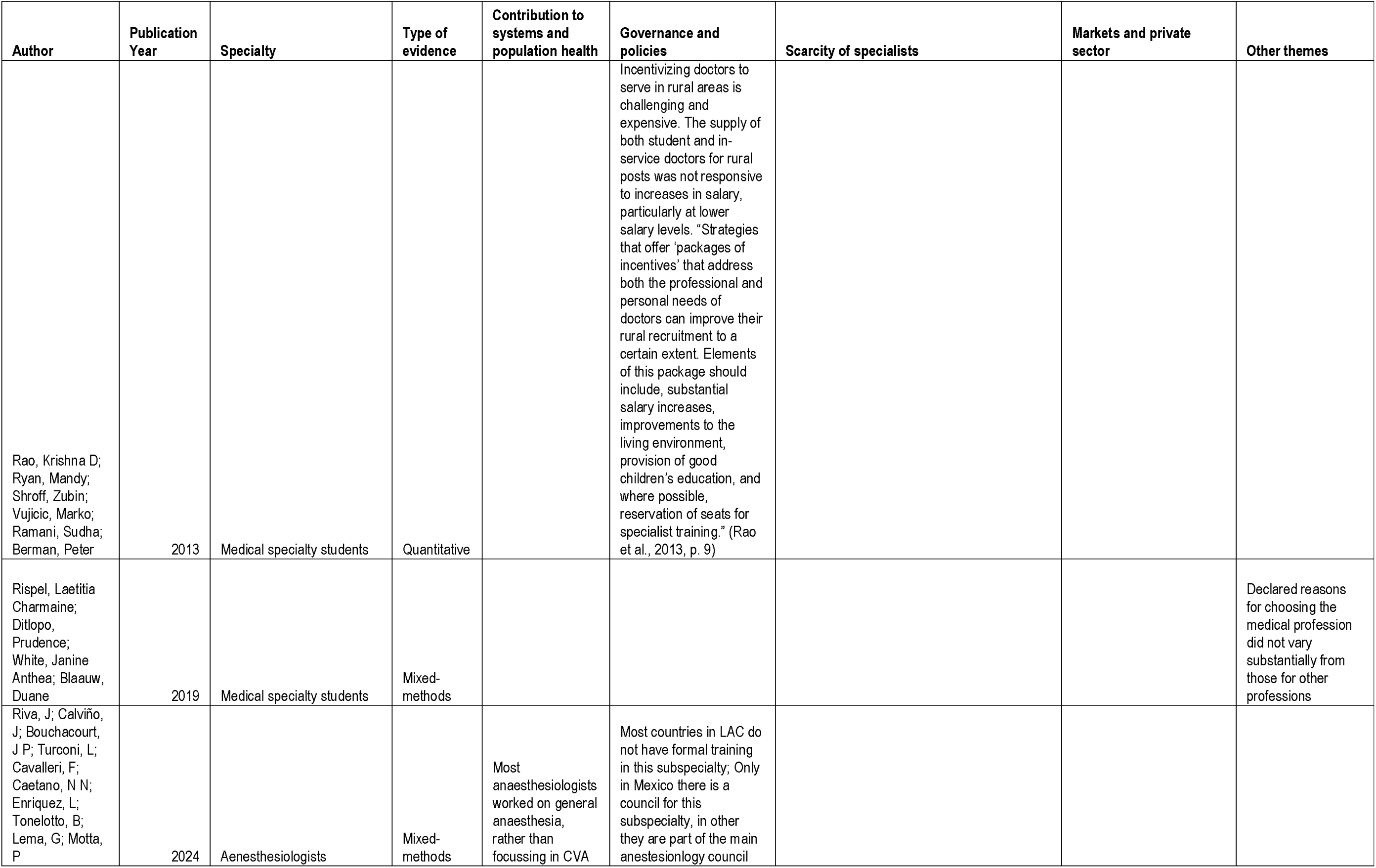
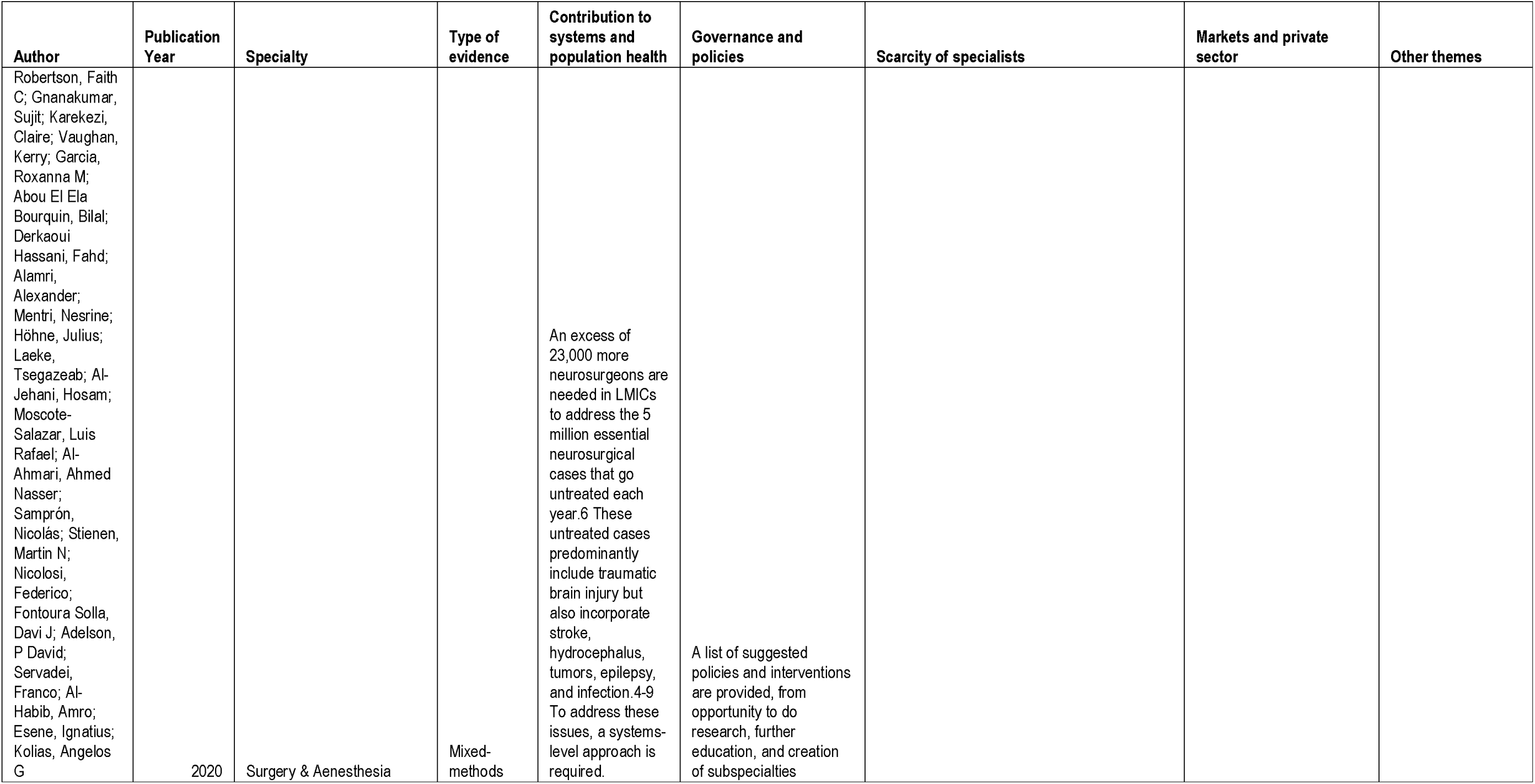
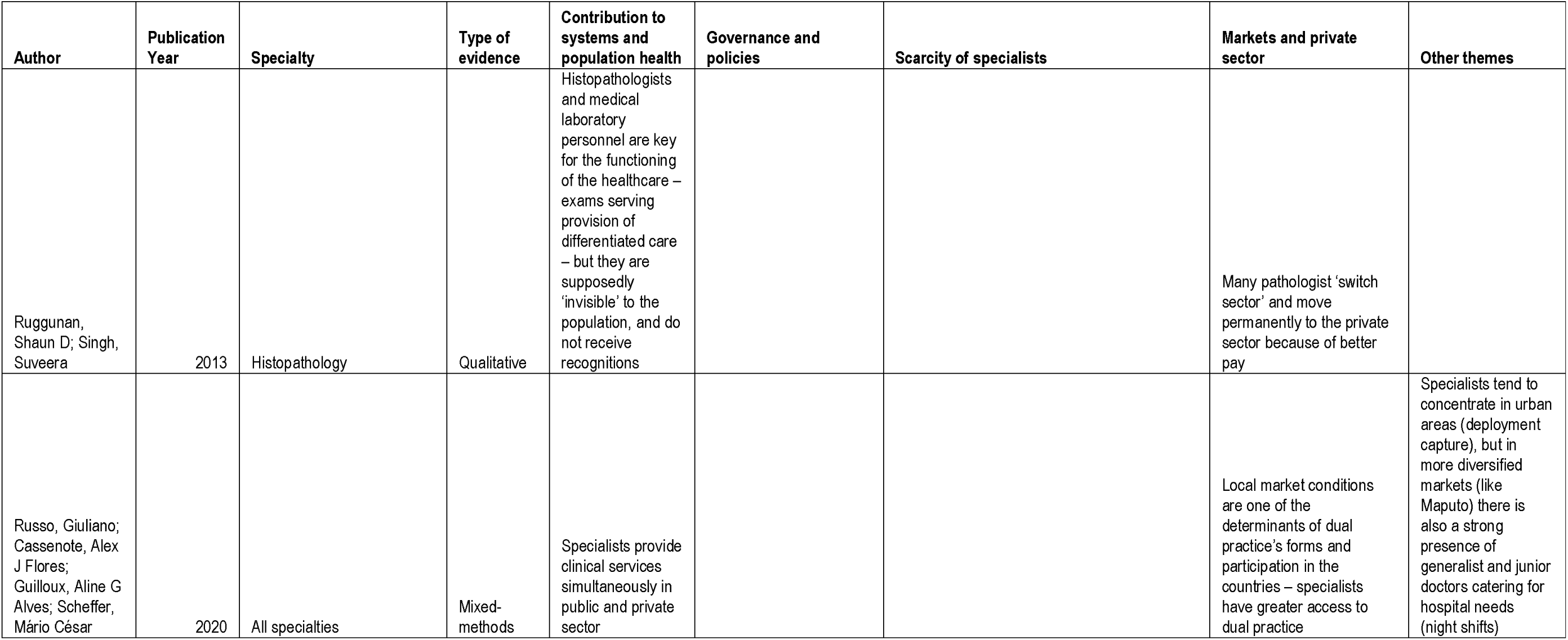
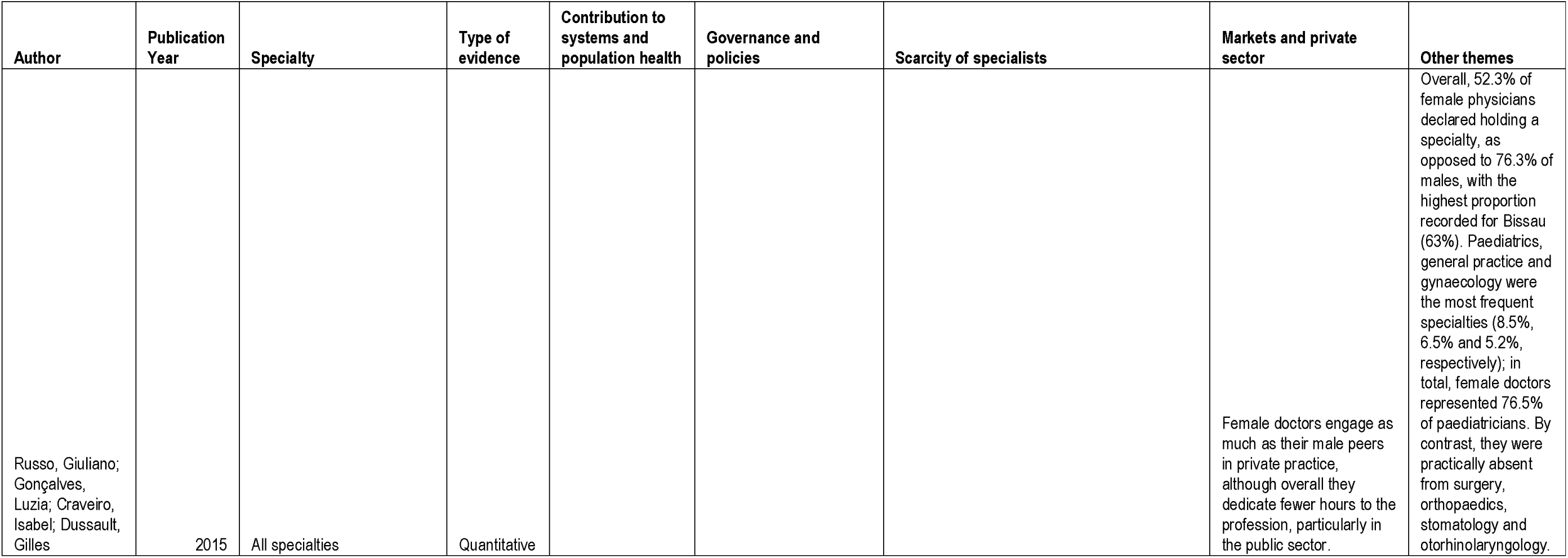
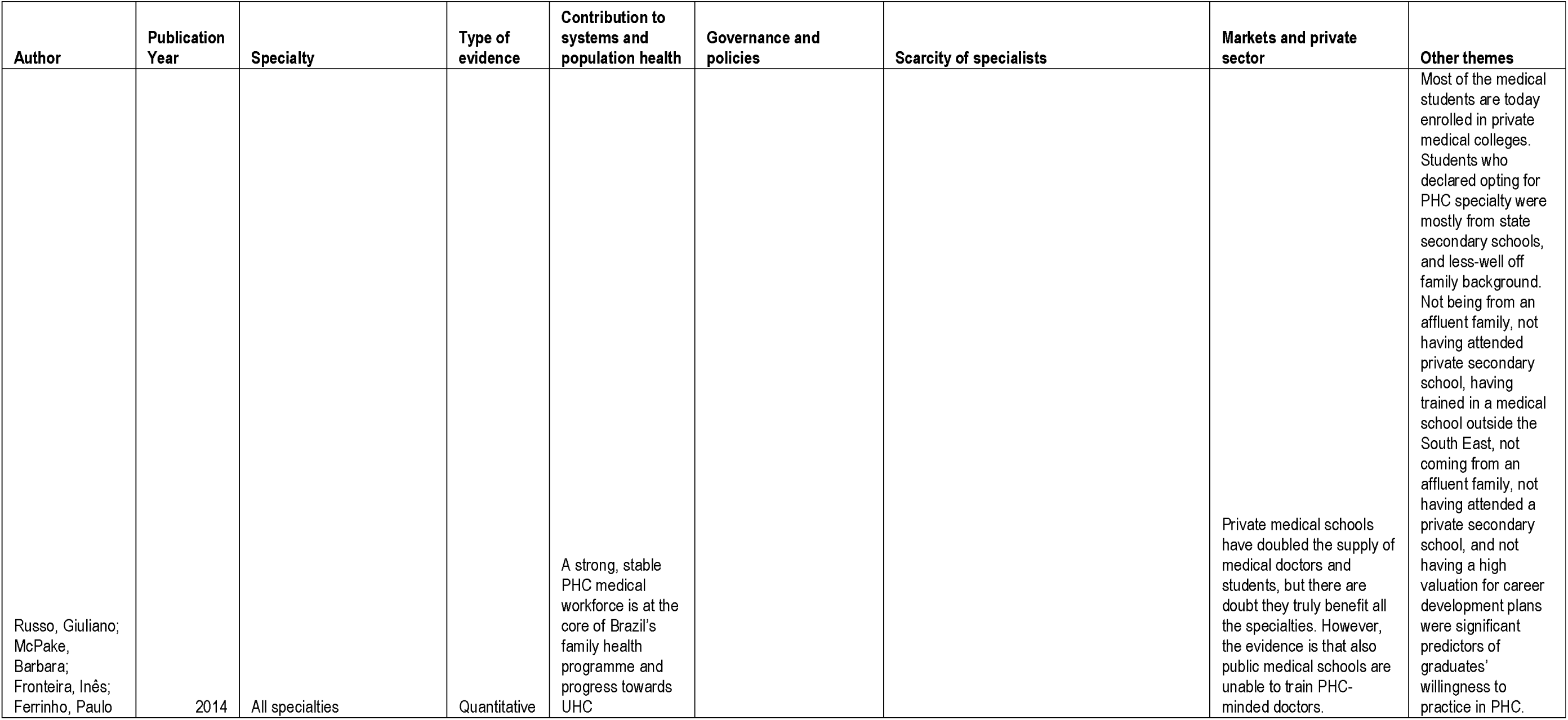
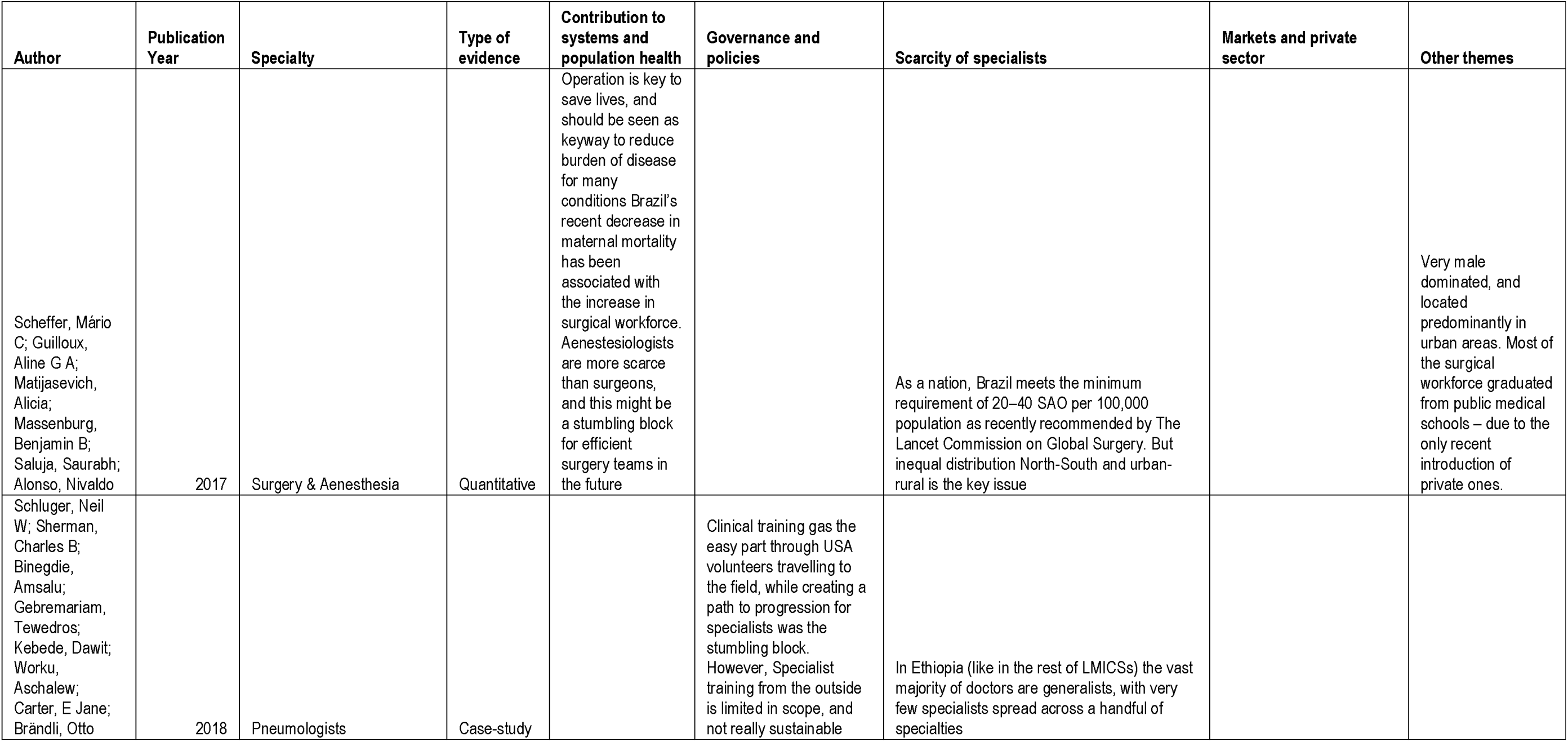
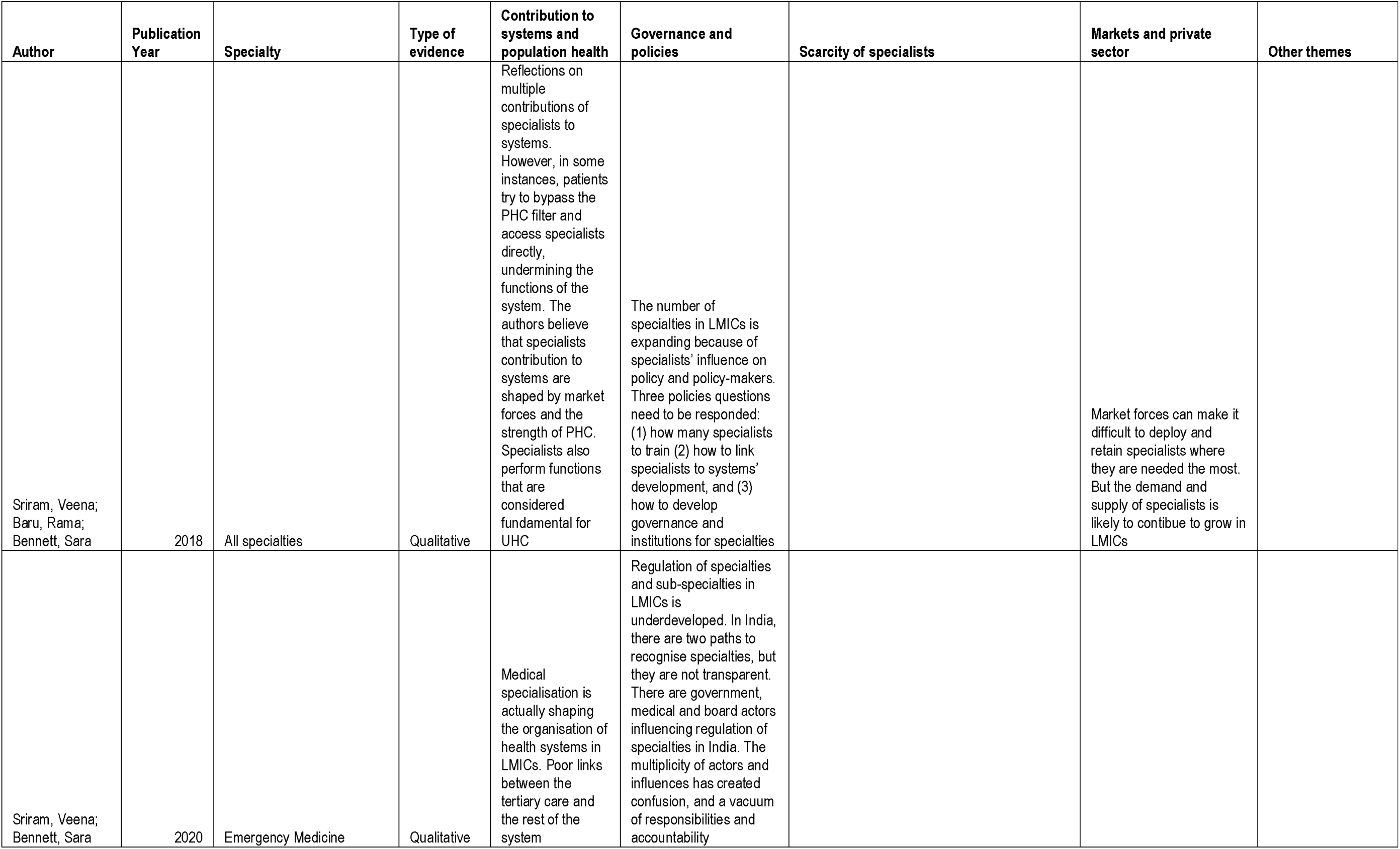
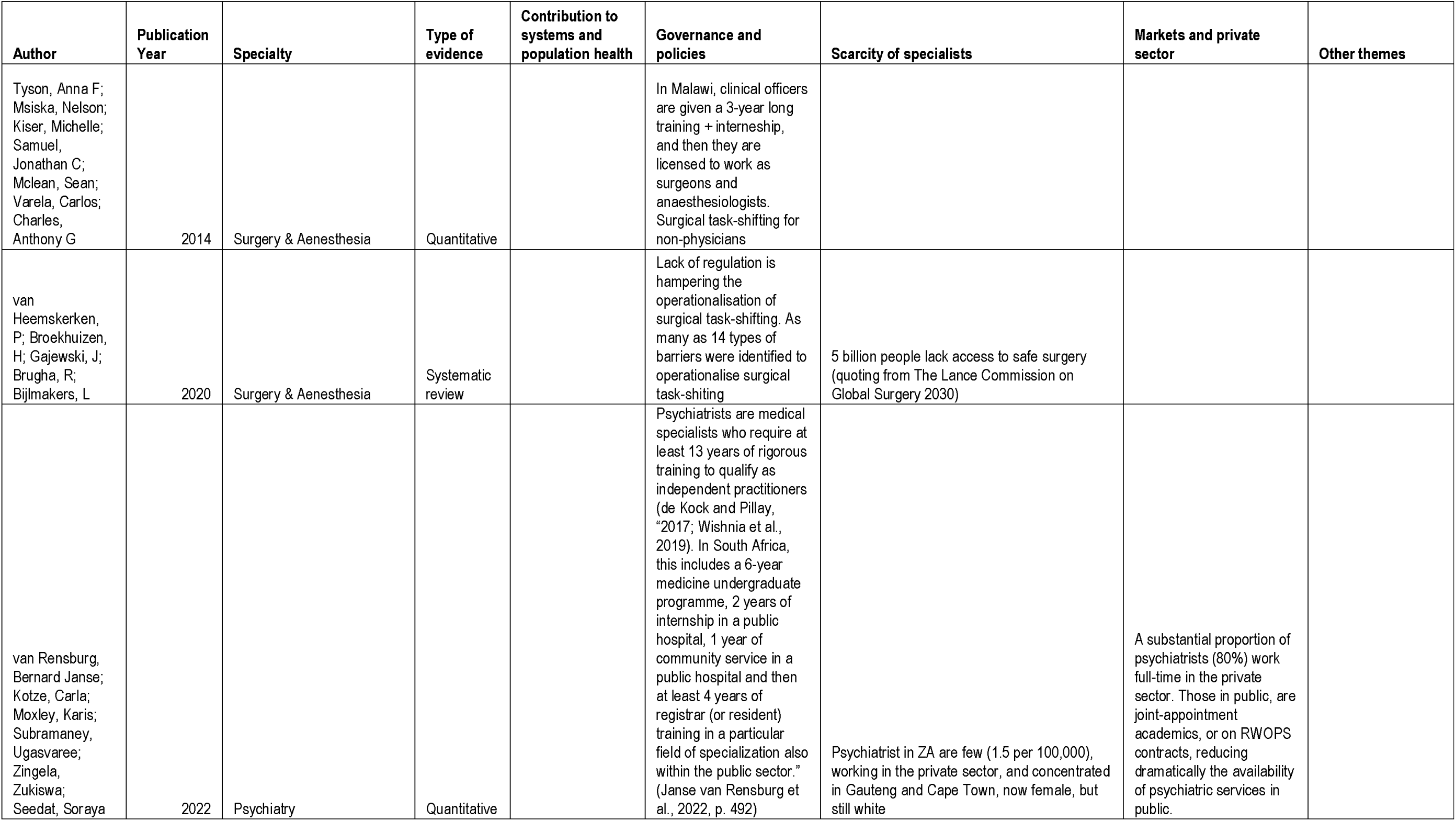
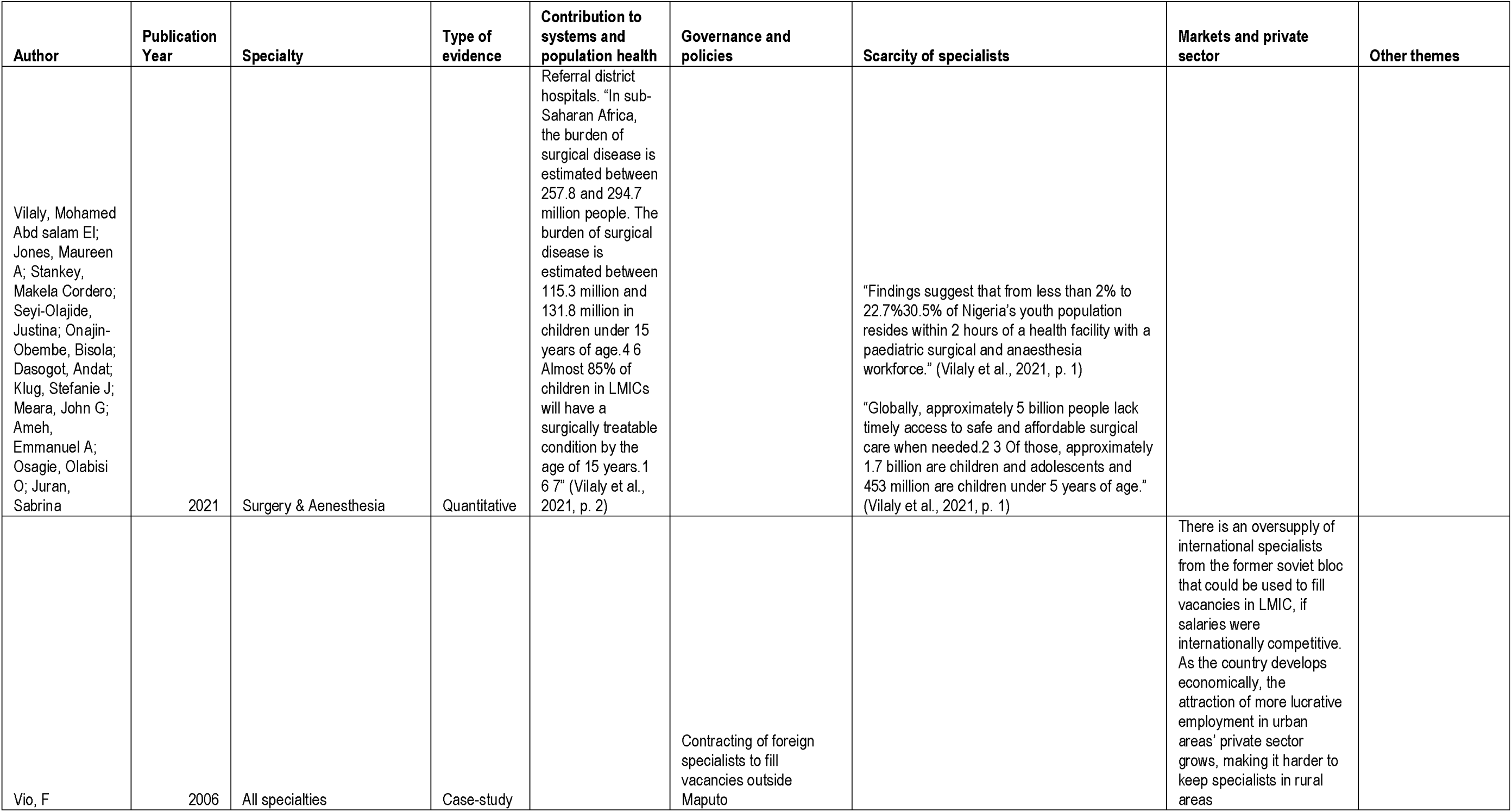
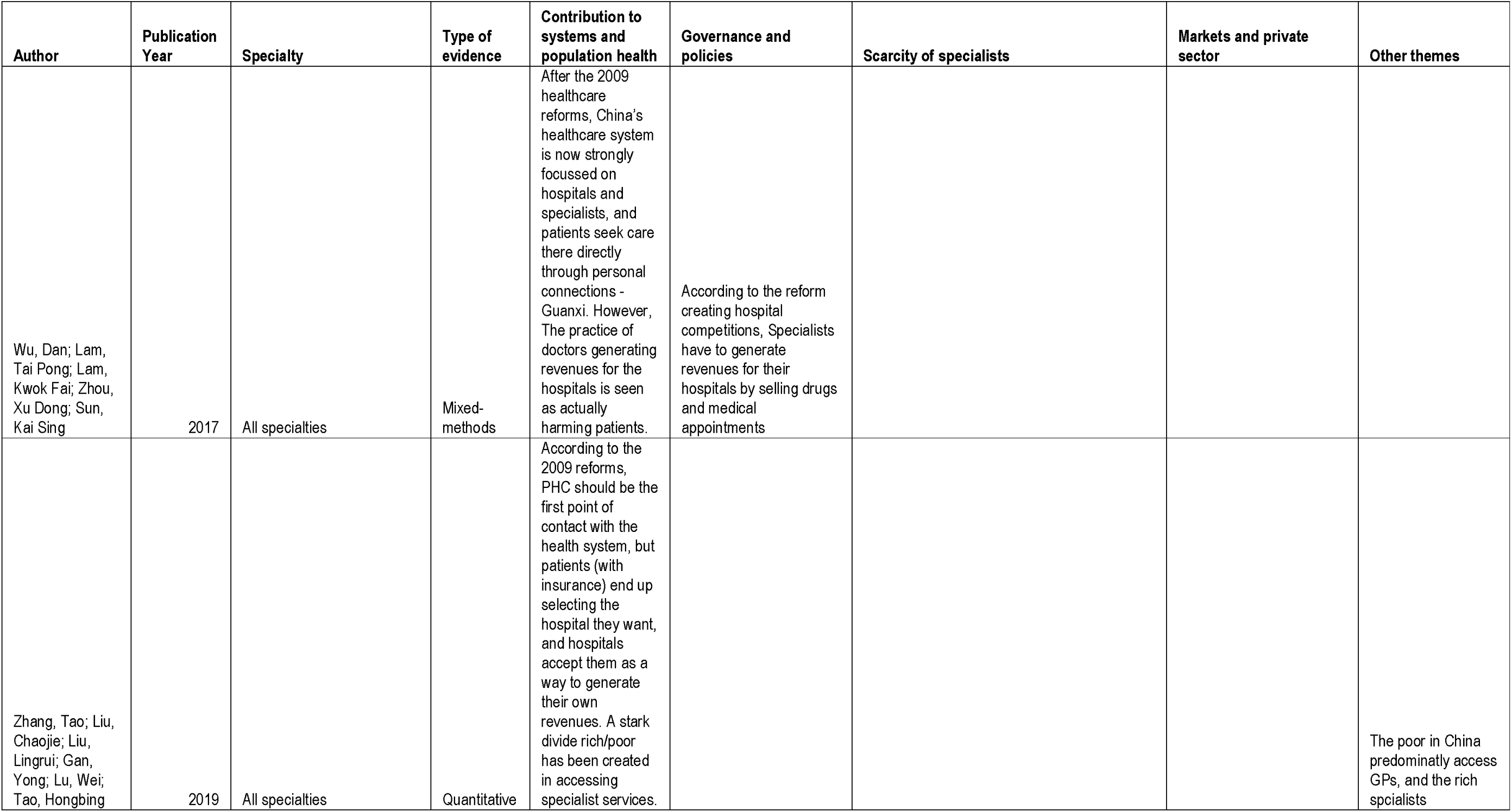
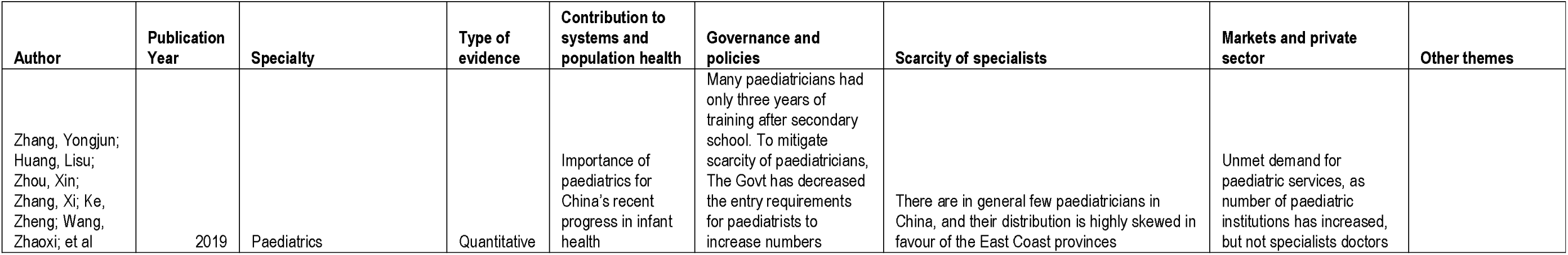
Full evidence table.

### 3.1 Evidence on scarcity and distribution of specialists

As many as 36 papers provided (mostly quantitative) evidence on scarcity of medical specialists in low-income countries, with many of such works (19) focussing on surgeons and anaesthetists (Table 1). Key papers for this area were the ones originating from the Lancet Commission on Global Surgery [4], that showed how, in low-income and lower-middle-income countries, nine people out of ten cannot access basic surgical care. Meara et al demonstrates how 143 million additional surgical procedures would be needed in LMICs each year to save lives and prevent disability, as only 6% of the world’s procedures occur in the poorest countries. The commission calculated that unmet surgical needs are the greatest in easter, western, and central Sub-Saharan Africa, and south Asia [4].

Building on the Lancet Commission’s estimates, in their modelling study Daniels and colleagues calculated that 94.9% of world countries have a density of surgeons lower than 20 surgeons/100,000 population, with the Sub-Saharan Africa region, East Asia and Pacific, Latin America and Caribbean, Middle East and North Africa, and South Asia the regions with more countries with sub-optimal surgeons’ density (<20 surgeons/100,000 population) [24]. In the 20 most populous countries evaluated (154, including both HICs and LMICs), the lowest surgeon densities were observed for Nigeria (1.4), Uganda (0.9), Ethiopia (0.6), Tanzania (0.3), and Democratic Republic of the Congo (0.2).

Henry and colleagues quantified the surgical capacity in Malawi’s hospitals in 2014, and found that, of the 370 surgical workers they surveyed, 92.7% were non-surgeons and 77% were clinical officers [25]; of the 109 anaesthesia providers, 95.4% were non-physician anaesthetists. Non-surgeons and anaesthesia clinical officers were the only providers of surgical services and anaesthetic services in 85% and 88.9% of hospitals, respectively, with no specialist serving in the district hospitals. Another study focussing on anaesthesia found that Uganda has 68 specialist physicians and approximately 600 non-physician anesthetist providers for 43 million people, with most providers located in Kampala and other urban centres [26].

As for scarcity of psychiatry specialists, van Rensburg and colleagues found in South Africa the number of active qualified psychiatrists in 2019 working in the public and private sectors was 850, giving a ratio of 1.53 of psychiatrists per 100 000 population [27]. Furthermore, of the total number of psychiatrists, close to 80% (n = 625) were working full time in the private sector, while only about 20% (n = 168) were working full time in the public sector. Another study on psychiatry capacity in Sierra Leone [28] calculated a 98% treatment gap for severe mental disorders nationwide, as there were only two psychiatrists and 20 psychiatric nurses in the country in 2020.

In their survey of the national cardiology workforce in China, Gong and colleagues found there were 25,240 cardiologists in the mainland in 2016; the ratio to population was 19 per million, which compares poorly with the ratios of around 50 per million in high-income countries [29]. Another study on paediatricians in China [30] talks of a full-blown crisis facing paediatric care services in China, with a single physician is typically responsible for 80 to 100 visits per day in some tertiary hospitals, with an average of .50 work hours per week. The same study reports how in 2006, the Paediatric Society of Chinese Medical Doctors Association reported 831 incidents of serious medical violence, including 319 attacks on paediatricians, with many being beaten to death or disabled.

### 3.2 Specialists’ contribution to population health

Overall, we found vast and consolidated evidence on the impact of specialist services on population health in LMICs, particularly for surgeons and anaesthesiologist, obstetricians, and cardiologists (Table 1).

Daniels et al (2020) reports that 90% of maternal mortality could be averted worldwide with timely surgical intervention, 32% of the global burden of disease requires surgical decision making; and that 77.2 million disability-adjusted life years could be averted [24]. Falk’s et al (2020)’s literature review finds evidence that up to 32% of the global burden of disease is surgical in nature, but 5 billion people on our planet have minimal access to surgical services and 2 billion have no access to basic surgical services [31]. Such finding is supported by Mock and colleagues, calculating that provision of essential surgical procedures would avert about 1·5 million deaths a year, or 6-7% of all avertable deaths in low-income and middle-income countries [32].

Based on the Lancet Commission’s global surgery dataset, Meara and colleagues (2016) calculate there are particularly steep improvements in maternal survival when specialist providers per 100 000 population are improved from 0 to 20; beyond densities of 40 per 100 000 population, gains are still present but the gradient of the curve is flatter, displaying diminishing returns of lives saved [4].

A doctors panel data study from Mexico [33] found the density of specialist physicians to be negatively associated with amenable mortality rate, as a unit increase in density of such doctors was associated with a 0.5% reduction in the overall amenable mortality. Also Davies and colleagues found an inverse linear relationship between density of anaesthesia providers and maternal mortality in LMICs [34]. On the same theme, the decrease of maternal mortality during the last decade in Brazil was associated with the contingent increase of the country’s surgical workforce [35].

Vilaly’s study on paediatric surgery reports that, in Sub-Saharan Africa, the burden of surgical disease is estimated between 115.3 million and 131.8 million in children under 15 years of age, and that 85% of children in LMICs will have a surgically treatable condition by the age of 15 years [36]. A smaller study on gynaecologic oncology in Ghana [37] reports that, in 2018, cervical cancer was the leading cancer in half of the countries in SSA and responsible for 21.7% of all cancer deaths in SSA women; a lack of trained gynaecologic oncology specialists contributes to poor patient outcomes, with a critical need for specialists to lead diagnosis, imaging, and surgical and oncologic care.

### 3.3 Specialists’ role in national health systems

The evidence on medical specialists’ contribution to health systems is comparatively patchier and fragmented (Table 1). Sriram and Bennet (2022) mention how, in high- as well as low-income settings, specialists help organise and lead systems of academic medicine, advance teaching and research, and play a major role in developing sector’s policies [2]. Such view is corroborated by a case-study from Mozambique [38] where, specialists initially employed only for teaching and supervision, later committed to developing the national medical research system, mentorship in bio-clinical science, regional networks for mutual support, attracting research funds, and establishing national research ethics institutional capacity.

Specialist services are typically an essential part of national curative systems; in China, after the 2009 reform, the health system has become crucially hinged to hospitals and specialists because of the current insurance-based financing system. However, Wu et al (2017) shows how such centrality has come at the expense of primary care services, with patients trying to bypass such parts of the system perceived as lower quality, and seek directly more sophisticated and more expensive specialist services [39].

Interviews and survey-based studies from Cape Verde, Guinea-Bissau, and Mozambique show how specialists are crucial for the development of private services in urban areas, expanding availability and diversity of healthcare services, catering for different sections of the population, and therefore increasing overall patients’ welfare [40,41].

Sriram and Bennet identify a recurrent trend in urban areas within LMICs with better specialist supply, experiencing a ‘bypassing’ of primary care, with patients directly seeking specialist care [2]. Such a phenomenon, where specialists work outside or in parallel to a primary care-led healthcare system, raises major concerns around efficiency, overmedicalisation, affordability and possibly, quality [42].

A debate exists on whether some of specialist, life-saving services should be located closer to communities, such as for basic surgical interventions in First (district) Referral Hospitals [43]. English and colleagues [43] make the argument that overproduction of and overreliance on specialists in Sub-Saharan African countries risks fragmenting care and undermining the functions of First Referral Hospitals (FRH), reduce the system’s resilience, and its ability to deal with multi-morbidity. In the same vein, Atiyeh and colleagues’ review argues that for years, surgeries were not planned considering similarities and disparities between developed or urban areas and rural and remote areas [44], and conclude that in LMICs surgery might have been thought to lie outside the scope of public health.

Less visible specialties such as histopathology, are also essential for medical tests and the functioning of laboratories, without which diagnostic systems could not truly work. To this respect, a qualitative study from Kwa Zulu-Natal, South Africa [45] describes how such specialty is perceived as comparatively less prestigious, its specialists subject to the attraction of the private sector, and therefore finding it particularly difficult to recruit and retain specialists in key parts of public health systems.

Specialists very often take charge of management and oversight responsibility [46]. However, in a case-study from Nigeria, Badejo and colleagues see such medical dominance as deleterious to collaboration across different hospital professions [47], while Binyaruka and colleagues show that such dominance helps specialists in Tanzania to access informal payments [48].

### 3.4 Evidence on the governance of specialties

The literature highlights different functions involved in governance of medical specialties, such as: Identification/approval of specialties; Seat allotment/forecasting; Approval of training programs; Curricular development; System integration; Continued medical education [46,49]. In their study of ophthalmologists in South Africa, Haastrup and colleagues highlight the absence of official governance systems for the specialty, as specialists are expected to self-govern to maintain the necessary quality of services [50]. Looking at the global anaesthesia workforce, Kemptorne and colleagues find training availability and duration of courses to be inconsistent across world regions [51]. De Vries and colleagues find that subspecialties such as paediatric urology, although being consolidated and registering an excess of professionals in high and middle-income countries, is absent in Africa, as urological care on the continent is provided by general surgeons or paediatric surgeons [52].

In China, the 2006 reform created hospital competition, specialists have to generate revenues for their hospitals by selling drugs and medical appointment; according to Wu and colleagues [53]; this would be distorting specialists governance, incentives and practice. In their survey of the cardiology workforce in China [29], Gong and colleagues point out the country only has national medical license certification and interventional cardiology license certification, but broader cardiovascular disease license certification did not exist until 2016. As for child and adolescent psychiatry, He Fan et al (2020) found how the sub-specialty is not yet comprehensively regulated in the country, and specialised child psychiatrists are not the only medical professionals providing their service to the population as this role is also fulfilled by adult psychiatrists and other physicians [54]. As for Emergency Medicine (EM), a specialty that is underdeveloped in China, specialists are recruited from regular doctors to deliver emergency care in their own departments [55]; with a view to attracting customers under the new health insurance system, hospitals have built new EM departments and actively advertise such services.

Nigenda and Muños report a generalised perception of weak regulation of medical training in Mexico, where specialist physicians are predominantly trained at public sector health institutions, and subject to political influences from the government and powerful medical councils [56]. In most countries, public doctors’ engagement with the private sector (a phenomenon known as dual practice) is only lightly regulated [57]; in South Africa, where the practice should be governed by the official and Gilson found that doctors do not always ask permission for dual practice [58].

For Belrhiti and colleagues, weak governance of the medical professions in Morocco created an opportunity for unofficial professional hierarchies to form within hospitals [59]. Such arguments are echoed by a study from India on emergency medicine [60], and arriving at the conclusion that regulation of medical specialization is only emerging aspect of health sector governance in LMICs, covering regulatory functions such as recognized categories for specialization, types of practitioners who may acquire skills in certain specializations, approval or accreditation of specialist training programmes, specialist licencing, registration and re-registration, continuing medical education and complaint investigations. Within such a context of weak governance, for Sriram and Bennet (2022) the number of specialties in LMICs is expanding because of specialists’ influence on policy and policy-makers [61].

### 3.5 Specialists and the markets

The literature shows the relation between specialists and market forces is complex and multidimensional; Zhang and colleagues talk about an un-met demand for paediatric services in China [30], Nigenda and Muños about the misalignment between types and numbers of specialists trained, market demand and opportunities for employment in Mexico [56]. For Sriram and Bennet, market forces can make it difficult to deploy and retain specialists where they are needed the most, but the market for specialist services is also likely to accompany economic growth of lower-income countries [2]. For Russo and colleagues, local market conditions are a key determinant of specialists’ engagement with different existing forms of private services [41].

We found comprehensive evidence of specialists’ participation in private services, either through the formal private sector, or within their own public institutions. In Iran Bayat and colleagues found that 48% of public sector specialists were engaged in dual practice [62]. In Brazil, a cross-sectional national survey [63] established that 51.45% were currently working in both the public and private services, while 26.95% and 21.58% were working exclusively in the private and public sectors, respectively; dual practitioners were mostly middle-aged, male specialists with 10 to 30 years of medical practice.

A study on neurosurgeons in LICs [64] shows that over half of neurosurgeons trained within in Sub-Saharan African countries return to SSA countries for professional practice and are involved in dual practice, and less than 10% work exclusively in private practice. Van Rensburg et al (2022) report that most of psychiatrists in South Africa (80%) work full-time in the private sector. Those in public, are joint-appointment academics, or on Remunerative Work Outside the Public Sector (RWOPS) contracts [65].

We found a wealth of evidence on the influence of market forces on specialists’ location decisions, both within and outside low-income countries. For the 2015 Lancet Commission on Global Surgery, specialist providers are often concentrated in urban areas, which have more surgical infrastructure and better-equipped tertiary care centres than do rural areas [4]. English and colleagues report that in Kenya, despite the government’s efforts to increase the supply of specialists, there has been no concern with the retention of professionals in the public sector, and it is estimated sector [66].

On the other hand, there is substantial evidence of the existence of a dynamic global market for medical specialists, with doctors pushed and pulled to foreign markets attracted by favourable salary differentials, security, and working conditions (Table 1). A study on the diaspora of specialist doctors from Romania [67] finds that most doctors who want to leave the country currently live in underserved or rural areas, with cardiology, surgery, psychiatry, radiology, or anaesthetics the most requested specialists. For Fijian specialists, the attraction of the Australian and New Zealand markets are difficult to resist, particularly for Indo-Fijan doctors looking to escape the island country’s political and economic insecurity [68]. For Palestinian specialists trained abroad [69], only a minority intends never to return to Palestine, while most intend to return after a few years of practice abroad, with the UK as the preferred destination.

However, Vio (2006) also talks about an oversupply of specialists from former Soviet countries and Cuba, that was exploited to staff rural hospitals in Mozambique using doctors recruited internationally [70]. In the same vein, De Vries discusses world market unbalance in the supply of paediatric urologists, pointing to an excess of professionals in some HICs and MICs (including Latin America), and complete lack of such specialists in Africa [52].

## 4 DISCUSSION

We found and reviewed 89 studies examining the relationship between specialists and health systems in LMICs. A significant portion of the literature highlights an absolute or relative scarcity of specialists, particularly surgeons, anaesthesiologists, and psychiatrists. Several studies explored the link between specialists’ availability and burden of disease. Evidence on the broader contributions of specialists to health systems remains less robust; a few studies focus on key system functions by specialists, such as referrals for curative care, hospital management, oversight, training and mentoring of junior colleagues, private sector development, and research. A consensus exists that governance of specialists is highly heterogeneous, characterized by substantial variation in number types of specialties, diversity of training curricula and accreditation systems, and limited regulation of private sector involvement.

Many reports document specialists’ engagement with private health markets, often revealing blurred boundaries between public and private healthcare services. We found evidence of a dynamic global market for specialist medical services, driven by the movement of doctors across national labour markets. The growing corporatization within particular domestic health labour markets, such as India, would be one factor impacting the growth and distribution of specialists [71].

Our expert-driven, a priori framework was broadly validated by the evidence identified in the review. It effectively captures specialists’ roles and highlights the linkages between external influences, health sector governance, healthcare systems, and population health outcomes. However, we found no clear evidence on the mechanisms through which regulation of medical specialties would influence the development of doctors’ roles in health systems or population health. Additionally, the literature did not explicitly examine the contribution of specialists’ private services to population health. Given the growing recognition of private healthcare providers’ role for service availability and achievement of universal health coverage [72–74], a more focused and narrowly scoped review may be necessary to explore the relationship between specialists’ private services and population health outcomes.

It looks like governance of medical specialties within national health systems has remained an underexamined area of health governance, as evidenced by the relatively few papers addressing this topic in our review. The papers we found provide an emerging picture of the complexity of institutional frameworks and organizations involved in governance of specialties [75], the relationships between national and state governments, statutory and/or self-regulatory mechanisms. Further research is needed to better understand the institutional frameworks shaping the development and oversight of medical specialties. These issues are also salient to questions of power and governance processes, given the evidence of elitism and professional hierarchies that emerged within the review [76], exacerbated by the challenges of regulating private healthcare markets in many LMICs [77].

A key debate emerging from our review concerns where life-saving specialist services should be made available within health systems. Due to their complexity and high costs, these services are often placed in urban locations, at the top of the curative care pyramid [1]. However, the *Lancet Global Surgery Commission* highlights that surgical interventions for treatable conditions—such as appendicitis, hernia, fractures, and obstructed labour—should be included in national Universal Health Coverage programs [4]. By the same token, there is ongoing advocacy for integrating psychiatric services into primary health care, enabling early identification and treatment at the community level [78,79]. Yet, at the district level such services are seldom delivered by specialists and are more commonly managed by surgical technical officers [80] or community psychosocial workers [81]. We propose a re-evaluation of where and by whom less complex specialist services are delivered within health systems. First-referral (district) hospitals could serve as their natural hub, offering care closer to communities in less complex settings and improving access for all [43] [82].

Some of our references emphasised the barriers specialists encounter when embarking on their chosen careers in LMICs. In countries like China those wanting to specialise in child psychiatry do not often have appropriate training placements to allow them to become competent in their chosen speciality [54]. Other scholars have written extensively about the difficulties surgeons face when training and attempting to practice across many countries in Sub-Saharan Africa [83]. We consider the barriers faced by specialists to train in their chosen speciality and practice are likely to work alongside market forces to push specialists to regions where both educational and financial opportunities align. The primary challenge, therefore, lies in retaining at least some specialist expertise within domestic public health systems.

Our review highlighted the significant influence of market forces in drawing specialists away from public health systems LMICs, either to wealthier nations or to more lucrative opportunities within the domestic private sector. If governments are unable to offer competitive, near-market salaries for highly qualified health professionals, it seems unrealistic to expect that regulatory measures such as bans would effectively attract specialists to the public sector [84]. A more practical approach may involve allowing limited private practice in exchange for a defined public service commitment, which could better balance these competing demands. Some countries are currently piloting the strategic purchasing of specialist services in large public hospitals [85]. Expanding such initiatives beyond the clinical sphere to encompass other essential health system functions could be beneficial. Encouraging specialists to contribute to national training, mentorship and quality control programs may yield great long-term benefits. In return, allowing these specialists to engage in private clinical practice could serve as an incentive, creating a mutually beneficial arrangement.

Despite the plentiful evidence on scarcity of specialists and lack of governance of medical specialties in LMICs, we encounter only limited literature on policy options to mitigate such gaps. Admittedly, there are studies on effectiveness and barriers of task-shifting specialist functions to physicians and clinical officers (also known as ‘task-sharing’), as a way to mitigate scarcity and weak governance [86], particularly as the practice has already been adopted in many LMICs for obstetrical and basic surgery services [80]. Other reports suggest alternative training modalities for specialists within and outside LMICs, with different retention rates [87]. However, to our eyes such measures appear to at least in part side-step the broader question of whether specialists are a priority investment for health systems in LMICs, and how these should be governed to support the achievement of UHC goals.

We recognize limitations in our systematic review. First, the literature on specialists is heavily dominated by clinical sciences, whereas our primary focus was on health systems and medical professions outcomes. This imbalance resulted in a high number of false positives during our searches, increasing the risk of inadvertently excluding relevant studies. Second, our use of an a priori framework guided both our choice of search terms and data extraction process. While such an approach aligns with the best-fit analysis framework methodology [16,23], we acknowledge it may have introduced some bias in identifying key findings.

Our working definition of ‘specialists’ might have underplayed the importance of family medicine, a recognised specialty in many contexts and key for the achievement of UHC goals [88]. Finally, in addition to employing general terms for medical specialists, we conducted targeted searches for those specialties considered essential by The Lancet NCDI Poverty Commission [18]; we recognise such an approach may have disproportionately inflated the number of hits for certain fields, such as surgery, psychiatry, cardiology, and emergency medicine, while other less common specialties might have been overlooked.

## CONCLUSIONS

Medical specialists are often regarded as the cornerstone of referral systems within health systems and constitute a significant portion of the medical workforce in high-income countries. However, in lower-income settings, specialist services frequently appear misaligned with local health needs, system capacities, and UHC objectives.

Our best-fit framework systematic review examined the available evidence on the role of specialists in health systems and their impact on population health in LMICs. The findings highlight critical shortages of specialists such as surgeons, anaesthesiologists, and psychiatrists, alongside their diverse system roles, including curative care referrals, hospital management, oversight, training and mentoring, research, and private sector development. Governance in this domain was found to be highly variable, with significant differences in the number and types of specialties, training curricula, accreditation systems, and regulation of specialist private practices. Additionally, the existence of a dynamic global labour market for specialists, fuelled by cross-border migration, was frequently noted. Despite this, there remains limited evidence on policies designed to mitigate shortages or improve governance.

Our work provides a conceptual framework for understanding the factors shaping specialists’ engagement with health systems and their impact on population health. However, we conclude that the current deployment of specialists within health systems in LMICs warrants reconsideration, with a focus on aligning their roles more effectively with UHC goals.

## Data Availability

All data produced in the present study are available upon reasonable request to the author

## REFERENCES

1 Britnell M. The role of the ‘specialist’ in healthcare. Clin Med (Lond). 2011;11:329–31. doi: 10.7861/clinmedicine.11-4-329

2 Sriram V, Bennett S. Strengthening medical specialisation policy in low-income and middle-income countries. BMJ Glob Health. 2020;5:e002053. doi: 10.1136/bmjgh-2019-002053

3 McPake B, Squires AP, Mahat A, et al. The economics of health professional education and careers□: insights from a literature review. The World Bank, Health, Nutrition, and Population Global Practice East Asia and Pacific Region 2015.

4 Meara JG, Leather AJM, Hagander L, et al. Global Surgery 2030: evidence and solutions for achieving health, welfare, and economic development. The Lancet. 2015;386:569–624. doi: 10.1016/S0140-6736(15)60160-X

5 The Lancet Editorial. Strengthening primary health care to achieve universal health coverage. The Lancet Regional Health – Europe. 2024;39. doi: 10.1016/j.lanepe.2024.100897

6 Russo G, McPake B, Fronteira I, et al. Negotiating markets for health: an exploration of physicians’ engagement in dual practice in three African capital cities. Health Policy Plan. 2014;29:774–83. doi: 10.1093/heapol/czt071

7 Starfield B, Shi L, Grover A, et al. The effects of specialist supply on populations’ health: assessing the evidence. Health Aff (Millwood). 2005;Suppl Web Exclusives:W5-97–W5-107. doi: 10.1377/hlthaff.w5.97

8 Hanson K, Brikci N, Erlangga D, et al. The Lancet Global Health Commission on financing primary health care: putting people at the centre. The Lancet Global Health. 2022;10:e715–72. doi: 10.1016/S2214-109X(22)00005-5

9 Zhang T, Liu C, Liu L, et al. General practice for the poor and specialist services for the rich: inequality evidence from a cross-sectional survey on Hangzhou residents, China. Int J Equity Health. 2019;18:1–10. doi: 10.1186/s12939-019-0966-6

10 Griswold DP, Makoka MH, Gunn SWA, et al. Essential surgery as a key component of primary health care: reflections on the 40th anniversary of Alma-Ata. BMJ Global Health. 2018;3:e000705. doi: 10.1136/bmjgh-2017-000705

11 Bhaumik S, Norton R, Jagnoor J. Structural capacity and continuum of snakebite care in the primary health care system in India: a cross-sectional assessment. BMC Prim Care. 2023;24:160. doi: 10.1186/s12875-023-02109-2

12 Giovanella L, Mendoza-Ruiz A, Pilar A de CA, et al. Universal health system and universal health coverage: assumptions and strategies. Ciênc saúde coletiva. 2018;23:1763–76. doi: 10.1590/1413-81232018236.05562018

13 Miseda MH, Were SO, Murianki CA, et al. The implication of the shortage of health workforce specialist on universal health coverage in Kenya. Hum Resour Health. 2017;15:80. doi: 10.1186/s12960-017-0253-9

14 Nigenda G, Muños JA. Projections of specialist physicians in Mexico: a key element in planning human resources for health. Human Resources for Health. 2015;13:79. doi: 10.1186/s12960-015-0061-z

15 Kutzin J, Sparkes SP. Health systems strengthening, universal health coverage, health security and resilience. Bull World Health Organ. 2016;94:2. doi: 10.2471/BLT.15.165050

16 Carroll C, Booth A, Leaviss J, et al. “Best fit” framework synthesis: refining the method. BMC Medical Research Methodology. 2013;13:37. doi: 10.1186/1471-2288-13-37

17 Pieper D, Rombey T. Where to prospectively register a systematic review. Systematic Reviews. 2022;11:8. doi: 10.1186/s13643-021-01877-1

18 Bukhman G, Mocumbi AO, Atun R, et al. The Lancet NCDI Poverty Commission: bridging a gap in universal health coverage for the poorest billion. The Lancet. 2020;396:991–1044. doi: 10.1016/S0140-6736(20)31907-3

19 OECD. DAC List of ODA Recipients. 2024. https://www.oecd.org/dac/financing-sustainable-development/development-finance-standards/daclist.htm (accessed 27 June 2024)

20 Sawyer SM, McNeil R, Francis KL, et al. The age of paediatrics. The Lancet Child & Adolescent Health. 2019;3:822–30. doi: 10.1016/S2352-4642(19)30266-4

21 Wise J. Life as a physician in general internal medicine. BMJ. 2020;368:m19. doi: 10.1136/bmj.m19

22 Joanna Briggs Institute. Critical Appraisal Tools. 2021. https://joannabriggs.org/critical-appraisal-tools (accessed 3 February 2021)

23 Carroll C, Booth A, Cooper K. A worked example of ‘best fit’ framework synthesis: A systematic review of views concerning the taking of some potential chemopreventive agents. BMC Medical Research Methodology. 2011;11:29. doi: 10.1186/1471-2288-11-29

24 Daniels KM, Riesel JN, Verguet S, et al. The Scale-Up of the Global Surgical Workforce: Can Estimates be Achieved by 2030? World J Surg. 2020;44:1053– 61. doi: 10.1007/s00268-019-05329-9

25 Henry JA, Frenkel E, Borgstein E, et al. Surgical and anaesthetic capacity of hospitals in Malawi: key insights. Health Policy Plan. 2015;30:985–94. doi: 10.1093/heapol/czu102

26 Bulamba F, Bisegerwa R, Kimbugwe J, et al. Development of the anaesthesia workforce and organisation of the speciality in Uganda: a mixed-methods case study. South AFRICAN J Anaesth Analg. 2022;28:109–18. doi: 10.36303/SAJAA.2022.28.3.2646

27 van Rensburg BJ, Kotze C, Moxley K, et al. Profile of the current psychiatrist workforce in South Africa: establishing a baseline for human resource planning and strategy. Health Policy Plan. 2022;37:492–504.

28 Fitts JJ, Gegbe F, Aber MS, et al. Strengthening mental health services in Sierra Leone: perspectives from within the health system. Health Policy Plan. 2020;35:657–64. doi: 10.1093/heapol/czaa029

29 Gong Y, Huo Y, CCCP. A survey of national cardiology workforce in China. Eur. Hear. J. Suppl. 2016;18:A1–5.

30 Zhang Y, Huang L, Zhou X, et al. Characteristics and Workload of Pediatricians in China. Pediatrics. 2019;144. doi: 10.1542/peds.2018-3532

31 Falk R, Taylor R, Kornelsen J, et al. Surgical Task-Sharing to Non-specialist Physicians in Low-Resource Settings Globally: A Systematic Review of the Literature. World Journal of Surgery. 2020;44:1. doi: 10.1007/s00268-019-05363-7

32 Mock CN, Donkor P, Gawande A, et al. Essential surgery: Key messages from Disease Control Priorities, 3rd edition. Lancet. 2015;385:2209–19.

33 Nikoloski Z, Albala S, Montero AM, et al. The impact of primary health care and specialist physician supply on amenable mortality in Mexico (2000–2015): Panel data analysis using system-Generalized Method of Moments. Social Science & Medicine. 2021;278:113937. doi: 10.1016/j.socscimed.2021.113937

34 Davies JI, Vreede E, Onajin-Obembe B, et al. What is the minimum number of specialist anaesthetists needed in low-income and middle-income countries? BMJ Global Health. 2018;3:e001005. doi: 10.1136/bmjgh-2018-001005

35 Scheffer MC, Guilloux AGA, Matijasevich A, et al. The state of the surgical workforce in Brazil. Surgery. 2017;161:556–61. doi: 10.1016/j.surg.2016.09.008

36 Vilaly MA salam E, Jones MA, Stankey MC, et al. Access to paediatric surgery: the geography of inequality in Nigeria. BMJ Global Health. 2021;6:e006025. doi: 10.1136/bmjgh-2021-006025

37 Erem AS, Appiah-Kubi A, Konney TO, et al. Gynecologic Oncology Sub-Specialty Training in Ghana: A Model for Sustainable Impact on Gynecologic Cancer Care in Sub-Saharan Africa. Front public Heal. 2020;8:603391. doi: 10.3389/fpubh.2020.603391

38 Noormahomed EV, Mocumbi AO, Preziosi M, et al. Strengthening research capacity through the medical education partnership initiative: the Mozambique experience. Human Resources for Health. 2013;11:62. doi: 10.1186/1478-4491-11-62

39 Wu D, Lam TP, Lam KF, et al. Challenges to healthcare reform in China: profit-oriented medical practices, patients’ choice of care and guanxi culture in Zhejiang province. Health Policy and Planning. 2017;32:1241–7. doi: 10.1093/heapol/czx059

40 Russo G, de Sousa B, Sidat M, et al. Why do some physicians in Portuguese-speaking African countries work exclusively for the private sector? Findings from a mixed-methods study. Human Resources for Health. 2014;12:51. doi: 10.1186/1478-4491-12-51

41 Russo G, McPake B, Fronteira I, et al. Negotiating markets for health: an exploration of physicians’ engagement in dual practice in three African capital cities. Health Policy and Planning. 2014;29:774–83. doi: 10.1093/heapol/czt071

42 Rao KD, Sheffel A. Quality of clinical care and bypassing of primary health centers in India. Soc Sci Med. 2018;207:80–8. doi: 10.1016/j.socscimed.2018.04.040

43 English M. Breaking the silence on first referral hospitals and universal health coverage.

44 Atiyeh BS, Gunn SWA, Hayek SN. Provision of essential surgery in remote and rural areas of developed as well as low and middle income countries. Int. J. Surg. 2010;8:581–5.

45 Ruggunan SD, Singh S. Sector switching among histopathologists in KwaZulu-Natal, South Africa: a qualitative study. Human Resources for Health. 2013;11:23. doi: 10.1186/1478-4491-11-23

46 Jones L, Fulop N. The role of professional elites in healthcare governance: Exploring the work of the medical director. Soc Sci Med. 2021;277:113882. doi: 10.1016/j.socscimed.2021.113882

47 Badejo O, Sagay H, Abimbola S, et al. Confronting power in low places: historical analysis of medical dominance and role-boundary negotiation between health professions in Nigeria. BMJ Glob Heal. 2020;5:e003349. doi: 10.1136/bmjgh-2020-003349

48 Binyaruka P, Balabanova D, McKee M, et al. Supply-side factors influencing informal payment for healthcare services in Tanzania. Health Policy Plan. 2021;36:1036–44. doi: 10.1093/heapol/czab034

49 Bauchner H, Fontanarosa PB, Thompson AE. Professionalism, Governance, and Self-regulation of Medicine. JAMA. 2015;313:1831–6. doi: 10.1001/jama.2015.4569

50 Haastrup OOO, Buchan JC, Cassels-Brown A, et al. Are we monitoring the quality of cataract surgery services? A qualitative situation analysis of attitudes and practices in a large city in South Africa. Middle East Afr J Ophthalmol. 2015;22:220–5. doi: 10.4103/0974-9233.151878

51 Kempthorne P, Morriss WW, Mellin-Olsen J, et al. The WFSA Global Anesthesia Workforce Survey. Anesth. Analg. 2017;125:981–90.

52 DeVries CR. A global view of pediatric urology. J Pediatr Urol. 2022;18:271–9. doi: 10.1016/j.jpurol.2022.02.002

53 Wu J. Measuring inequalities in the demographical and geographical distribution of physicians in China: Generalist versus specialist. Int. J. Health Plann. Manage. 2018;33:860–79.

54 He F, Chen S, Ke X, et al. Training status of child and adolescent psychiatrists in China. Eur. CHILD & Adolesc. PSYCHIATRY. 2020;29:83–8.

55 Pei YV, Xiao F. Emergency medicine in China: present and future. World J Emerg Med. 2011;2:245–52. doi: 10.5847/wjem.j.1920-8642.2011.04.001

56 Nigenda G, Muños JA. Projections of specialist physicians in Mexico: a key element in planning human resources for health. Human Resources for Health. 2015;13:79. doi: 10.1186/s12960-015-0061-z

57 Russo G, McPake B, Fronteira I, et al. Negotiating markets for health: an exploration of physicians’ engagement in dual practice in three African capital cities. Health Policy Plan. 2014;29:774–83. doi: 10.1093/heapol/czt071

58 Ashmore J, Gilson L. Conceptualizing the impacts of dual practice on the retention of public sector specialists - evidence from South Africa. Hum Resour Health. 2015;13. doi: 10.1186/1478-4491-13-3

59 Belrhiti Z, Belle SV, Criel B, et al. How medical dominance and interprofessional conflicts undermine patient-centred care in hospitals: Historical analysis and multiple embedded case study in Morocco. BMJ Glob. Heal. 2021;6:e006140.

60 Sriram V, Baru R, Bennett S. Regulating recognition and training for new medical specialties in India: the case of emergency medicine. Health Policy Plan. 2018;33:840–52. doi: 10.1093/heapol/czy055

61 Sriram V, Bennett S. Strengthening medical specialisation policy in low-income and middle-income countries. BMJ Glob. Heal. 2020;5.

62 Bayat M, Salehi Zalani G, Harirchi I, et al. Extent and nature of dual practice engagement among Iran medical specialists. Hum. Resour. Health. 2018;16.

63 Miotto BA, Guilloux AGA, Cassenote AJF, et al. Physician’s sociodemographic profile and distribution across public and private health care: an insight into physicians’ dual practice in Brazil. BMC Health Services Research. 2018;18:299. doi: 10.1186/s12913-018-3076-z

64 Karekezi C, El Khamlichi A, El Ouahabi A, et al. The impact of African-trained neurosurgeons on sub-Saharan Africa. Neurosurg Focus. 2020;48:E4. doi: 10.3171/2019.12.FOCUS19853

65 Janse van Rensburg A, Petersen I, Wouters E, et al. State and non-state mental health service collaboration in a South African district: a mixed methods study. Health Policy and Planning. 2018;33:516–27. doi: 10.1093/heapol/czy017

66 English M, Strachan B, Esamai F, et al. The paediatrician workforce and its role in addressing neonatal, child and adolescent healthcare in Kenya. Arch Dis Child. 2020;105:927–31. doi: 10.1136/archdischild-2019-318434

67 Botezat A, Moraru A. Brain drain from Romania: What do we know so far about the Romanian medical diaspora? East. J. Eur. Stud. 2020;11:309–34.

68 Oman K, Rodgers E, Usher K, et al. Scaling up specialist training in developing countries: lessons learned from the first 12□years of regional postgraduate training in Fiji – a case study. Human Resou^LJ^rces for Health. 2012;10:48. doi: 10.1186/1478-4491-10-48

69 Abukmail E, Albarqouni L. Postgraduate training abroad and migration intentions of medical doctors and students in Gaza: a cross-sectional survey. Lancet. 2021;398:S6. doi: 10.1016/S0140-6736(21)01492-6

70 Vio F. Management of expatriate medical assistance in Mozambique. Hum Resour Health. 2006;4:26. doi: 10.1186/1478-4491-4-26

71 Chakravarthi I, Hunter B, Marathe S, et al. Corporatisation in Private Hospitals Sector in India. Economic and Political Weekly. 2023;58.

72 Mcintyre D, Meheus F, Røttingen J-A. What level of domestic government health expenditure should we aspire to for universal health coverage? *Health Economics*, Policy and Law. 2017;12:125–37. doi: 10.1017/S1744133116000414

73 McPake B, Hanson K. Managing the public–private mix to achieve universal health coverage. The Lancet. 2016;388:622–30. doi: 10.1016/S0140-6736(16)00344-5

74 Coveney L, Musoke D, Russo G. Do private health providers help achieve Universal Health Coverage? A scoping review of the evidence from low-income countries. Health Policy Plan. 2023;38:1050–63. doi: 10.1093/heapol/czad075

75 Sriram V, Baru R, Bennett S. Regulating recognition and training for new medical specialties in India: the case of emergency medicine. Health Policy Plan. 2018;33:840–52. doi: 10.1093/heapol/czy055

76 Friedson E. The reorganization of the medical profession. Med Care Rev. 1985;42:11–35. doi: 10.1177/107755878504200103

77 Dalglish SL, Sriram V, Scott K, et al. A framework for medical power in two case studies of health policymaking in India and Niger. Glob Public Health. 2019;14:542–54. doi: 10.1080/17441692.2018.1457705

78 Davies T, Lund C. Integrating mental health care into primary care systems in low- and middle-income countries: lessons from PRIME and AFFIRM. Glob Ment Health (Camb). 2017;4:e7. doi: 10.1017/gmh.2017.3

79 Jacob KS. Mental health services in low-income and middle-income countries. The Lancet Psychiatry. 2017;4:87–9. doi: 10.1016/S2215-0366(16)30423-0

80 Gajewski J, Monzer N, Pittalis C, et al. Supervision as a tool for building surgical capacity of district hospitals: the case of Zambia. Hum Resour Health. 2020;18:25. doi: 10.1186/s12960-020-00467-x

81 Bolton P, West J, Whitney C, et al. Expanding mental health services in low- and middle-income countries: A task-shifting framework for delivery of comprehensive, collaborative, and community-based care. Glob Ment Health (Camb). 2023;10:e16. doi: 10.1017/gmh.2023.5

82 Jeffries Mazhar R, Willows TM, Bhattarai S, et al. First referral hospitals in low- and middle-income countries: the need for a renewed focus. Health Policy and Planning. 2024;39:224–32. doi: 10.1093/heapol/czad120

83 Gajewski J, Bijlmakers L, Brugha R. Global Surgery – Informing National Strategies for Scaling Up Surgery in Sub-Saharan Africa. International Journal of Health Policy and Management. 2018;7:481. doi: 10.15171/ijhpm.2018.27

84 Alaref J, Awwad J, Araujo E, et al. To Ban or Not to Ban? Regulating Dual Practice in Palestine. Health Systems & Reform. 2017;3:42–55. doi: 10.1080/23288604.2016.1272980

85 Fadzil MM, Wan Puteh SE, Aizuddin AN, et al. Specialists’ Dual Practice within Public Hospital Setting: Evidence from Malaysia. Healthcare (Basel). 2022;10:2097. doi: 10.3390/healthcare10102097

86 Ashengo T, Skeels A, Hurwitz EJH, et al. Bridging the human resource gap in surgical and anesthesia care in low-resource countries: a review of the task sharing literature. Hum. Resour. Health. 2017;15:77.

87 Naidu P, Fagan JJ, Lategan C, et al. The role of the University of Cape Town, South Africa in the training and retention of surgeons in Sub-Saharan Africa. The American Journal of Surgery. 2020;220:1208–12. doi: 10.1016/j.amjsurg.2020.06.070

88 Gupta A, Prasad R, Abraham S, et al. The emergence of family medicine in India–A qualitative descriptive study. PLOS Global Public Health. 2023;3:e0001848. doi: 10.1371/journal.pgph.0001848

